# Nasal systems immunology identifies inflammatory and tolerogenic myeloid cells that determine allergic outcome following challenge

**DOI:** 10.1101/2020.09.09.20189886

**Authors:** Astrid L. Voskamp, Maarten L. Gerdes, Roberta Menafra, Ellen Duijster, Szymon M. Kielbasa, Tom Groot Kormelink, Tamar Tak, Koen A. Stam, Nicolette W. de Jong, Rudi W. Hendriks, Suzanne L. Kloet, Maria Yazdanbakhsh, Esther C. de Jong, Roy Gerth van Wijk, Hermelijn H. Smits

## Abstract

Innate mononuclear phagocytic system (MPS) cells preserve mucosal immune homeostasis. Here, we investigated their role at nasal mucosa following challenge with house dust mite. We combined single cell proteome and transcriptome profiling on immune cells from nasal biopsy cells of allergic rhinitis and non-allergic subjects, before and after repeated nasal allergen challenge. Nasal biopsies of patients showed infiltrating inflammatory HLA-DR^hi^ CD14^+^ monocytes and CD16^+^ monocytes, and transcriptional changes in resident CD1C^+^ CD1A^+^ conventional dendritic cells (cDC)2 following challenge. Importantly, although clinically silent, non-allergic individuals displayed a distinct innate MPS response to allergen challenge: predominant infiltration of myeloid-derived suppressor cells (HLA-DR^low^ CD14^+^ monocytes), as well as cDC2 clusters expressing increased inhibitory/tolerogenic transcripts. Therefore, we identified not only clusters involved in airway inflammation but also a non-inflammatory, homeostatic blueprint of innate MPS responses to allergens in non-allergic individuals. Future therapies should target innate MPS for treatment of inflammatory airway diseases.

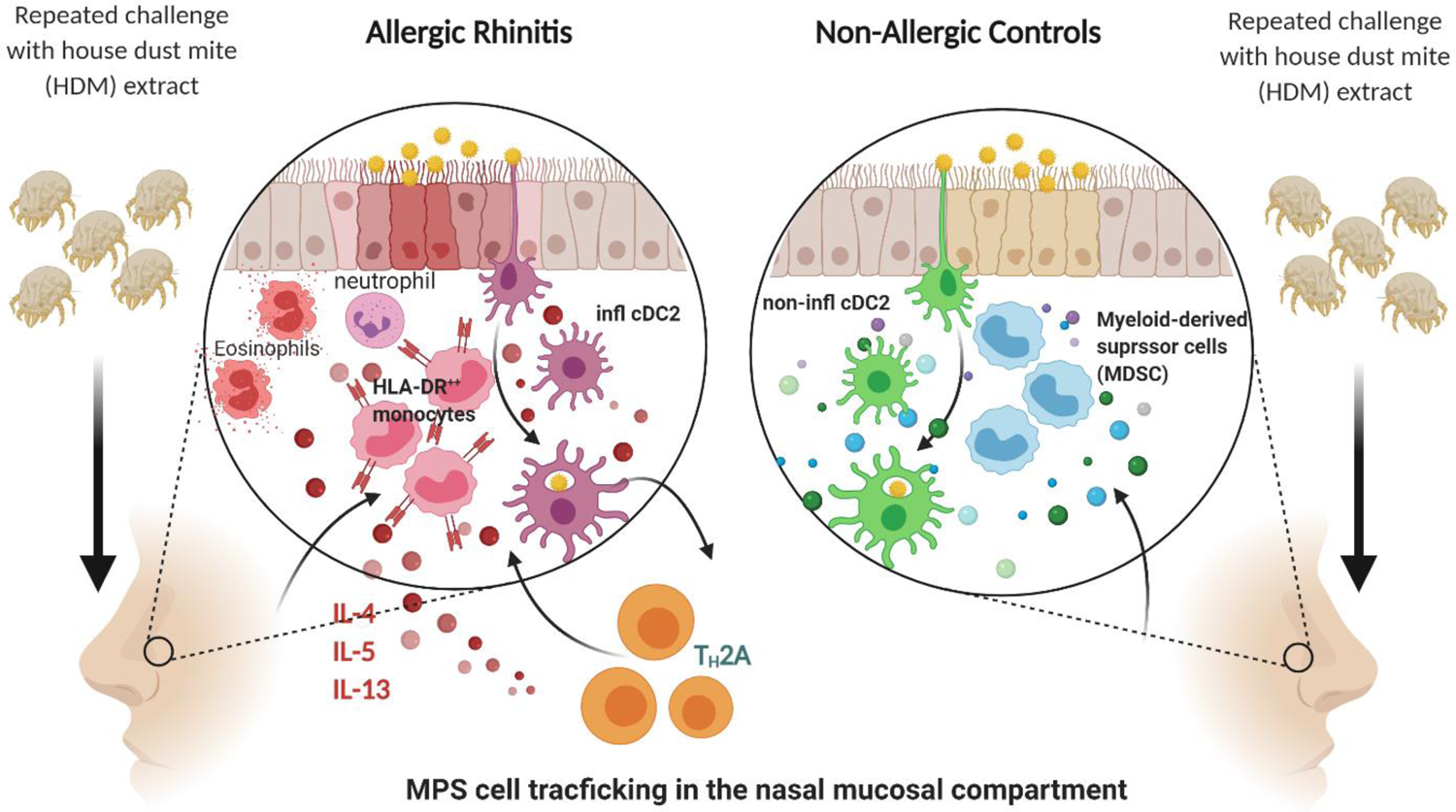

## INTRODUCTION

Innate cells comprising the Mononuclear Phagocytic System (MPS) are key players in guarding immune homeostasis: driving anti-microbial responses to intruders and enhancing anti-inflammatory/regulatory responses preventing tissue damage (*1, 2*). Allergic airway diseases, such as allergic rhinitis (AR) or allergic asthma (AA), are characterized by sensitization to allergens upon inhalation, type 2 immunity and disturbance of local immune homeostasis (*3*).

MPS cells include macrophages (MF), monocytes and dendritic cells (DC); namely CD141^+^ conventional DC (cDC)1, CD1c^+^ cDC2 and plasmacytoid (p)DCs. MF efficiently phagocytose cellular debris and pathogens and induce both inflammatory and anti-inflammatory responses (*4, 5*). Monocytes enter tissue from the circulation (*6*) and are involved in pathogen clearance and wound healing. They can develop into inflammatory monocyte-derived DC (mo-DC) (*7*) and MF, depending on tissue factors and state of inflammation (*8*). In contrast, DC are effective antigen-presenting cells and activate naïve T cells. CDC1 cross-present antigens and prime CD8^+^ T cell responses to intracellular pathogens, while cDC2 preferentially activate CD4^+^ T helper (Th) cells and support their development into e.g. Th1 or Th17 cells to fight viruses, bacteria and/or fungi (*9*). Recent studies suggested that cDC2 are heterogenous in their function and appearance, depending on tissue-specific priming and inflammatory signals (*10*). PDCs produce type I IFNs to viral infections.

Inhalation of harmless environmental substances (including allergens) normally does not result in inflammatory events. Lessons can be learned from the gut, the organ most exposed to microbial substances and dietary products without development of inflammation. Here, local MPS cells produce anti-inflammatory molecules such as retinoid acid, TGFβ and IL-10, promoting immune regulation and homeostasis (*11–14*). Their production is promoted by low dose exposure to TLR ligands, microbial metabolites, or vitamin derivatives (*15*). However, in allergic patients mucosal exposure to harmless allergens instead leads to sensitization and type 2 immunity. Previous studies with repeated grass and tree pollen challenge showed a rapid influx of monocytes in nasal tissue of AR patients, preceding Th2 cells and eosinophils, suggesting an early inflammatory role for MPS cells (*16*).

Much of our knowledge on human MPS cells has been derived from peripheral blood. In mucosal tissue, MPS cells may differ due to the influence of tissue environment and proximity to foreign compounds, commensals, or pathogens. Nasal biopsies provide valuable information on upper airway responses and recent studies using single cell analysis platforms, such as mass-cytometry and single-cell RNA sequencing, have contributed to our understanding of airway epithelial and immune cells in both health and disease states (*17, 18*). Here, we apply those tools following controlled environmental challenge with allergens.

We describe extensive, single cell (sc) proteome and transcriptome profiling on immune cells in nasal tissue of both AR and non-allergic subjects. We studied tissue MPS composition and gene signatures before and after repeated House Dust Mite (HDM) challenge. Our findings showed both emerging inflammatory or tolerogenic MPS cell clusters at nasal mucosa, imposing either allergic symptoms or clinically silent responses, depending on the disease state.

## RESULTS

### Immune cell composition in nasal tissue

Biopsies were obtained from non-allergic and AR subjects (**Table 1**) before and after three days of repeated HDM challenge (**Figure 1A**). Proteomic and transcriptomic profiles of CD45^+^ live cells were measured by mass-cytometry and single cell RNA-sequencing, respectively. In the first level of clustering of the proteomic data, 752,348 cells from 53 samples were divided into 13 lineages (**Figure 1B, S1**) and identified as CD8^+^ T cells, CD8^+^ mucosal associated invariant T (MAIT) cells, CD8^-^ T cells (including CD4^+^ T cells), TCRγδ cells, B cells, innate lymphoid cells (ILC) including natural killer (NK) cells, neutrophils, eosinophils, basophils, mast cells, monocytes, MF/DC, and an unidentified group of cells lacking clear cell lineage markers. In the following round of clustering, subpopulations of each lineage were generated. The transcriptomic data consisted of 46,238 cells from 75 samples, from which 26 cell clusters were generated (**Figure 1C, S2**). Major lineages including CD4^+^ and CD8^+^ T cells, B cells, NK cells, monocytes, DC and mast cells were identified on established markers and enrichment analysis, corresponding with populations detected in the mass-cytometry data (**Table S1**). Granulocyte populations, including neutrophils, eosinophils and basophils were not identified in the transcriptomic data. Cryopreservation prior to scRNA-sequencing will have contributed to the loss of granulocytes, as well as difficulty in their capture with the 10X Genomics Chromium platform (10xgenomics.com).

**Figure 1.**
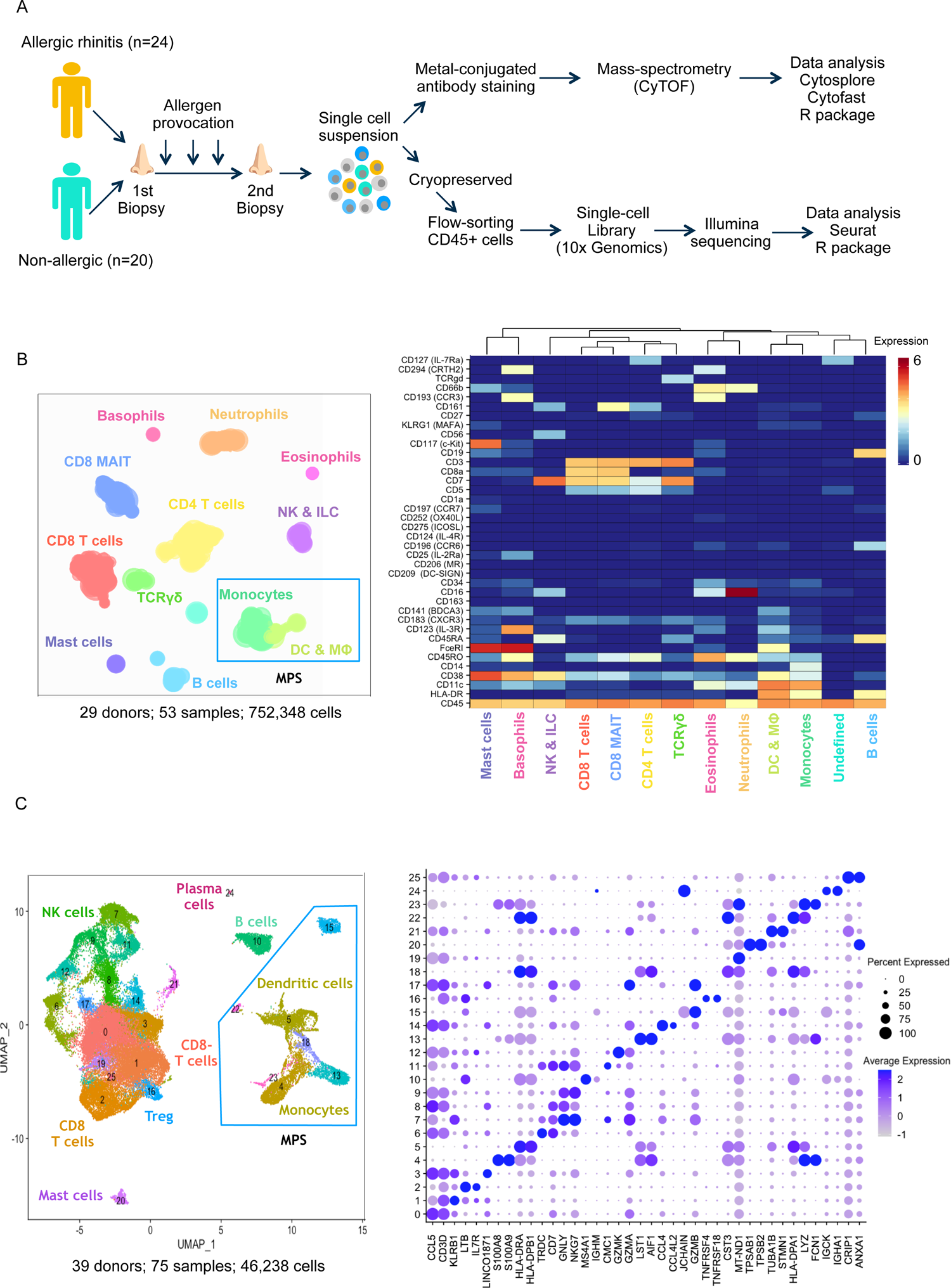
Study set up, cell clustering and lineage identification. A. Schematic of study design and protocol for single cell transcriptomic and proteomic data collection and analysis of nasal biopsy cells from allergic and non-allergic individuals. B. HSNE plot of cell lineages identified in proteomic data and heatmap of markers used. MPS cells, based on HLA-DR and CD11c or CD123, are indicated in the blue box within the HSNE plot. C. UMAP of cell lineages identified in proteomic data and dot plot representing the top two markers of each cluster sorted by log fold change. MPS cells, expressing LYZ and/or CST3 together with HLA-DR/CD74 are indicated in the blue box within the HSNE plot.

**Table 1.**
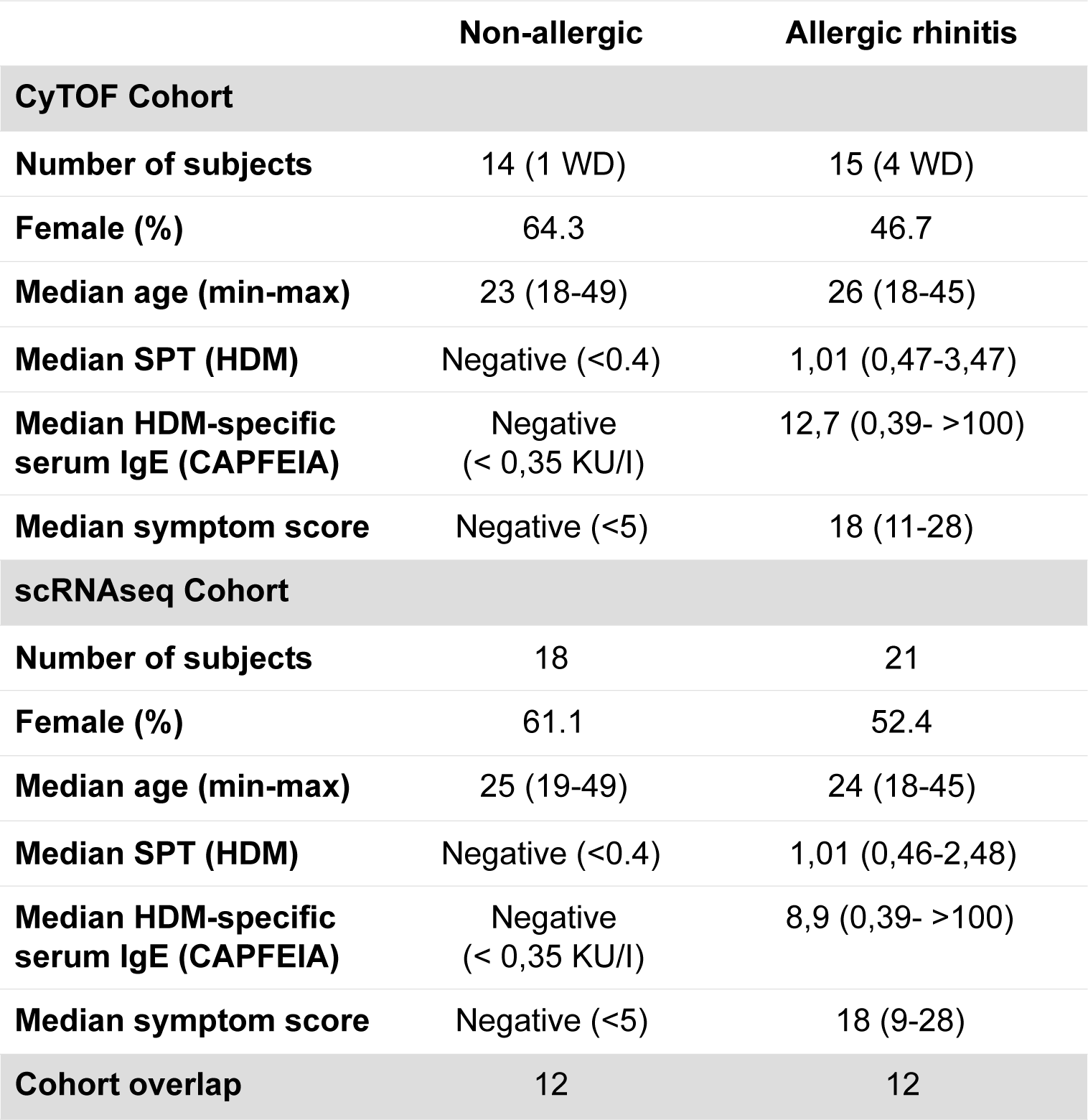
Cohort characteristics

**Table 2.**
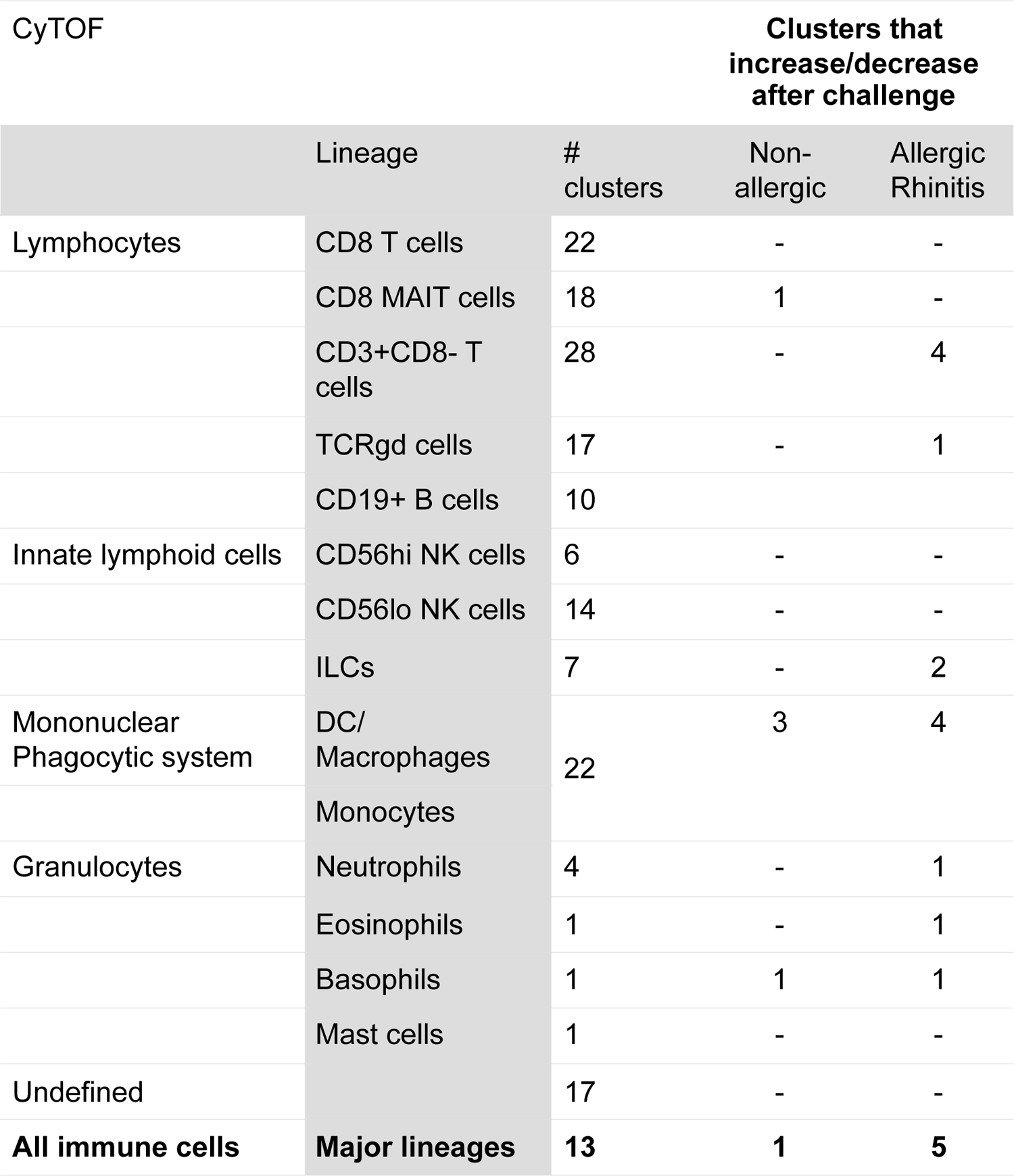

### AR subjects display a type 2 phenotype

Following the first nasal allergen challenge, symptom scores, included sneezing, itchy eyes, nasal congestion, and rhinorrhea, were recorded to monitor allergic responses. AR subjects responded positively to HDM challenge, with a group average summed symptom score of 18 (range 9-28) (*19*). Non-allergic subjects had symptom scores below 5 (threshold for a positive response) (**Table 1**). In allergic subjects, several immunological indications of type 2 responses were found (**Figure S3**): higher levels of the high affinity IgE receptor FCER1A on mast cells (**Figure S3A**); pro-inflammatory TH2A CD4^+^ T cells (*20*) (**Figure S3B**); influx of eosinophils and neutrophils in response to allergen challenge (**Figure S3C**); and higher percentages of ILC2 after allergen challenge (**Figure S3D**). These data clearly show that allergic subjects develop a type 2 signature following allergen challenge, which was distinct from non-allergic subjects.

### Composition of MPS cells in nasal tissue

Since mucosal MPS cells play key roles in maintaining immune homeostasis or driving inflammation, we further explored phenotype and responses of different MPS clusters to allergen challenge in both groups. In the single-cell transcriptomic data, MPS cells were distinguished from other populations based on expression of signature markers LYZ, CST3, AIF1, LST1, and HLA-DRA (**Figure 2A**, **Figure S4**). Top differentially expressed genes of individual MPS clusters were identified (**Table S2**), and each cluster was annotated based on expression of established markers (**Figure S5**) and Enrichr gene set enrichment analysis with the Human Gene Atlas and ARCHS4 Tissues databases (**Figure 2B**). To support cluster annotation, average gene expression of each cluster was compared with signature profiles of MPS cells (**Figure 2C**) from previous (single cell) RNA sequencing studies of peripheral blood and tissue DC, MF and monocytes (*18,21-25*). Cluster 4, 13 and 23 display transcript profiles consistent with monocytes. Cluster 4 corresponds with CD14^+^ monocytes, cluster 13 with CD16^+^ monocytes and cluster 23 with monocytes with a cytotoxic profile, previously described by Villani *et al.* (*21*), along with CD3 expression (**Figure 2A, 2C**, **Table S2**) which may be an indication of doublet formation. These cells (cluster 4, 13 and 23) clustered separately from DC (cluster 5, 15 and 22) and expressed established monocyte markers S100A8, S100A9, FCN1 and MNDA. We identified CLEC4C^+^ pDC (cluster 15), CLEC9A^+^ cDC1 (cluster 22) and CLEC10A^+^, CD1C^+^ cDC2 (cluster 5) populations. Cluster 5 gene expression profile correlated with that of several distinct cell types including cDC2, Langerhans cells and ‘AS’ DC (*21, 26*) (**Figure 2C**, **Table S2**). Cluster 18 could be identified as MF expressing APOC1, APOE and MRC1, along with components of both M1 and M2 profiles **(Figure 2C**), previously described in alveolar MF (*23*).

**Figure 2.**
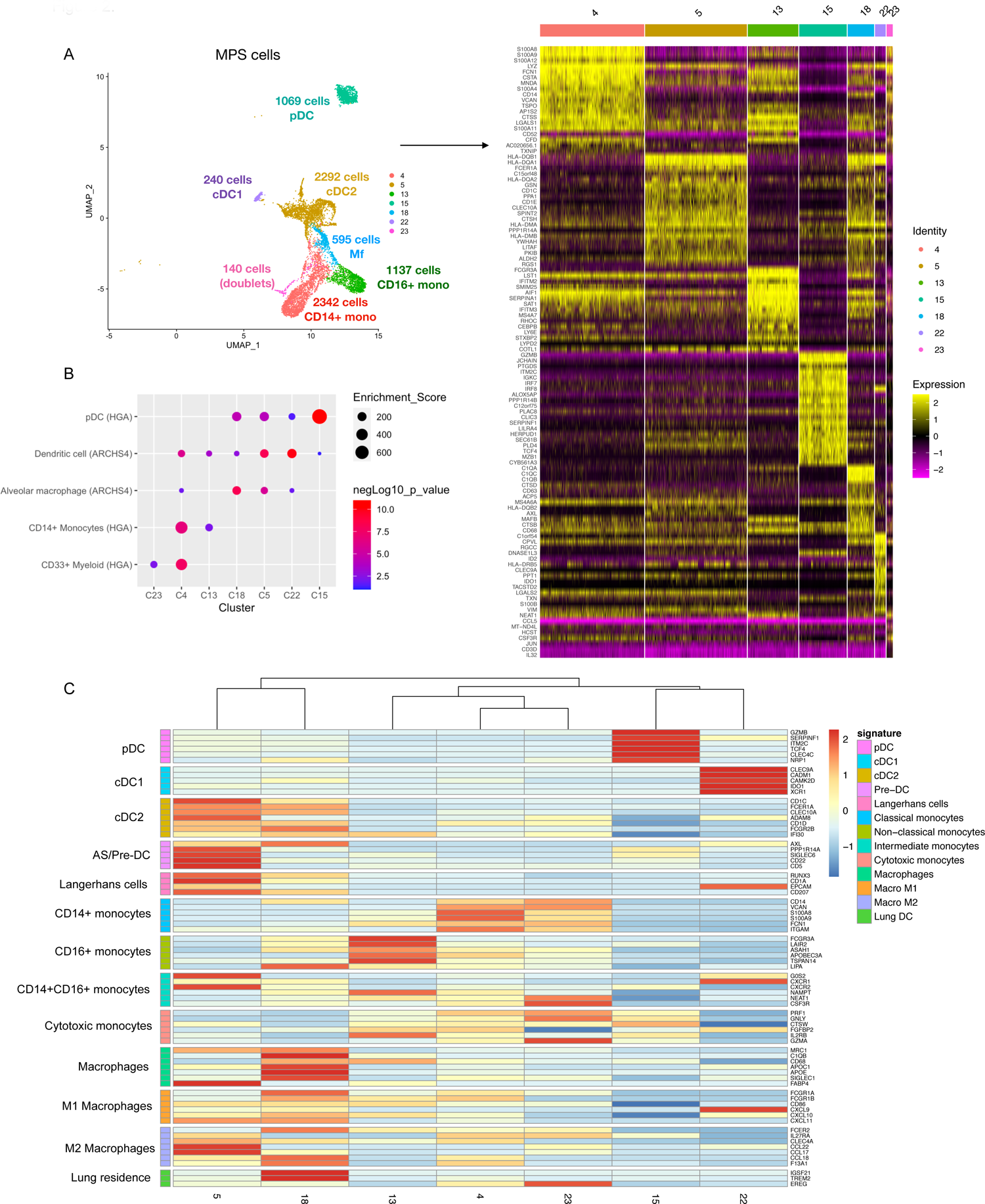
MPS cell cluster annotation in transcriptomic data. A. UMAP of MPS clusters with accompanying heat map of top 10 marker genes sorted by log fold change of each of the clusters within the MPS compartment. B. Dot plot of Gene Ontology Enrichment analysis performed in Enrichr on the top 40 markers obtained for each MPS cell cluster, showing the top combined score for each cluster. Human cell atlas and ARCHS4 Tissue gene set libraries were used for enrichment analysis. C. Heat map of average expression for each MPS cluster (identity listed below) of signature genes (listed on the right) of known cell types (listed on the left).

In the mass-cytometry proteomic data, MPS cells were identified as separate lineages of HLA-DR^+^ cells (**Figure 3A**) expressing MPS cell markers such as CD11c, CD14, CD141 or CD123 (pDC), and lacking B cell marker CD19 (**Figure 1B**). The next level of clustering resulted in 23 MPS cell subpopulations. Based on the markers expressed, populations of CD14^+^ and CD16^+^ monocytes, CD34^+^ progenitor cells, CD206^+^ MF, CD141^+^ cDC1s, FCER1A^+^ cDC2s and CD123^+^ pDCs could be identified (**Figure 3B**). Overall, the populations in the proteomic data corresponded well with the major populations identified in the transcriptomic measurements, i.e. CD14^+^ and CD16^+^ monocytes, pDC, cDC1, MF and multiple cDC2 clusters (**Figure 3C**). Therefore, combination of proteomic and transcriptomic data will augment the analysis of (sub)clusters of MPS cells with low cell numbers and strengthen conclusions.

**Figure 3.**
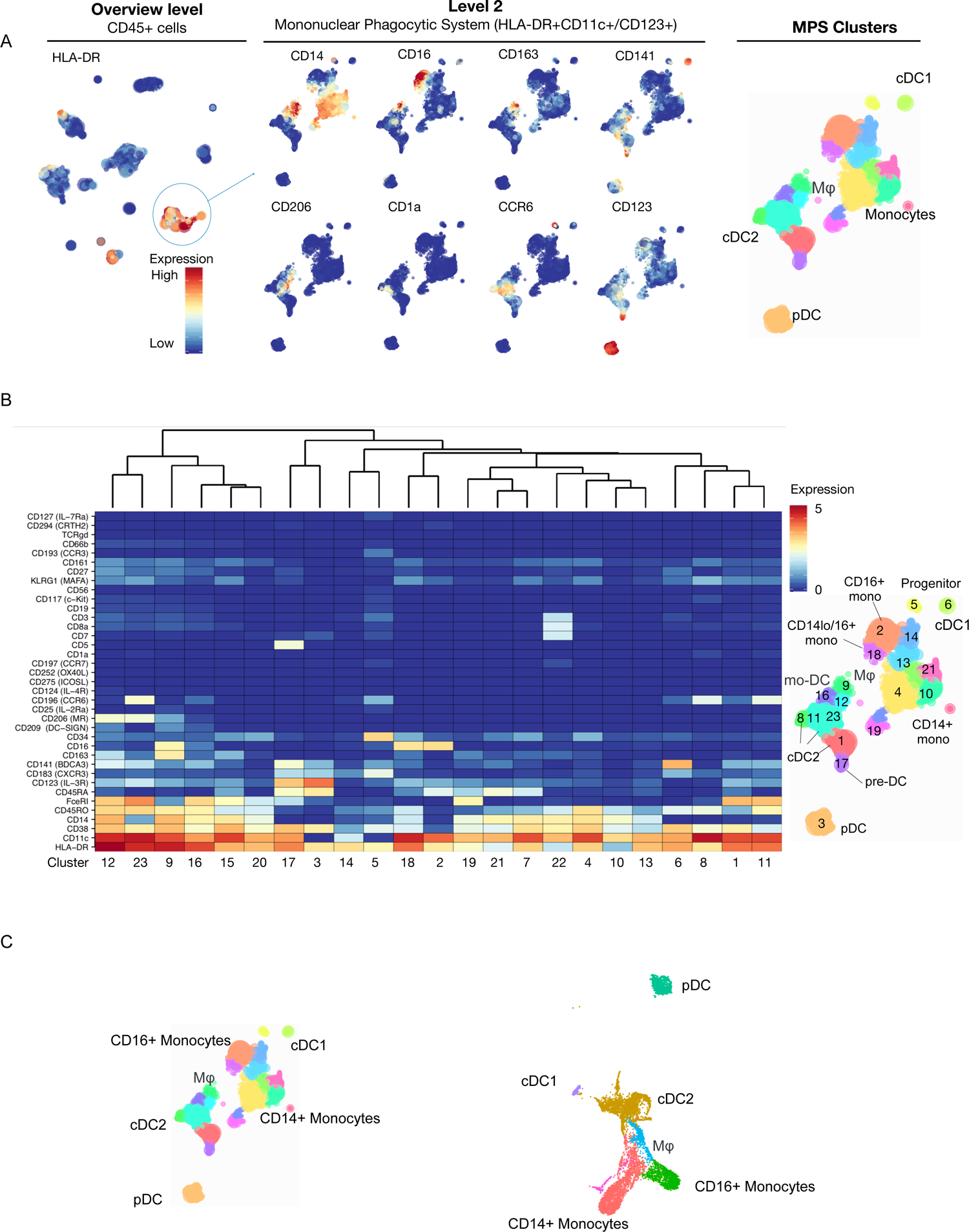
MPS cell cluster annotation in proteomic data. A. MPS cell location in HSNE plot and expression of relevant markers for major MPS cell type identification. B. Heat map of cell marker expression in proteomic data with identification of DC, monocyte and macrophage populations. C. Alignment of major MPS cell populations between proteomic and transcriptomic data sets.

### Recruitment of different monocyte populations in response to allergen challenge

In response to allergen challenge, we observed significant increases in percentages of CD14^+^ monocytes in both non-allergic and allergic subjects in the transcriptomic data (**Figure 4A**). Further clustering of CD14^+^ monocytes (cluster T4) revealed five sub-clusters (**Figure 4B and 4C**, **Table S3**) which could be distinguished by HLA-DR expression (**Figure 4C and 4D**): two clusters with high HLA-DR expression (T4.1 and T4.4) and three clusters with a low or intermediate HLA-DR expression (T4.0, T4.2 and T4.3). HLA-DR^hi^ sub-clusters T4.1 and T4.4 expressed higher levels of HLA-related and IFN-related genes compared to other monocyte clusters, corresponding with antigen presentation, cell activation and response to stimuli (**Figure S6**), suggesting a pro-inflammatory role. Furthermore, sub-cluster T4.4 expressed genes related to cell adhesion, migration, proliferation, regulation of ERK1/2 cascade and Th2 cytokine signaling (**Figure S6**, **Table S3**) along with genes of MF profiles (MRC1, APOC1) in some of the cells. The HLA-DR^low^ sub-clusters T4.0, T4.2 and T4.3 expressed higher levels of anti-microbial response genes, such as S100A8 and S100A9 (**Figure 4C and 4E**). Interestingly, S100A9 is associated with immunoregulatory functions in Mo-MF involving ROS and IL-10 production (*27*). Sub-cluster T4.2 expressed the lowest levels of HLA-DRA along with the highest levels of S100A8, S100A9 and S100A12, when compared to all other monocyte sub-clusters (**Figure 4D, 4E**) and is comparable to myeloid-derived suppressor cells (MDSC) (*28*). Sub-cluster T4.3 expressed genes related to RNA translation (RLP36A, RABPC1, EIF3E, EIF3M, ZFP36L2; **Figure S6** and **Table S3**), while cells of sub-cluster T4.0 expressed higher levels of CRIP1 (cysteine-rich intestinal protein (*29*). Overexpression of CRIP1 in transgenic mice resulted in enhanced Th2 cytokines, suggesting a ‘pro-allergic’ role of this protein in T cell differentiation (*30, 31*).

**Figure 4.**
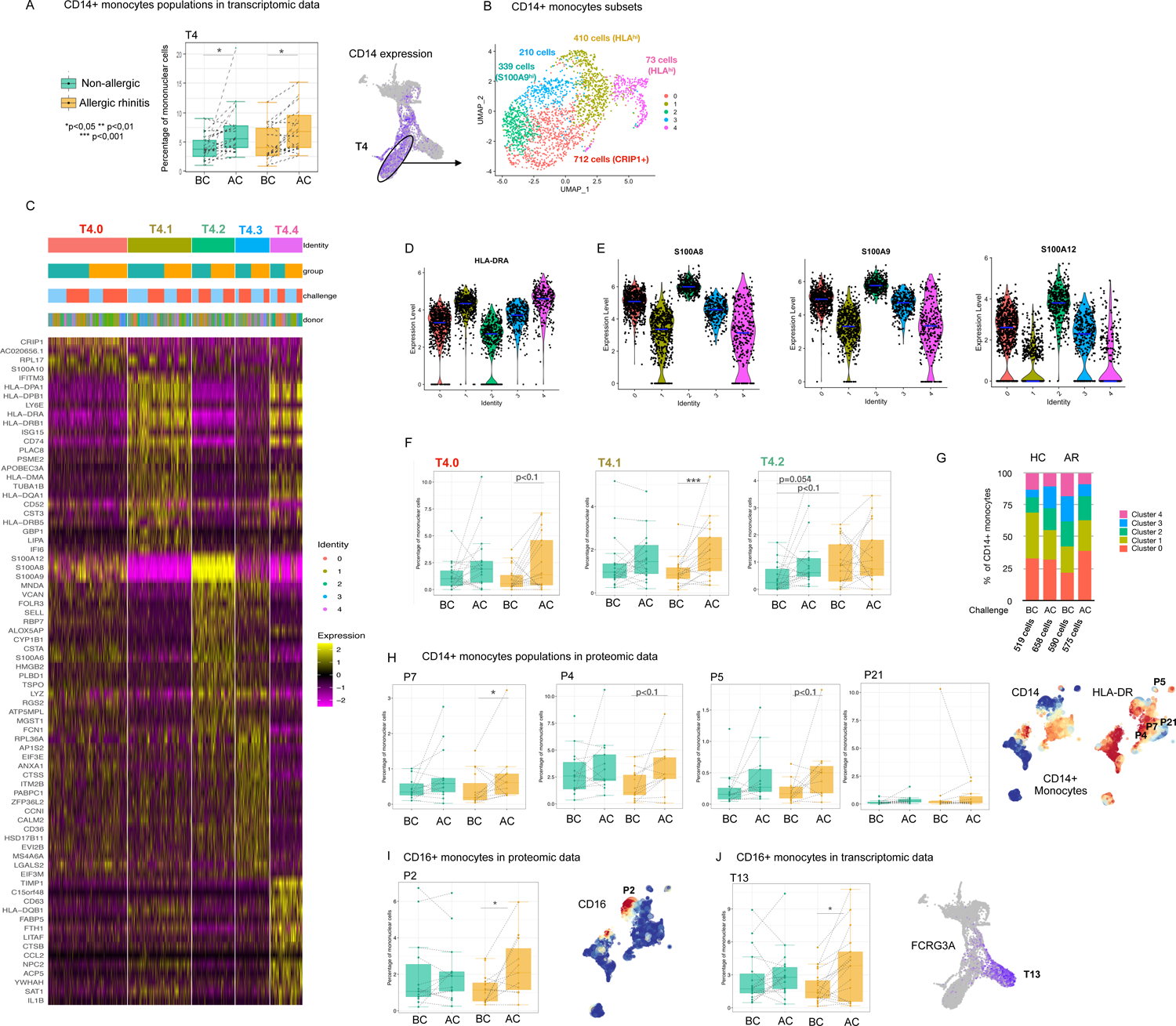
Recruitment of different monocyte populations in response to allergen challenge. A. Box plots of frequency of CD14+ monocytes (Cluster T4) in allergic (orange boxes) and non-allergic individuals (green boxes) before (BC) and after allergen challenge (AC) in the transcriptomic data. * p < 0.05 B. UMAP of five sub-clusters generated from CD14+ monocytes, cluster T4 of the original clusters. Total cell number and distinguishing marker per sub-cluster depicted in the plot. C. Heat map of top 20 gene markers sorted by log fold change of the sub-clusters within the CD14+ monocytes. Bars above heat map indicate composition of clusters based on group (allergic in orange; non-allergic in green), allergen challenge (blue before challenge; red after challenge) and donor. Colours of sub-clusters (identity) correspond with those depicted in UMAP in Figure 4C. D. Violin plot of HLA-DR expression by sub-cluster of CD14+ monocytes. E. Violin plot of S100A8, S100A9 and S100A12 expression by sub-cluster of CD14+ monocytes. F. Box plots of frequency of HLA-DR^hi^ cluster T4.1 and HLA-DR^low^ CD14+ monocyte clusters T4.2 and T4.0 in allergic (orange boxes) and non-allergic individuals (green boxes) before (BC) and after allergen challenge (AC) in the transcriptomic data. *** p < 0.001 G. Composition of CD14+ monocytes by sub-cluster for non-allergic individuals (NA) and allergic individuals (AR), before (BC) and after allergen challenge (AC). Colours of sub-clusters correspond with those depicted in UMAP in Figure 4B. H. Box plots of frequency of HLA-DR^int/hi^ (cluster P4 and P7) and HLA-DR^low^ CD14+ monocytes (cluster P5 and P21) in allergic (orange boxes) and non-allergic individuals (green boxes) before (BC) and after allergen challenge (AC) in the proteomic data. * p < 0.05 I. Box plot of frequency of CD16+ monocytes (cluster P2) in allergic (orange boxes) and non-allergic individuals (green boxes) before (BC) and after allergen challenge (AC) in the proteomic data. * p < 0.05. Expression of CD16 depicted in HSNE, with cluster P2 indicated in the plot. J. Boxplot of frequency of CD16+ monocytes (cluster T13) in allergic (orange boxes) and non-allergic individuals (green boxes) before (BC) and after allergen challenge (AC) in the transcriptomic data. *p < 0.05. Gene expression of FCGR3A depicted in UMAP, with cluster T13 indicated in the plot.

The cell frequency in the HLA-DR^hi^ sub-cluster T4.1 increased significantly in allergic subjects, whereas the cell frequency of the HLA-DRA^low^ sub-cluster T4.2 increased in non-allergic subjects after challenge (**Figure 4F**). Of note, in allergic subjects the sub-cluster T4.2 was abundantly present at baseline, but in contrast to non-allergic individuals, this sub-cluster did not change upon challenge. In contrast, the frequency of the CRIP1^+^ HLA-DR^low^ sub-cluster T4.0 (**Figure 4F**) did show an increasing trend in allergic subjects upon challenge. Lastly, the proportions of each sub-cluster within the CD14^+^ monocytes (T4) suggest that in non-allergic subjects after challenge the HLA-DR^low^ sub-clusters T4.2 and T4.3 form a larger portion of CD14^+^ monocytes compared to before challenge, resulting in relatively fewer HLA-DR^hi^ sub-clusters T4.1 and T4.4 (**Figure 4G**). This is further underlined by significant decreases in ISG15 and IFITM3 gene expression, top gene markers of the HLA-DR^high^ sub-clusters T4.1 and T4.4, in CD14^+^ monocytes of non-allergic subjects **(Table S4**).

Similar to the transcriptomic data, the CD14^+^ monocyte population in the proteomic data was heterogenous and consisted of several sub-clusters, differing in expression of HLA-DR, CD14, CD38, CD11c, CD34, CD45RO and CD45RA (**Figure 3B**). In allergic subjects the percentage of monocytes expressing intermediate HLA-DR and low CD34 levels increased significantly upon allergen provocation (**Figure 4H;** cluster P7). In addition, a similar cluster (P4) along with a small progenitor monocyte population (P5), expressing low HLA-DR and intermediate CD34 levels (**Figure 4H**) showed an increasing trend in allergic subjects (p<0.1). These findings align with the transcriptomic data. In non-allergic individuals, the proteomic data did not show significant increases in any CD14^+^ monocyte cluster, however, similar to the transcriptomic data, visual increases were observed in monocyte clusters P5 and P21 expressing low HLA-DR and CD14 levels (**Figure 4H;** cluster P5), reaching significance prior to multiple comparison correction. Furthermore, significant increases in CD16^+^ monocytes were observed in allergic subjects only after allergen provocation in both the proteomic (**Figure 4I**) and transcriptomic (**Figure 4J**) data sets.

Multiple (sub)clusters could be cross validated in both the proteomic and transcriptomic datasets, supporting differences observed between study participants and/or following challenge, despite the low cell counts in certain (sub)clusters and the loss of significance after multiple comparison correction. Combined, we were able to identify five different subclusters of monocytes in the nasal mucosa that show dynamic changes: following allergen challenge in allergic subjects a predominant increase in pro-inflammatory HLA-DR^hi^ CD14^+^ monocyte and CD16^+^ monocyte clusters, while in non-allergic subjects primarily the frequency of HLA-DR^low^ CD14^+^ monocytes with anti-microbial/regulatory genes increased after allergen challenge.

### Difference between effector and anti-inflammatory cDC2 in allergic and non-allergic subjects

In both the proteomic and transcriptomic data, multiple cell types within the original cDC2 cluster were identified. In the proteomic data four cDC2 clusters (1,8,11,23) were distinguished, along with CD14^+^ DC (16, 12), CD5^+^ CD123^+^ DC (17) and CD163^+^ CD14^+^ CD16^+^ macrophages (9) (**Figure 3B**). In the transcriptomic data, sub-clustering of the cDC2 cluster T5 resulted in seven distinct sub-clusters (**Figure 5A**) that were aligned with recently identified cell types (**Figure 5B-C**) and with some of the proteomic cDC2 clusters (**Figure 5D, S7**), and further characterised by top differentially expressed genes of each sub-cluster (**Figure 5E-F**). Five cDC2 sub-clusters (T5.0-T5.3 and T5.6) were identified as cDC2 cells based on CD1C, FCER1A and/or CLEC10A, while the sub-clusters T5.4 and T5.5 were lacking these markers (**Figure 5-C**).

**Figure 5.**
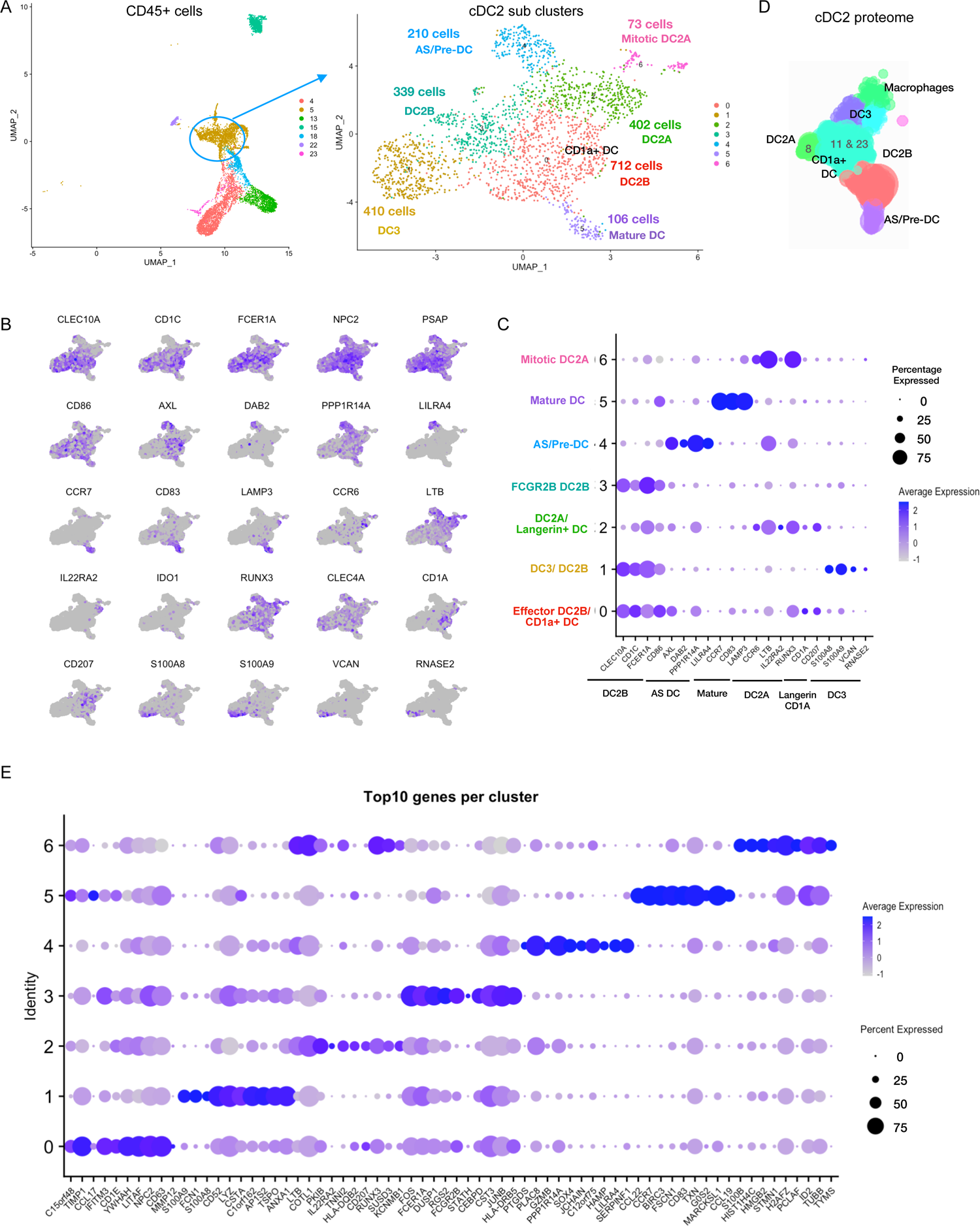
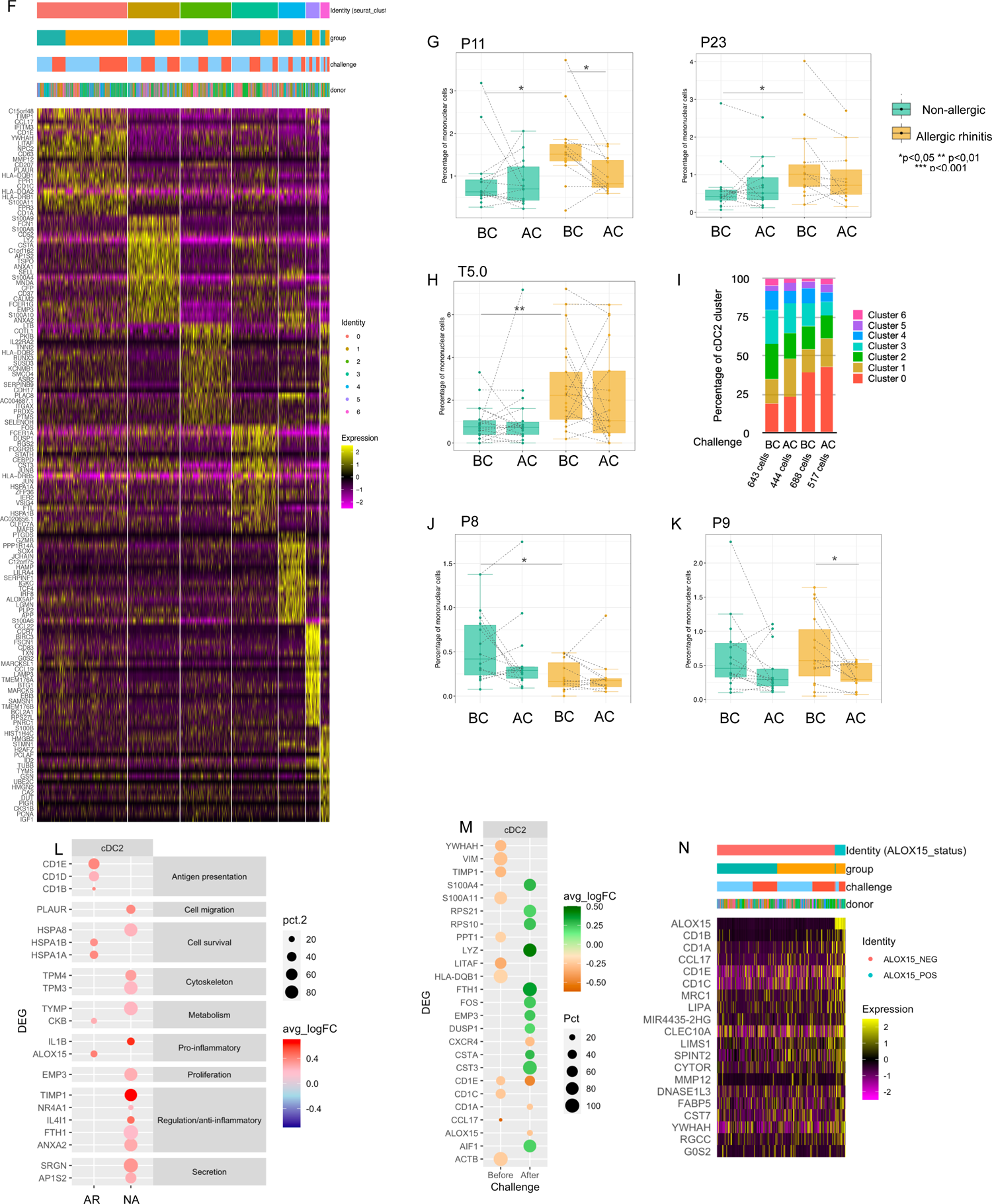
Differences in the balance between effector and anti-inflammatory cDC2 in allergic and non-allergic subjects A. UMAP of seven sub-clusters generated from cDC2, cluster T5 of the original clusters. Total cell number and distinguishing marker per sub-cluster depicted in the plot. B. UMAP of gene expression of established cell type markers. C. Dot plot of percentage of cells expressing marker genes of established cell types including DC2A, DC2B, DC3, AS DC, mature DC, Langerin+ DC and CD1a+ DC. D. Alignment of cDC2 cell clusters with those identified in proteomic data. E. Dot plot representing the top ten markers of each cluster sorted by log fold change for each cDC2 sub-cluster. F. Heat map of top 20 gene markers sorted by log fold change of the sub-clusters within the cDC2 compartment. Bars above heat map indicate composition of clusters based on group (allergic in orange; non-allergic in green), allergen challenge (blue before challenge; red after challenge) and donor. Colours of sub-clusters (identity) correspond with those depicted in UMAP in Figure 6A. G. Box plot of frequency of cDC2 clusters P11 and P23 of the proteomic data for in allergic (orange boxes) and non-allergic individuals (green boxes) before allergen challenge. * p < 0.05 H. Box plot of frequency of effector cDC2 (cluster T5, sub-cluster 0) of the transcriptomic data in allergic (orange boxes) and non-allergic individuals (green boxes) before (BC) and after allergen challenge (AC). ** p < 0.01 I. Composition of cDC2 by sub-cluster for non-allergic individuals (NA) and allergic individuals (AR), before (BC) and after allergen challenge (AC). Colours of sub-clusters correspond with those depicted in UMAP in Figure 6A. J. Box plot of frequency of cDC2 clusters P8 of the proteomic data in allergic (orange boxes) and non-allergic individuals (green boxes) before (BC) and after allergen challenge (AC). * p < 0.05 K. Box plots of frequency of macrophages (P9) in allergic (orange boxes) and non-allergic individuals (green boxes) before (BC) and after allergen challenge (AC) in the proteomic data. * p < 0.05 L. Dot plot of differentially expressed genes in cDC2 cells (Cluster T5) between before and after allergen challenge for allergic (AR) and non-allergic (NA) individuals. Red represents up-regulation of genes, blue represents down-regulation of genes. Colour intensity represent log fold change (with 0.2 cut off). Size of dot represents the percentage of cells genes are expressed in after allergen challenge. M. Dot plot of differentially expressed genes in cDC2 cells (Cluster T5) between allergic (AR) and non-allergic (NA) individuals, before (BC) and after allergen challenge (AC). Orange represents gene expression higher in allergic individuals, green represents gene expression higher in non-allergic individuals, colour intensity represent log fold change. Size of dot represents the percentage of cells in which the genes are expressed. N. Heat map of top 20 gene markers sorted by log fold change of ALOX15 expressing cells (green; top bar) compared to ALOX15 negative cells (red; top bar) within the cDC2 compartment. Bars above heat map further indicate composition of clusters based on group (allergic in orange; non-allergic in green), allergen challenge (blue before challenge; red after challenge) and donor.

Recently, Brown *et al*. (*32*) identified effector/inflammatory cDC2 termed cDC2B and anti-inflammatory cDC2 termed cDC2A. Sub-cluster T5.2 and T5.6 contain cells that correspond to anti-inflammatory cDC2A (*32*), based on CD1C, CCR6, LTB, RUNX3, IL22RA2 expression and absence/reduced expression of CLEC10A, PSAP and CD86 (**Figure 5B-C**). However, some of the typical cDC2A markers (*32*) were absent (CLEC4A, AREG and NR4A3). Sub-cluster T5.6 additionally expressed genes related to mitosis (**Figure S8**), representing a ‘mitotic cDC2A’ subset (*32*). The remaining cDC2 sub-clusters T5.0, T5.1 and T5.3 expressed cDC2B markers (CLEC10A, CD1C, FCER1A), but each expressed a distinct profile (**Figure 5E-F**). Sub-cluster T5.1 expressed monocyte-like genes (FCN1, SELL, S100A8, S100A9), corresponding to mo-DC or the inflammatory DC3 population described by Villani *et al.* (*21,33,34*). In contrast, sub-cluster T5.3 expressed higher levels of regulatory/inhibitory genes (FCER1A, FTL, FCGR2B, RGS2, VSIG4) (*8,35-40*) along with MAPkinase signaling genes (FOS, FOSB, JUN, JUNB, DUSP1) **(Figure 5E, S8, Table S5**), representing the ‘non-inflammatory’ FCGR2B cDC2 described by Villani *et al*. (*21*), but may also be related to cellular stress response (**Figure S8**). Sub-cluster T5.0 was distinguished by effector DC markers (NMES1 IFITM3, NPC2) (*32, 41*), and higher expression of genes related to cell activation, defence response, and antigen processing and presentation (**Figure S8**, **Table S5**). Sub-cluster T5.0 also contained CD1A-expressing cells, which is similar to cluster P11 and P23 in the proteomic data (**Figure 3B, S6**). In the lung, CD1a^+^ “pulmonary DC” are considered potent T cell stimulators (*2,42,43*). Allergic subjects had higher percentages of the CD1A cDC2 (cluster T5.0, P11) compared to non-allergic subjects already at baseline in both data sets (**Figure 5G-I**). In contrast, the percentage of cluster P8 (proteomic data), containing FceRI^low^ CCR6^+^ HLA-DR^low^ cDC2 cells and similar to the anti-inflammatory cDC2A sub-cluster T5.2 and T5.6 (transcriptomic data), was higher in non-allergic subjects compared to allergic subjects (**Figure 5J**). Following challenge, a decrease was detected in the percentage of CD1a^+^ cDC2 (cluster P11) and macrophages (cluster P9) in allergic subjects in the proteomic data, possibly reflecting migration of these cells (**Figure 5G, 5K**). In contrast, in non-allergic subjects, MF (cluster T18 – **Figure 2A**) expressed more Major Vault Protein (MVP) along with SRGN and LIMS1 **(Table S4**). MVP inhibits NF-kB signaling and attenuated murine allergic rhinitis (*44*), suggesting a role for MF in reducing or preventing allergic symptoms.

Sub-cluster T5.4 was identified as AS-DC or pre-DC (*21, 26*), based on AXL, DAB2, PPP1R14A and LILRA4 expression (**Figure 5B-C**) and aligned with cluster P17 (proteomic data), expressing CD123, CD5 and CD11c (**Figure 3B, S7, 5D**). Pre-DC are potent T cell stimulators and infiltrate the airways within 8 hours in response to LPS inhalation (*45*). Sub-cluster T5.5 was identified as a small mature DC population (106 cells; **Figure 6A,6C, S8, Table S5**), expressing CCR7, CD83, CD127 and LAMP3, resembling a CCR7 and IL7R (CD127) expressing DC subset (*46*). Genes related to immune regulation and Treg induction (SAMSM1, IL4I1, RELB, IDO1, IL7R) were among the top markers for this subset (**Figure 5E-F**). An equivalent CCR7^+^ DC cluster in the proteome dataset was not detected (**Figure 3B**), although CD127 expression was detected on a small portion of cluster P23 possibly pointing at the same population (**Figure S7**). The percentage of T5.5 and T5.4 within all mononuclear cells did not differ between groups, nor following allergen challenge.

Differential gene expression analysis of all cDC2 cells (cluster T5) showed upregulation of ALOX15, CD1B and CD1E in allergic subjects, but not non-allergic subjects, following allergen challenge (**Figure 5L-M**, **Table S4**). These genes link to CD1A cDC2 cells (sub-cluster T5.0). ALOX15 is induced by Th2 cytokines IL-4 and IL-13, suggesting local IL-4/IL-13 release in response to allergen challenge in allergic subjects. CD1 genes belong to the lipid antigen presentation pathway and are associated with DC maturation. Upregulation of these genes suggest increased capacity to present antigens to T cells following allergen encounter. CKB is primarily expressed in non-classical monocytes and might indicate the differentiation of these cells into (mo-)DC (*47*), following infiltration into the nasal tissue. Analysis of ALOX15-expressing cells revealed higher levels of CD1 (CD1A, CD1B, CD1D, CD1E), and IL-4/STAT6 inducible genes (CCL17, LIPA, FABP5, DNASE1L3, SPINT2) compared to ALOX15-negative cells within cluster T5 (**Figure 5N**). Interestingly, more genes were upregulated in cDC2 cells (T5) of non-allergic subjects compared to allergic subjects in response to allergen. These genes were related to ‘regulated exocytosis’, cell metabolism, cell survival and mediator secretion (**Figure 5L**). Furthermore, several genes have anti-inflammatory or regulatory functions, or expressed in tolerogenic DC (NR4A1, IL4I1, FTH1, ANXA2, TIMP1) (*48–54*), and several of these genes are also up-regulated in response to LPS (*50,55-57*).

Up to five distinct cDC2 clusters were identified in nasal tissue, of which several have been described by others including the anti- and pro-inflammatory cDC2A and cDC2B subsets, respectively. A predominance of effector CD1a^+^ cDC2 cells was found in allergic subjects while regulatory subtypes were overrepresented within non-allergic individuals. Pro-inflammatory cDC2 subsets were reduced in allergic subsets following challenge, presumably due to migration, whereas anti-inflammatory/regulatory gene expression in cDC2 clusters further increased after challenge in non-allergic individuals.

## DISCUSSION

Mucosal surfaces in the upper airways are constantly challenged by pathogens and foreign particles and while protecting against invaders, should also maintain immune homeostasis to protect against unnecessary and chronic inflammation. We performed a comprehensive single cell proteome and transcriptome analysis on nasal tissue biopsies, following controlled allergen challenge in allergic and non-allergic individuals, to identify cells that are involved in inflammation and tolerance.

While the known cell subsets such as ILC2, Th2 and eosinophils were associated with allergic patients, we discovered that additional cell clusters that largely segregated the allergic and non-allergic susbjects following allergen challenge belonged to MPS. Five sub-clusters were identified within CD14^+^ monocytes (transcriptomics), which could be grouped into HLA-DR^hi^ and HLA-DR^low^ populations. In non-allergic individuals, only HLA-DR^low^ CD14^+^ monocytes increased in frequency in nasal tissue in response to allergen, whereas in allergic subjects the frequency of HLA-DR^hi^ CD14^+^ monocytes and CD16^+^ monocytes significantly increased. CD14^+^ CD16^-^ and CD14^+^ CD16^+^ monocytes express high levels of HLA-DR are considered pro-inflammatory, and their recruitment has been implicated in various inflammatory diseases (*67, 68*). CD16^+^ CD14^-^ monocytes have been termed patrolling monocytes and are recruited to areas of inflammation to assist in tissue repair. They have been shown to differentiate into mo-DCs, with migratory abilities (*69*) that produce TNFa (*47*) or induce higher levels of IL-4 production from CD4^+^ T cells compared to CD16^-^ monocytes (*69–71*). The increased levels of CD16^+^ monocytes in allergic subjects following challenge will perpetuate the Th2 type response.

Interestingly, HLA-DR^low^ CD14^+^ monocyte clusters in non-allergic subjects expressed genes similar to myeloid-derived suppressor cells (MDSC), including the anti-microbial S100A8/9/12 genes (*58*). These cells accumulate in cancer and chronic inflammatory conditions (*59–61*) but can also be involved in maintaining immune homeostasis (*62*). The striking combination of anti-microbial and regulatory genes has also been found in mo-MF with immunoregulatory functions (*27*). Such cells are expanded through stimulation with butyrate (*63*), an immunoregulatory short chain fatty acid (SCFA) produced by microbiota. Butyrate can act distally in the (upper) airways, influencing local immune cells (*64, 65*). Of note, the population of HLA-DR^low^ monocytes in AR patients was higher at baseline, but did not change upon challenge, in contrast to the increased influx in non-allergic controls. This may align with a previous study (*66*) where increased numbers, but functionally impaired, CD14^+^ HLA-DR^low^ MDSC were found in peripheral blood of psoriasis patients.

In the cDC2 compartment we found different cDC2 clusters, of which the effector CD1a^+^ cDC2 subpopulation dominated in allergic subjects at baseline, and these cells responded to allergen challenge in allergic individuals. This population seems similar to the cDC2B subset, which has been defined as pro-inflammatory by Brown *et al*. (*32*). Furthermore, previous studies on nasal mucosa have found elevated numbers of mature CD1a^+^ DC in allergic rhinitis compared to non-allergic individuals (*72, 73*) and support our findings regards CD1a^+^ cDC2 cells identified here. CD1a^+^ cDC2 cells can rapidly activate pre-existing allergen-specific Th cells, as HLA genes are upregulated (**Table S5**). In contrast, in non-allergic individuals the various cDC2 subpopulations are more balanced, and include the anti-inflammatory cDC2A population and FCGR2B^+^ cDC2 cells (*21, 32*). cDC2A have been described as anti-inflammatory T-bet^+^ cDC2 (*32*). A recent study of migratory CD11b^+^ DC in mice revealed upregulation of T-bet, which was induced through HDM and LPS stimulation. This, in turn, promoted T-bet expression in T cells and prevented allergen-specific Th2 responses (*74*). It is yet unclear how this population (or cDC2A) respond to stimuli, however both anti-inflammatory and Th1 responses are beneficial in preventing/reducing Th2 responses towards allergens. Interestingly, as cDC2A and cDC2B cells likely acquire their specific transcriptional profiles through environmental cues (*32*), the inflammatory status of the tissue will influence the balance between the different cDC2 subtypes in allergic and non-allergic individuals.

FCGR2B cDC2 represent a separate cluster distinguished by higher expression of several anti-inflammatory/regulatory markers (FCER1A, FCGR2B, FTL, VSIG4), although expression of MAPkinase signalling genes (FOS, JUN) was also increased, suggesting responses to inflammatory cytokines or stress. However, this may also be induced during collagenase treatment of tissue biopsies (*75*). Nevertheless, their expression profile indicates a regulatory role relevant to maintain immune homeostasis. Furthermore, Villani *et al*. (*21*) made the distinction between ‘non-inflammatory’ FCGR2B^+^ DC2 and ‘inflammatory’ FCGR2B**^-^** CD163^+^ CD36^+^ DC3. Although those were analysed from blood, our tissue populations seem similar, and expression of some signature genes was enhanced in cDC2 cells from non-allergic individuals following allergen challenge.

A previous study with grass and tree pollen challenge showed a rapid influx of CD14^+^ HLA-DR^+^ monocytes in the nasal tissue of AR subjects. This was detectable 12 hours after allergen challenge (*16*), followed by recruitment of Th2 cells and eosinophils. Transcriptome analysis of CD45^+^HLA-DR^+^ cells within nasal biopsies after an 8-day allergen challenge revealed upregulation of genes regulated by IL-4 and or IL-13, including ALOX15, CD1A and CD1B, similar to our findings here following HDM challenge. The origin of these transcripts (monocytes, cDC or pDC) was not identified in that study due to low cell numbers, however we have now determined that these transcripts are derived from CD1A^+^ cDC2 cells (within sub-cluster T5.0 of cDC2 cells), expressing CCL17 and other IL-4-inducible genes. Furthermore, our data shows that this expression profile is already present in resident tissue cDC2, prior to allergen challenge. Several studies have shown this to emerge after 7-8 days of repeated challenge (*16,76,77*), whereas we show here that these transcripts are present at baseline and further increase compared to baseline.

In contrast to our findings in the transcriptome data, Gracia *et al.* (*16*) using flow cytometry did not detect an increase in monocytes of non-allergic subjects in response to pollen allergen challenge. However, this is consistent with our proteome data, suggesting that slight differences may exist in the cell cluster annotation between transcriptome and proteome data sets. Alternatively, this may be due to the composition and the type of the allergen extract used. We used a total HDM extract, possibly containing trace amounts of various microbial compounds but representing the allergen composition encountered in non-experimental, everyday settings. Inhalation of microbial compounds, such as LPS, has been shown to induce neutrophil, CD14^+^ CD16^-^ monocyte and CD1c^+^ DC recruitment to the airways in non-allergic individuals. Additionally, monocytes expressed lower levels of antigen presenting genes, higher levels of LPS-response genes (e.g. IL1A, IL1B), and retained S100A8/9 and SELL expression in comparison to steady state, which could be similar to infiltrating HLA-DR^low^ monocytes/MDSC we also observed. Furthermore, IL1B expression was increased in recruited CD1c^+^ DC, which we also observed in DCs of non-allergic individuals after allergen challenge (*45*).

Despite the large number of subjects included in this study, there are specific limitations. Cell numbers varied per sample and sometimes was too low to contribute to smaller (sub)clusters, affecting the identification of differentially expressed genes between groups or conditions. However, this limitation was partly compensated by the side-by-side comparison of two independent datasets, allowing for more confidence with respect to changes in (sub)clusters despite the low cell number counts. The transcriptomic data may be affected by the technical handling of cells (e.g. tissue digestion, cryopreservation, FACS sorting), but this was partly compensated by the distribution of samples over only five carefully assigned batches, reducing sample to sample variation. It was therefore reassuring that we were able to align and validate most of our cell types with those described in previous studies.

Collectively, our data indicate a distinct, local, innate immune response to allergen in non-allergic individuals, characterized by infiltration of HLA-DR^low^ CD14^+^ monocytes and transcriptional activation of cDC2, including upregulation of tolerogenic genes. The response in allergic individuals indicates a role for infiltrating HLA-DR^hi^ CD14^+^ monocytes, CD16^+^ monocytes, and ALOX15 upregulation in CD1A^+^ cDC2, in the development or maintenance of allergic responses. Future therapies should be tailored towards those innate populations, either enhancing or reducing their activity for the treatment of allergic airway disease.

## MATERIALS AND METHODS

### Study design and sample collection

Adult subjects sensitized to house dust mite (HDM) who fulfilled the ARIA criteria (*78*) for moderate to severe persistent allergic rhinitis and non-allergic subjects, without sensitization to inhalant allergens, and without clinical features of allergic rhinitis were included in the study. Allergic subjects had a history of perennial rhinitis symptoms with or without conjunctivitis and were skin prick test positive to HDM extract (Der. Pteronyssinus, ALK-Abello, Denmark) (skin wheal area ≥ 0.4 HEP, corresponding with the internationally accepted wheal diameter of ≥ 3 mm). Non-allergic subjects had no history of allergic rhinitis and were SPT negative to HDM, birch pollen, grass pollen, cat and dog or other animals with which they were in daily contact with. Exclusion criteria consisted of pregnancy, nasal polyps and anatomical or other disorders of the nose. In total 30 allergic subjects with and 27 non-allergic subjects were recruited (**Table 1**).

### Nasal Allergen Provocation

Nasal allergen provocations with HDM extract (ALK-Abello) were performed according to the protocol shown in Figure 1A, as described previously (*79–81*). Provocations were performed once a day on three consecutive days. Nasal corticosteroids and antihistamines were withdrawn 3 weeks and 3 days respectively before the provocation. Provocations were performed in the absence of total nasal obstruction or infection as assessed by rhinoscopy. The nasal provocation on the first day was performed with 3 increasing doses of allergen extract (100,1000,10000 BU/ml) into both nostrils at 10-minute intervals after sham challenge with PBS containing human serum albumin 0.03% and benzalkonium chloride 0.05%. (ALK Abelló). The nasal response was assessed with a score system according to Lebel (*19*). The second and third challenge into one nostril were performed with PBS and HDM 10000 BU/ml only. PBS and the allergen extract were sprayed into the nostrils with a nasal pump spray delivering a fixed dose of 0.125 ml solution. Nasal provocations were performed out of pollen season between September 2017 and February 2018.

### Nasal biopsies

Biopsies were taken twice: once before the allergen provocations from one nostril and once one day after the third allergen provocation from the other nostril. Three 2mm^2^ biopsy samples were taken from the inferior turbinate. Beforehand, the nasal mucosa was anaesthetized by inserting 3 pieces of cotton wool on the mucosa soaked in 5% cocaine hydrochloride that was inserted onto the mucosa. 15 Minutes were allowed for the local anaesthetic to take effect. Following this, biopsy samples were taken using a specially developed Gerritsma biopsy forceps (Phoenix Surgical Instruments Ltd, Hertfordshire, UK). Haemostasis was achieved by packing the nose with cotton wool balls soaked in 0.5mL of 1:1000 adrenalin. These were removed after 15 minutes and the nose was then examined for sites of bleeding, if present, areas were cauterised with a bipolar electrocoagulation (Erbe Surgical Systems, Marietta, Georgia, USA). The patient was observed for a further 15 minutes and cauterisation was performed again if necessary.

### Nasal biopsy digestion

At each visit, before and after allergen challenge, three individual nasal biopsies were taken and stored in 5mL of cold 10% FCS IMDM for no longer than 1 hour. The biopsies were then finely cut using a sterile scalpel and digested in 10% FCS IMDM with Liberase TL (125 μg/mL, Sigma-Aldrich, MO, USA) and DNAse I (100 μg/mL, Sigma-Aldrich) overnight at 4°C. After digestion, an equal volume of FCS was added and the suspension was vortexed for 30 seconds. The biopsies were then pressed through a 100μm filter, rinsed thoroughly with IMDM and filtered over a 70μm filter. Cells were spun down for 10 minutes at 400 x g followed by red blood cell lysis with an osmotic lysis buffer (eBioscience). Cells were then resuspended in RPMI 1640 and counted. From each sample, 1×10^5^ cells were removed and cryopreserved, as described in the PBMC isolation section, for single cell RNA sequencing at a later date. The remaining cells were spun down for 10 minutes at 400 x g and resuspended in staining buffer (Fluidigm, CA, USA) for mass cytometry staining with metal-conjugated antibodies.

### Mass Cytometry Measurements and Analysis

Antibody-metal conjugates **(Table S1**) were purchased from Fluidigm or conjugated using 100 μg of purified antibody and the Maxpar X8 Antibody Labelling kit (Fluidigm). Conjugated antibodies were stored at 4**°**C in Antibody Stabilizer PBS (Candor Bioscience, GmbH, Wangen, Germany). All antibodies were titrated prior to use. Cells were stained with metal conjugated antibodies for mass cytometry according to the Maxpar Surface Staining protocol V2. Briefly, cells were stained with 1 μM intercalator Rh-103 (Fluidigm) for 15 minutes, washed and incubated for 10 minutes with TruStain FcX-receptor block (Biolegend) prior to a 45-minute incubation period with the antibody cocktail **(Table S1**). Cells were washed twice with staining buffer and incubated for one hour with 1 mL of 1000x diluted 125 μM Cell-ID intercalator-Ir191/Ir193 (Fluidigm) to stain DNA for cell identification. Cells were then washed three times with staining buffer and twice with deionized H2O. Finally, EQ Four Element Calibration Beads were added for normalization and cells were acquired on a Helios 2 mass cytometer (DVS Sciences). In addition to the metals included in the panel, channels to detect intercalators (103Rh, 193IR, 193IR), calibration beads (140Ce, 151Eu, 153Eu, 165Ho and 175Lu) and contamination (133Cs, 138Ba, 206Pb) were included during measuring. After data acquisition, the files were normalized with the reference EQ passport P13H2302 and where applicable concatenated. The median biopsy yield measure by mass-cytometry was 6,76 x 10^4^ cells (IQR: 2,46 x 10^4^ to 9,72 x 10^4^) per sample, of which 80,6% (median) were stromal cells. Viable, single CD45+ cells were pre-gated according to previously a described gating strategy (*82*) (**Figure S9**) and exported as new FCS files with Flowjo V10 for Mac (FlowJo LLC, Ashland, OR, USA). Data was transformed with hyperbolic arcsinh using a cofactor of 5 and distinct cell clusters were identified with Hierarchical Stochastic Neighbor Embedding (HSNE) in Cytosplore (https://www.cytosplore.org/) (**Figure S1**). Clustering was performed in two levels, once to determine the lineages present and another on the identified lineages. The second level of clustering on the MPS lineage was performed without the FceRI marker, as inclusion of this marker lead to additional clusters solely based on FceRI expression, which differed significantly between the non-allergic and allergic cohort (**Figure S3A).** The original MPS cluster 11 could be further split into two populations (cluster 11 and 23) which differed in CD206 expression **(Figure 3B**). Cytosplore output was analysed with the ‘Cytofast’ package (rdrr.io/github/KoenAStam/cytofast) (*92*) in RStudio (Rstudio, Inc., Boston, MA, USA, www.rstudio.com), to produce heatmaps, scatterplots of subset abundance and histograms of the median signal intensity distribution of markers. Cell numbers were normalized to the total number of CD45+ cells or mononuclear cells (CD45+ cells with granulocyte numbers removed, in order to compare to findings from the transcriptomic data) for each subject to correct for varying biopsy yields.

### Single cell RNA sequencing

Cryopreserved cells from digested nasal biopsies were thawed in pre-warmed 10% FCS RPMI 1640 and spun down at 300 x g for 10 minutes. Cells were resuspended in Cell Staining Buffer (Biolegend) containing TruStain FcX-receptor block (Biolegend) and incubated for 10 minutes on ice. TotalSeq cell hashing antibodies were then added following manufacturers recommendations, along with anti-CD45-APC (HI30, Biolegend). After a 15 minute incubation period on ice, the cells were washed with staining buffer and spun down at 300 x g for 10 minutes at 4**°**C. Supernatant was removed, cells were resuspended in staining buffer, and all samples were combined before adding 7AAD for exclusion of dead cells. Live, single CD45+ cells were sorted on a FACSAriaIII (BD Biosciences) into eppendorfs containing 0,04% BSA PBS. Cells were delivered to the Leiden Genome Technology Centre for single cell sequencing on the 10X Genomics Chromium system. A maximum of 30000 cells, from 15 barcoded samples, were encapsulated in each individual run, for a total of 5 runs (**Figure S10**). Cells were loaded according to the standard protocol of the Chromium single cell 3’ kit at a maximum concentration of 1000 cells/μl. Sequencing was performed on two lanes of an Illumina Hiseq 4000 to obtain coverage of at least 30000 reads/cell.

### Transcriptomic data mapping cells to subjects

Each barcode log-counts distribution over all cells of a single run followed a bimodal shape with well separable peaks. The high-count peak was interpreted as the signal for a particular barcoded donor. For each sample a two-component gaussian mixture model was fitted and a cell was assigned to a donor when it was at least 3 times more likely to be explained by the donor’s high-count peak than by the donor’s noise low-count peak (**Figure S11).** The majority of the cells were assigned to a single donor only (**Figure S12**). Cells that could not be assigned to a single donor were excluded from further analyses.

The barcode-based assignment was validated by investigating concordance between genotypes of samples assigned to the same donor but sequenced in different runs. In the pileup of pooled, aligned reads from all runs, genomic positions (SNPs) likely to differ between donors were identified. At each such position and separately in each cell we generated the count of transcripts with the reference base and count of transcripts with the alternative base(s). Cumulated positional counts of cells were assigned to the same sample and sample genotypes were called. Concordances were calculated for all sample pairs, and clear separation was observed for concordances expected to belong to the same donor vs. different donors (**Figure S13 and S14**). Positional counts of cells assigned to the same donor across multiple runs were cumulated and genotypes of all donors were called. A genotype-based donor assignment was then performed, by identifying the most likely genotype for each cell. Cells that were consistently assigned to the same donor by barcode-based and genotype-based methods were selected for further analysis (**Figure S15**).

### Single cell RNA sequencing analysis

The five demultiplexed gene expression matrices were imported in R package Seurat v3, retaining only genes expressed in at least three cells, and merged into a combined Seurat object. Cells with less than 200 or more than 4000 UMI or more than 25% of mitochondrial RNAs were filtered out. After filtering a total of 46238 cells were left for subsequent analysis. In order to identify shared clusters of cells collected from different treatments (before and after challenge) and groups (patient and control), the gene-barcode matrix was integrated according to the integration workflow (83) using SCT-transform to normalize and scale the gene-expression and reciprocal PCA (rPCA) instead of CCA, for dimensionality reduction (with 30 principal components). SCT normalised data was then used for further analysis of generated clusters.

Next, principle components analysis (PCA) and UMAP was performed and 30 PCs were used as input for graph-based clustering (resolution = 0.5). In order to assign the resulting 26 clusters to a cell type, differential gene expression was performed using the Wilcox test (FindAllMarkers function, logfc.threshold = 0.25, only.pos = TRUE, min.diff.pct = 0.2, assay = “SCT”). Top 10 gene-markers of each cluster were displayed with the DotPlot function. The number of cells per cluster in each condition was also calculated and plotted using UMAP projection and ‘split.by’ options in Seurat.

Cluster 4, 5,13,15,18,22 and 23 showed a typical MPS-like signature and were subset from the rest. To assign them more precisely to canonical MPS subgroups average gene expression of each cluster was calculated and plotted with the pheatmap package, along with a collection of canonical markers from the literature. To have an unbiased approach we also iterated the FindAllMarkers function on this specific subgroup (FindAllMarkers function, logfc.threshold = 0.25, min.diff.pct = 0.2, assay = “SCT”) FindAllMarkers compares each cluster to all the others, so in this case a more specific comparison to MPS only clusters will be performed. Top gene-markers per cluster were generated, and sorted by fold change values in descending order. Dot plots and heat maps (DoMultiBarHeatmap function, https://gitcrcm.marseille.inserm.fr/herault/scHSC_herault/blob/ecf93aa2d914d0b3bd508d066_ca887717d78b771/R_src/<u>DoMultipleBarHeatmap.R) were then generated from these lists with the Seurat R package.</u>

Cluster 4 and 5 were shown to contain multiple sub-clusters and were therefore individually subset and re-clustered: data was first re-scaled using the SCTtransform function and dimensionality reduction and clustering (res=0.5) was performed as in the general workflow. Differentially expressed genes between sub-clusters were then identified with the FindAllMarkers function without a min.diff.pct threshold, to include genes that are expressed in multiple sub-clusters but at different levels. ALOX15 positive cells (ALOX15_POS) within the cDC2 cell cluster (T5) were subset from ALOX15 negative cells (ALOX15_NEG) with the WhichCells function and a metadata field based on ALOX15 expression was added. Differentially expressed genes between the two subsets were determined with the FindAllMarkers function.

Each MPS cluster was analyzed to identify differentially expressed genes between conditions in each cell type (cluster). A combined metadata field was added, containing the combination of the cluster, the group (patient/control) and the treatment (before/after challenge) information. In this way the FindMarkers function in Seurat allowed the identification of DEG genes between 2 different subgroups of the same cell type (e.g ‘control_Before_4’ and ‘control_After_4’ indicate a comparison between the control group before and after challenge for cluster 4). The test used for analysis between before and after challenge was the LR (linear regression method) that allowed the inclusion of donor information as a latent variable, to correct for an uneven donor distribution in some of the clusters between timepoints. For this analysis we used the RNA assay in the FindMarkers function.

### Gene Ontology Enrichment Analysis

Enrichr (<u>amp.pharm.mssm.edu/lib/chea.jsp</u>) (84) gene ontology enrichment analysis was performed on the top 15 (as sorted by absolute log2 fold change) significantly differentially expressed genes (adjusted p-value <0.05) of each major cluster within the MPS compartment (4, 5, 13, 15, 18, 22, 23) to determine the association with specific cell types, based on annotations in the Human Cell Atlas and ARCHS4 tissue databases. The terms with the highest combined score (calculated by multiplying the log of the p-value of the Fishers exact test by the z-score of the deviation from the expected rank) were selected for visualization.

In order to phenotype/identify the individual cell clusters generated by sub-clustering the cDC2 (cluster 5) and CD14^+^ monocyte (cluster 4) populations, Gene Ontology (GO) enrichment analysis of biological processes (BP) protein complex database (CORUM) and Reactome pathways was performed with g:profiler (85) on the top 40 (as sorted by absolute log2 fold change) significantly differentially expressed genes (adjusted p-value <0.05) between sub-clusters. Genes were analyzed as an unordered query using g:profiler’s default g:SCS threshold based methods to correct for multiple testing, and all annotated genes detected in the transcriptomic data as a background set. Lists of enriched Gene Ontology categories were collapsed and simplified with Revigo (http://revigo.irb.hr) (86) using the Homo sapiens set of GO terms. GO terms with dispensability values less than or equal to 0.5 were then visualized in a dot plot (ggplot function, RStudio version 4.0), depicting the negative adjusted Log10 p value and the gene ratio, which equals the number of differentially expressed genes against the number of genes associated with the GO term.

## Statistical Analysis

Analysis of proteomic lymphocyte and granulocyte cell frequency data was performed with two-tailed, Wilcoxon matched-pairs signed rank tests, or Mann-Whitney test with a 95% confidence interval (Prism, GraphPad Software).

The normalized frequencies of the MPS cell clusters in both the transcriptomics and proteomic data were compared between non-allergic and allergic groups and between before and after allergen challenge. For this analysis a generalized linear mixed model was fitted with an underlying binomial distribution. For each cluster a separate model was fitted for the control group and the allergic group. The challenge was modeled as fixed effect. Lastly, volunteer ID was included as a random effect and a sample specific random effect was included to deal with the overdispersion. To control the FDR, p-values were corrected based on the Benjamini-Hochberg method, with p<0.05 considered significant. *p < 0.05; **p < 0.01;***p<0.001. Median and interquartile are range shown in the bar graphs.

## Study Approval

All subjects gave written informed consent and research was conducted in compliance with all relevant ethical regulations. Ethical approval was given by Erasmus Medical Centre Ethics Committee in Rotterdam (MEC2016-560).

## Author contributions

Conceptualization: ALV, MY, ECJ, RGW, HHS

Methodology: ALV, TGK, TT

Investigation: ALV, MLG, ED

Formal analysis: ALV, RM, SMK, KAS

Data curation: RM, SMK

Visualization: ALV, RM, KAS

Funding acquisition: HHS, RGW, ECJ

Project administration: HHS, RGW

Resources: SLK, RWH

Supervision: HHS, RGW, ECJ, MY

Writing – original draft: ALV, HHS

Writing – review & editing: ALV, MLG, RM, ED, TGK, TT, SMK, KAS, SLK, NWJ, RWH, MY, ECJ, RGW, HHS

## Data Availability

all data will be made available upon reasonable requests.

## Acknowledgments

We received a consortium grant from Netherlands Lung Foundation (5.1.15.015) and from the Netherlands Science Counsil (ZonMW) (40-43500-98-4066). We thank staff from the flow core facility for help with FACS sorting of cells from nasal biopsies prior to single cell transcriptomics.

## Supplementary Materials

**Figure S1.**
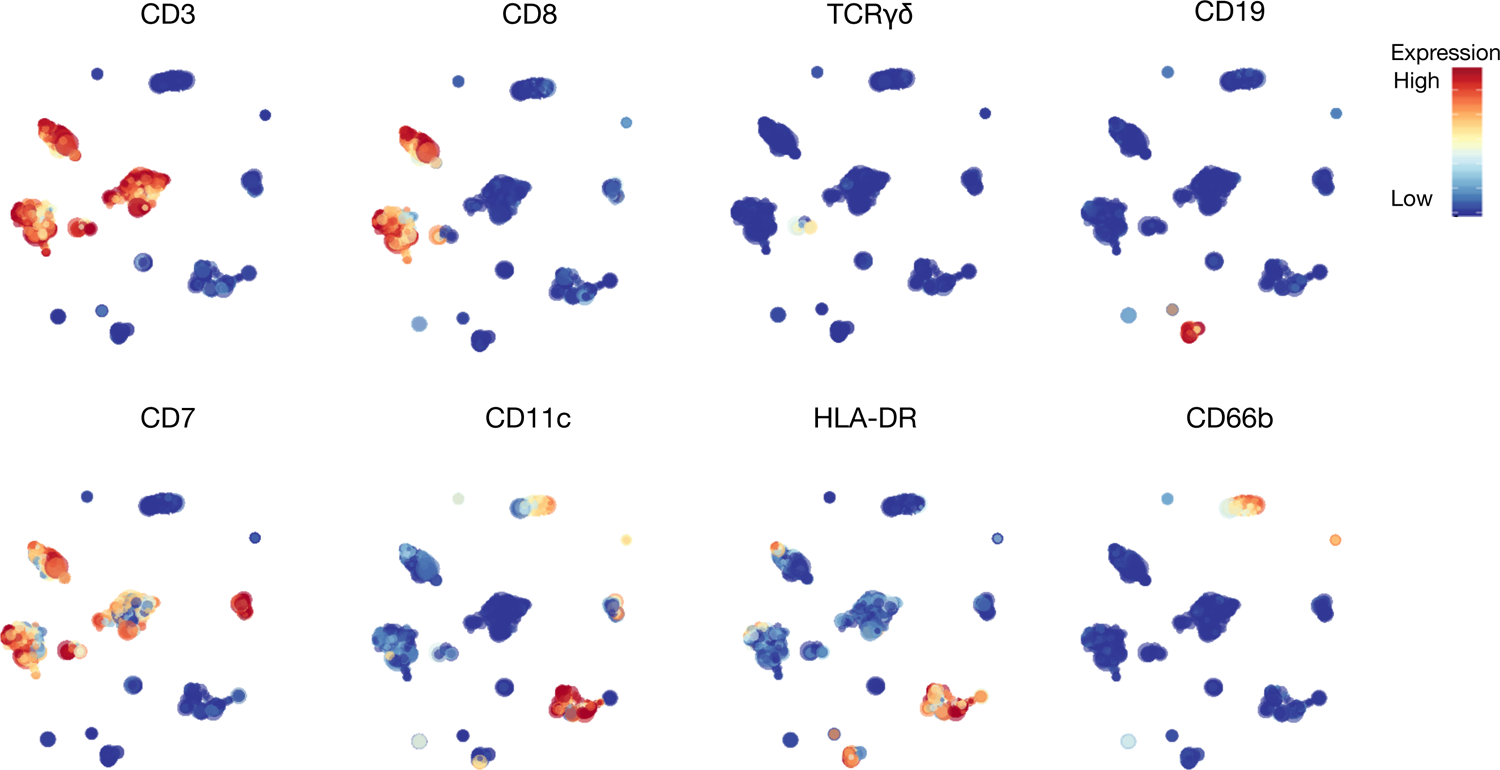
Cytosplore clustering of immune cells. Automated clustering was performed by hierarchical stochastic neighbour embedding (h-SNE) using Cytosplore software. Cell clusters were identified based on known markers of major immune cell lineages.

**Figure S2.**
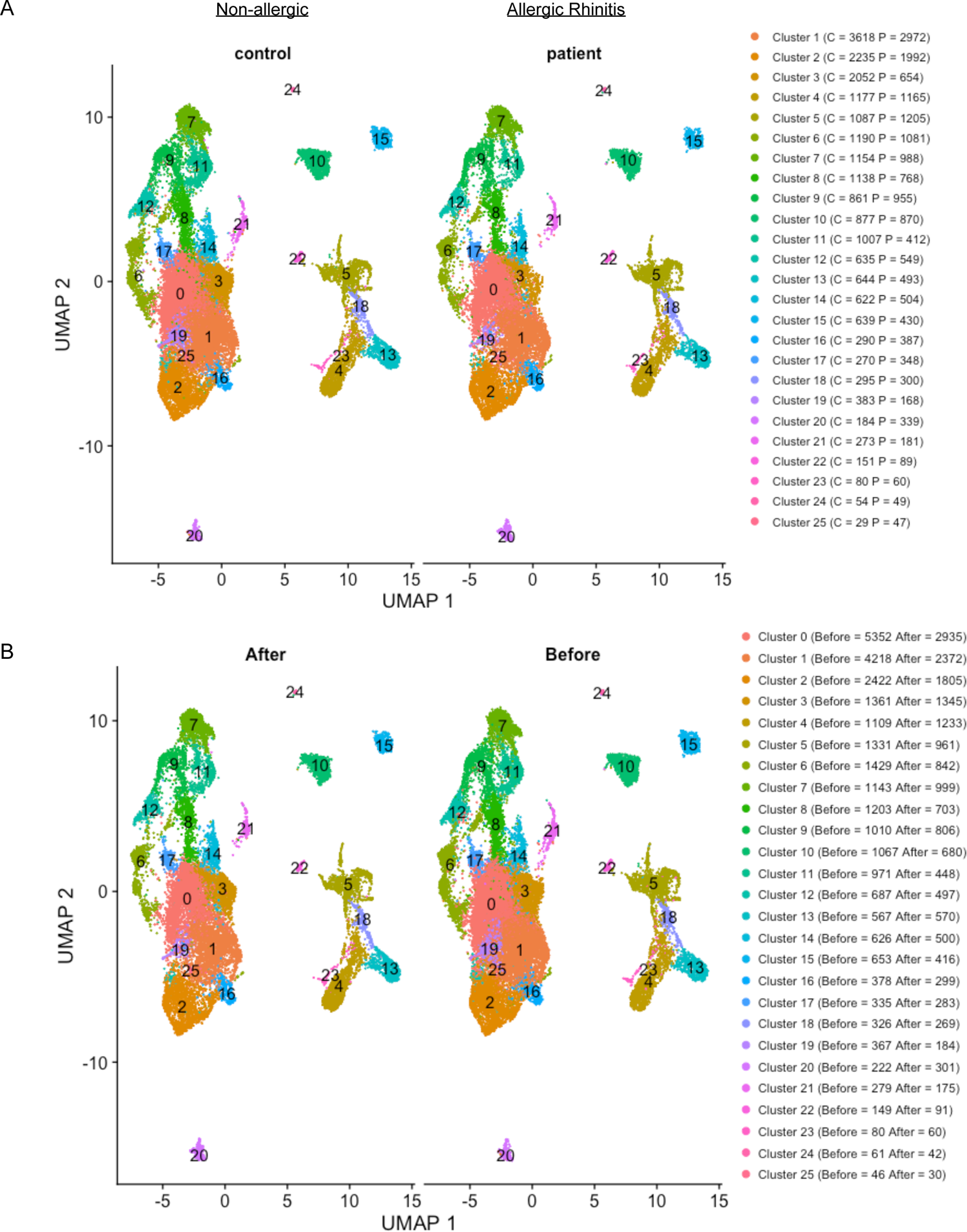
Single cell sequencing clusters and cell numbers UMAP of all cells measured by single cell RNA sequencing. Individual clusters are represented by colour, number of cells from each group (C, non-allergic controls; P, Allergic rhinitis patients) shown in A, number of cells before and after allergen challenge shown in B.

**Figure S3.**
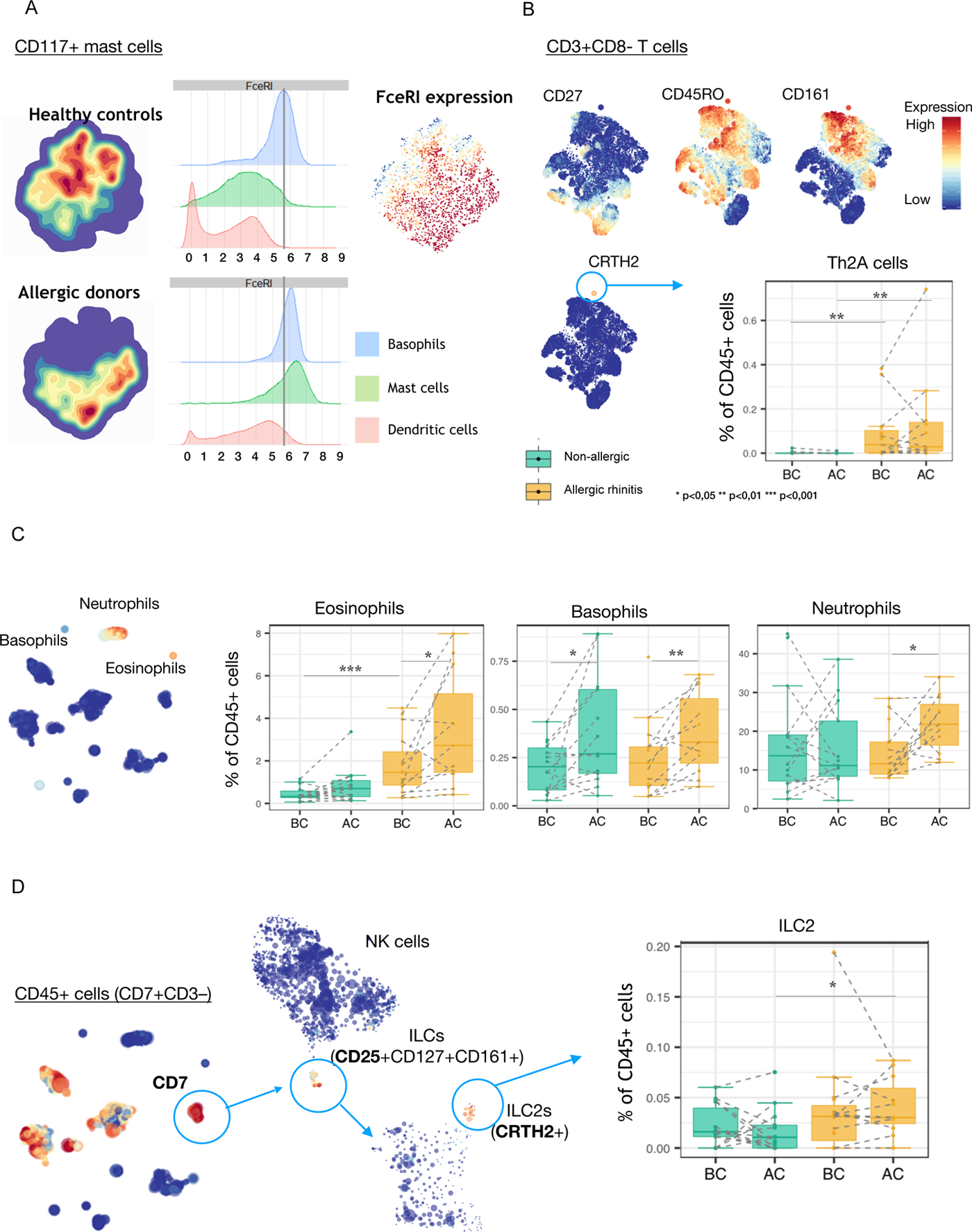
Evidence of type TH2 phenotype and response to allergen challenge A. In the mass-cytometry data analysis, clustering of the mast cell lineage resulted in a division of the population by group (non-allergic or allergic rhinitis) due to high FceRI expression on mast cells associated with allergic disease. Increased FceRI expression in allergic rhinitis compared to non-allergic subjects was most prominent on mast cells, but was also visible on basophils and dendritic cells. B. Pro-inflammatory CD4+ TH2A cells are characterised by CD45RO-CD27-CD49b+ CD161+ CRTH2+ and have previously been shown to represent allergen-specific, Th2 type cytokine producing T cells (ref 2,3). In allergic rhinitis subjects, nasal tissue Th2A cells were detected, expressing CD3+ CD8-CD45RO-CD27-CD161+ CRTH2+ whereas these cells were not present in non-allergic subjects. The prevalence of these cells did not change after allergen challenge in either group. C. A well defined characteristic of allergic inflammation is the infiltration of eosinophils into the site of allergen exposure. At baseline, allergic rhinitis subjects showed a higher prevalence of eosinophils in nasal tissue compared to non-allergic subjects, which increased significantly upon allergen challenge. In addition, an infiltration of neutrophils and basophils was also observed after challenge in allergic rhinitis subjects. In non-allergic subjects, an increase in basophils, but not eosinophils or neutrophils, was observed in response to the allergen challenge. D. The presence of ILC2 was also observed, albeit at very low levels (less than 0.05% of all CD45+ immune cells), in both groups. ILC2 were identified through 3 levels of cell clustering and defined as CD7+(CD3-) CD25+ CD127+ CD161+ CRTH2+. Although the prevalence of these cells between groups did not differ at baseline, there was a significant difference between the groups after allergen challenge, which could be attributed to a small (non-significant) increase in allergic rhinitis subjects, and a concurrent small (non-significant) decrease in the non-allergic subjects.

**Figure S4.**
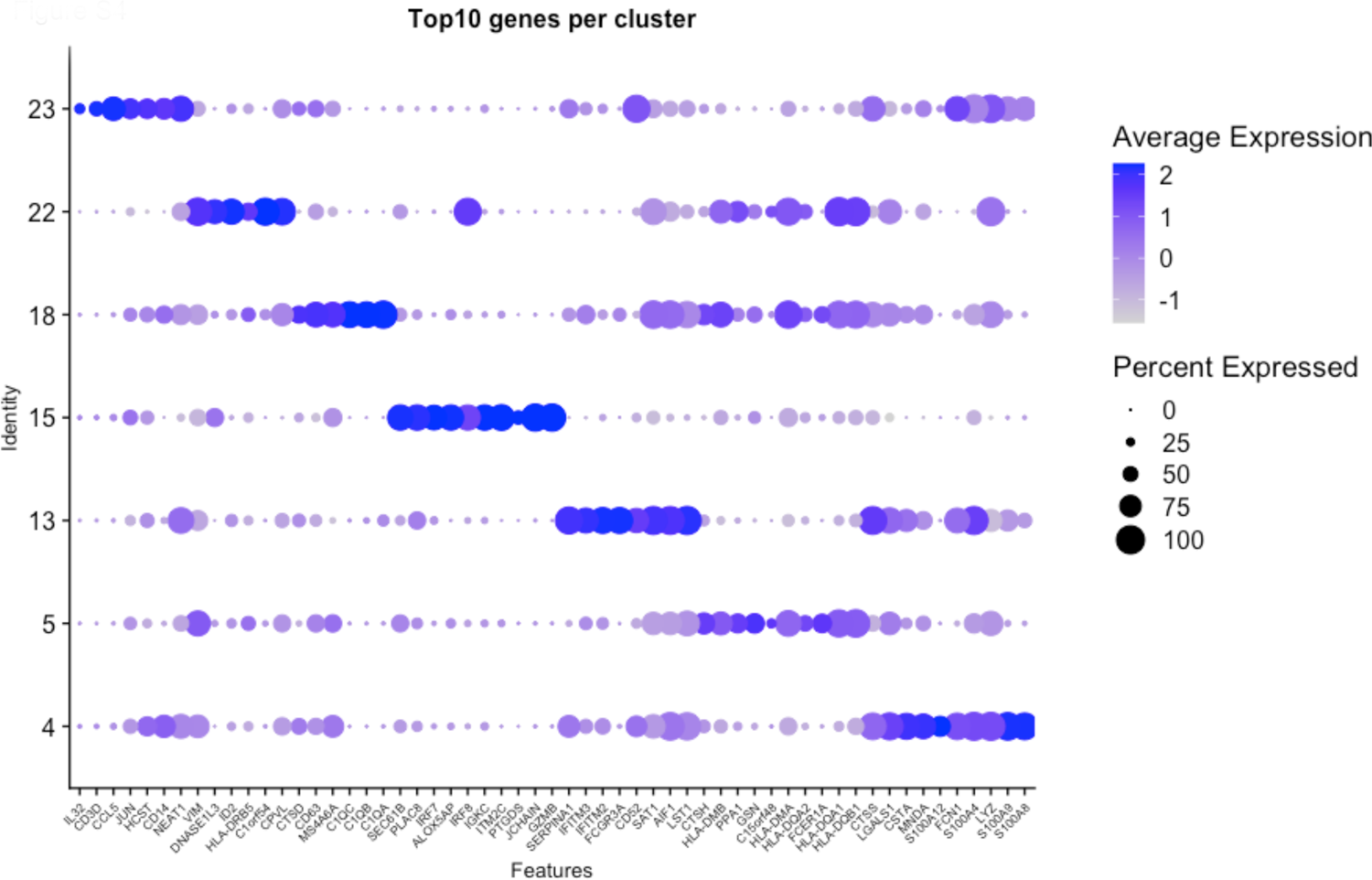
Top genes expressed by MPS clusters Top 10 genes, sorted by log fold change, expressed by each cluster of MPS cells, whereby average expression and percentage of cells expressing the gene are shown.

**Figure S5.**
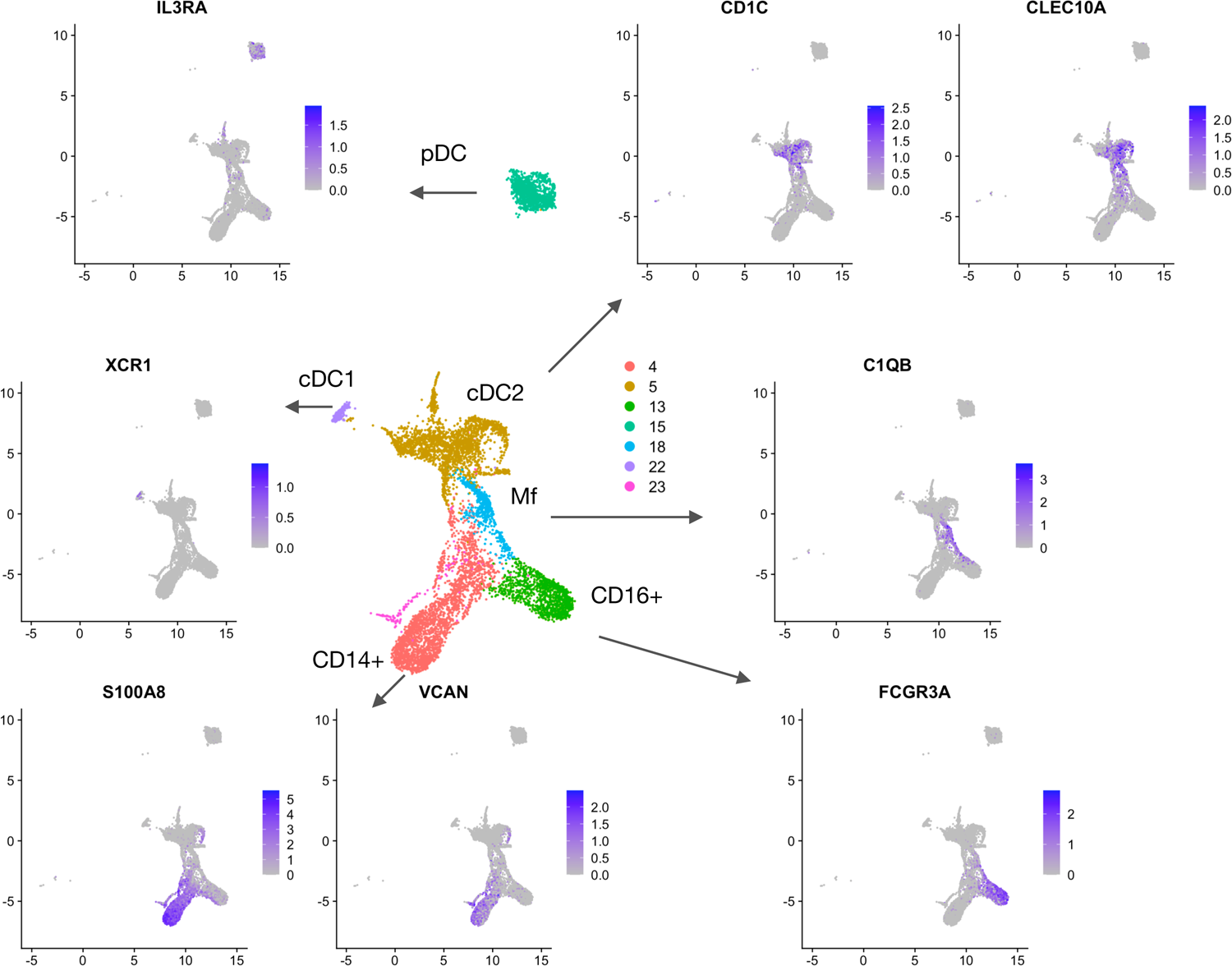
Annotation of MPS clusters by established markers UMAP of MPS cells depicting average expression of known myeloid cell type markers including IL3RA (pDC),XCR1 (cDC1), C1QB (macrophages), CLEC10A, CD1C (cDC2), VCAN, S100A8, and FCGR3A (monocytes).

**Figure S6.**
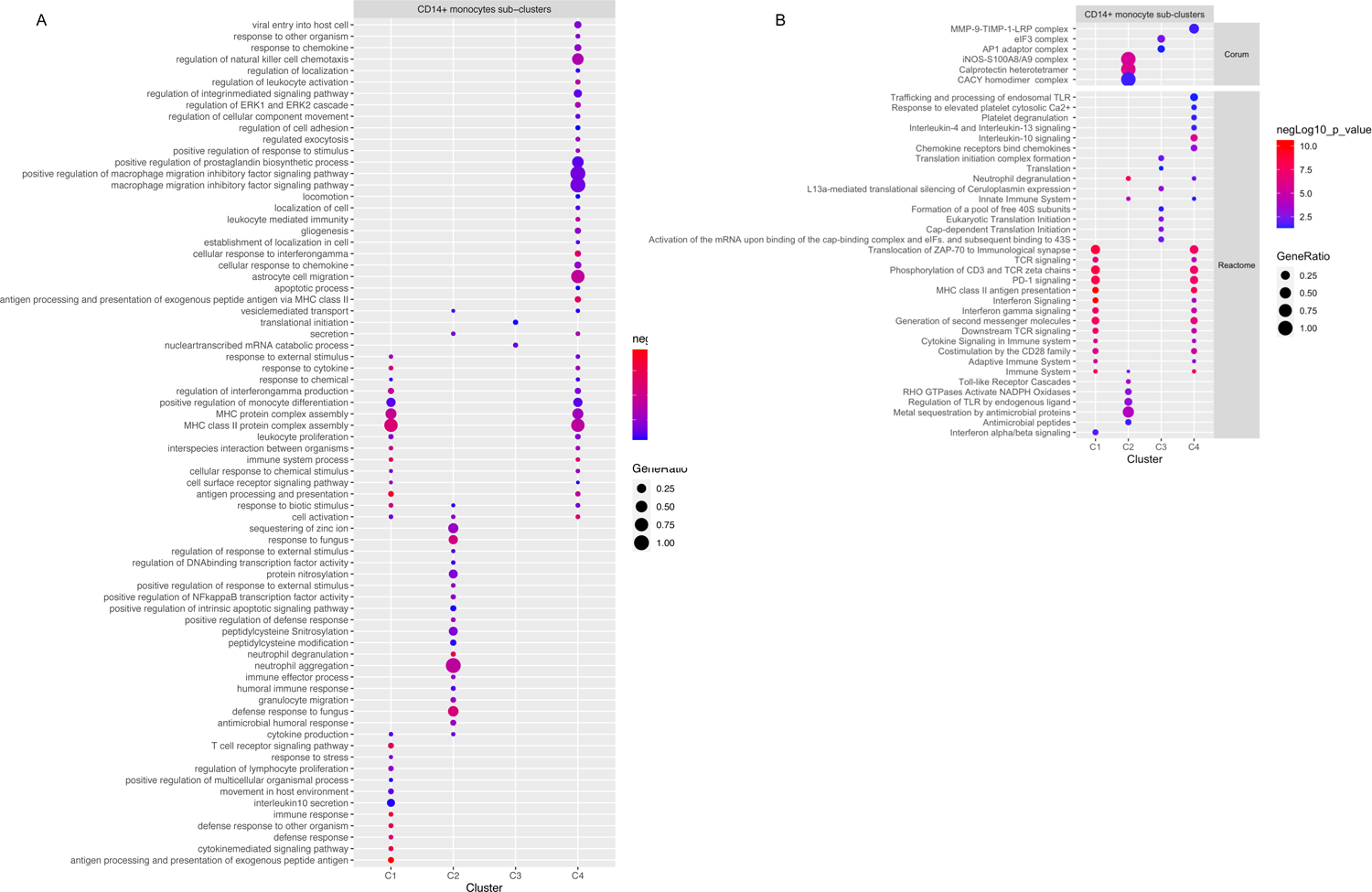
Gene Ontology terms corresponding with CD14+ monocyte sub-cluster gene expression Dot plot of Gene Ontology terms of biological processes (GO:BP) (A) and Reactome/Corum (B) generated with the top 40 (as sorted by absolute log2 fold change) significantly differentially expressed genes (adjusted p-value <0.05) between CD14+ monocyte sub-clusters in g:Profiler, and simplified with Revigo (GO:BP only). Colour of dots represent the negative Log10 of the adjusted p-value and the size represents the Gene Ratio, equaling the number of differentially expressed genes against the number of genes associated with the GO term.

**Figure S7.**
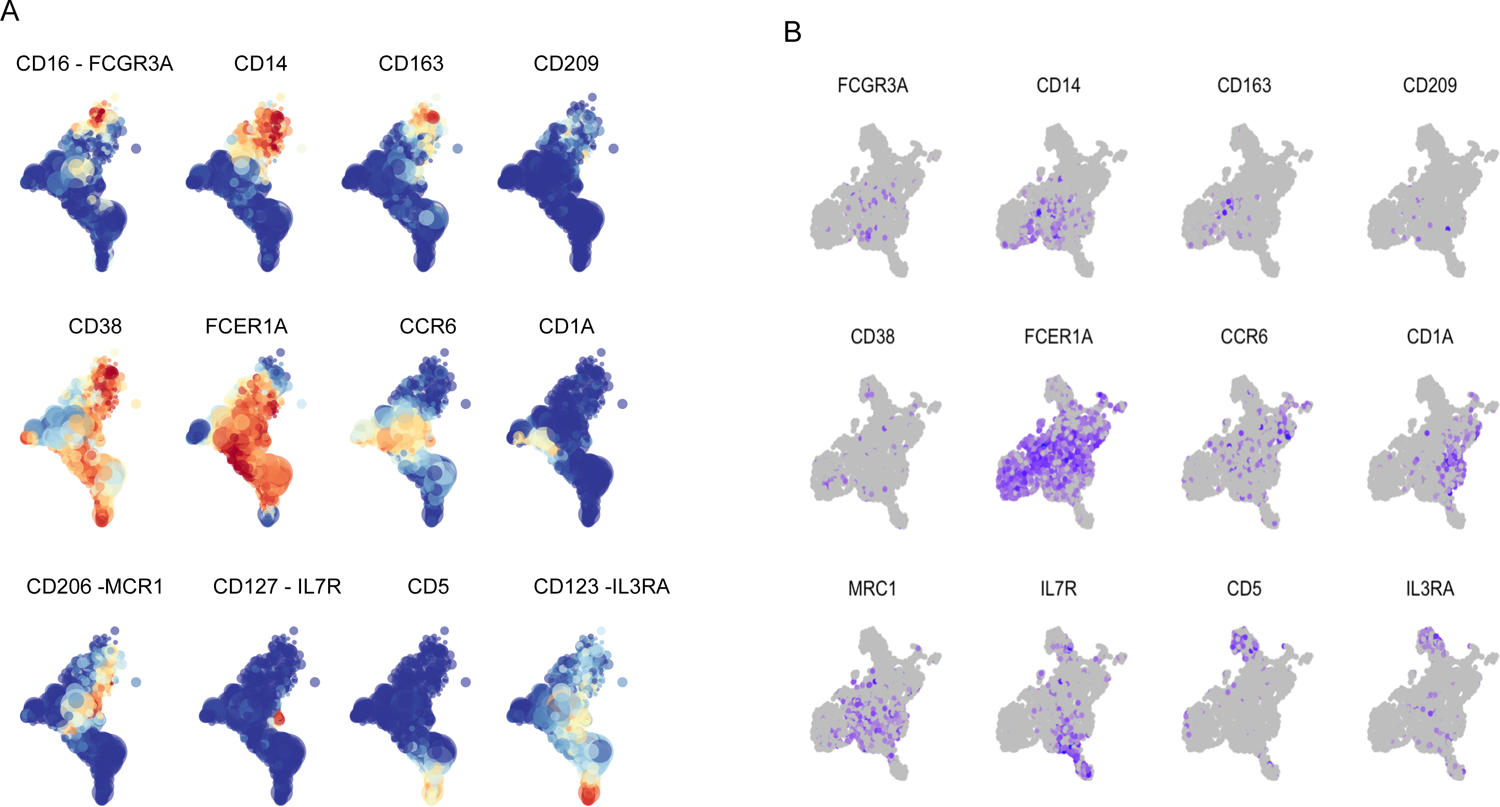
Alignment of transcriptomic and proteomic cDC2 clusters Alignment of cDC2 clusters between the proteomic (A) and transcriptomic (B) data by overlapping markers. In the proteomic data, CD16+ CD14+ CD163+ macrophages (cluster 9), CD14+ DC (cluster 16), CD14+ CD206+ DC (cluster 12), CD14-CD206+ DC, CD5+ CD123+ pre-DC (cluster 17) and CCR6+/CD1a+ cDC2 (cluster 8,11) are identified. In the transcriptomic data, CD5 IL3RA pre-DC (cluster T5.4), CCR6 CD1A cDC2 (within cluster T5.0, T5.2, T5.6) and CD14+ DC (within cluster T5.3) could be identified. While CD14+ or CD206+ (MCR1) cells cluster separately in the proteomic data (cluster 12, 16) these cells are distributed between cluster T5.0, T5.1 and T5.3 in the transcriptomic data.

**Figure S8.**
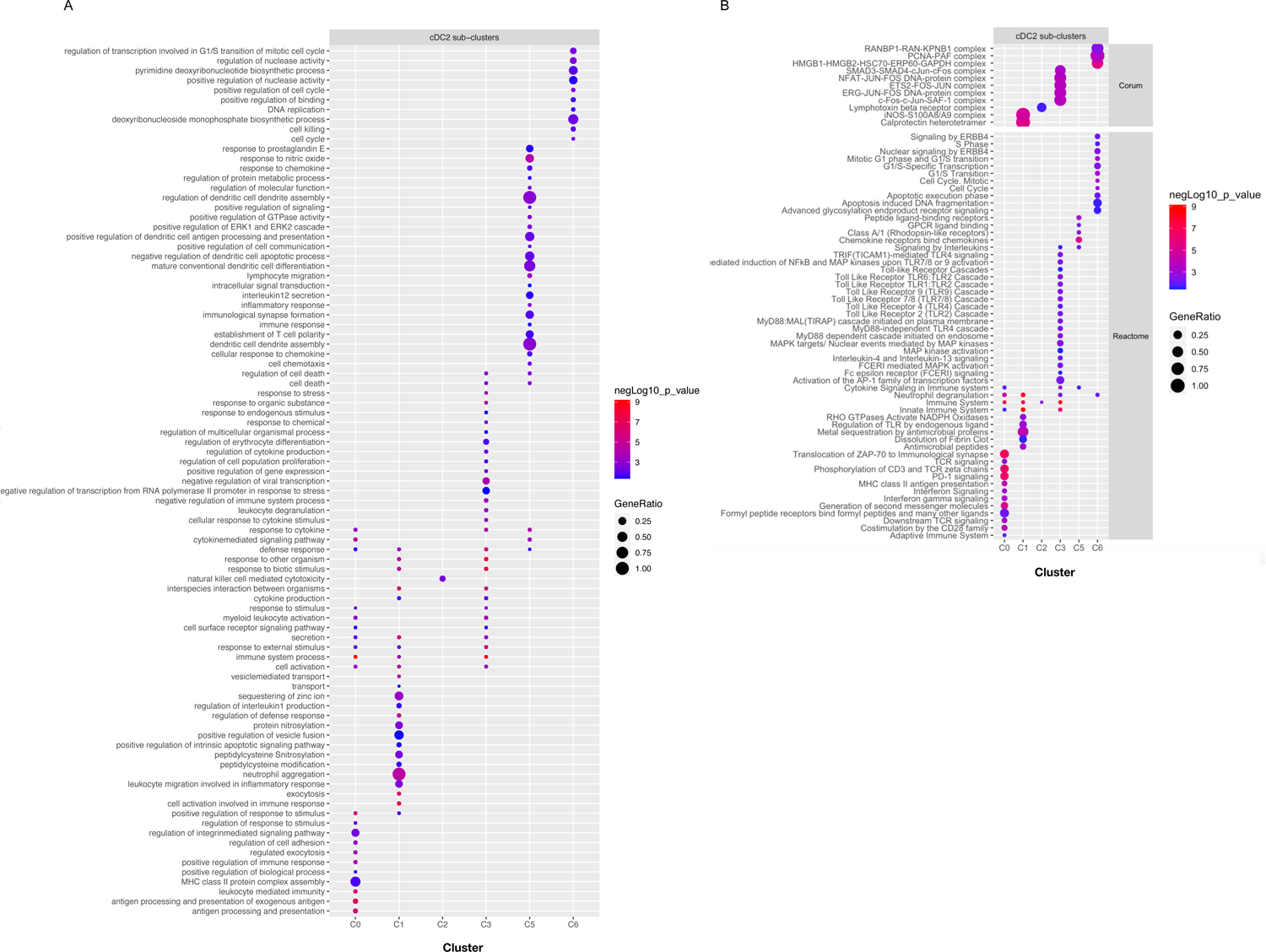
Gene Ontology terms corresponding with cDC2 sub-cluster gene expression Dot plot of Gene Ontology terms of biological processes (GO:BP) (A) and Reactome/Corum (B) generated with the top 40 (as sorted by absolute log2 fold change) significantly differentially expressed genes (adjusted p-value <0.05) between cDC2 sub-clusters in g:Profiler, and simplified with Revigo (GO:BP only). Colour of dots represent the negative Log10 of the adjusted p-value and the size represents the Gene Ratio, equaling the number of differentially expressed genes against the number of genes associated with the GO term.

**Figure S9.**
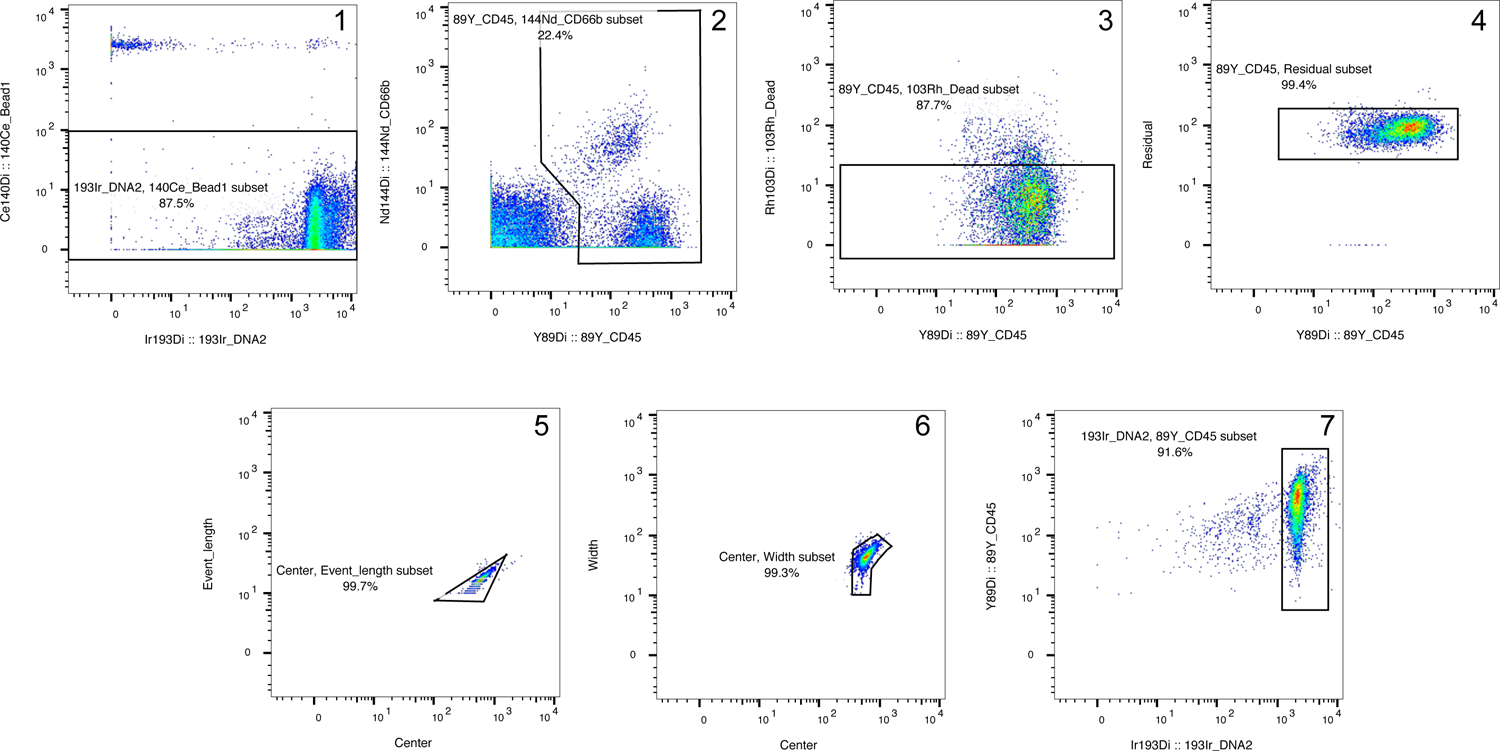
FCS data gating strategy Beads were excluded from analysis (1), CD45+ cells were gated (2), 103Rh+ cells were removed to retain only viable cells (3), residual, centre, event length, width and 193Ir DNA parameters were then used to obtain single cells according to established methods (4–6).

**Figure S10.**
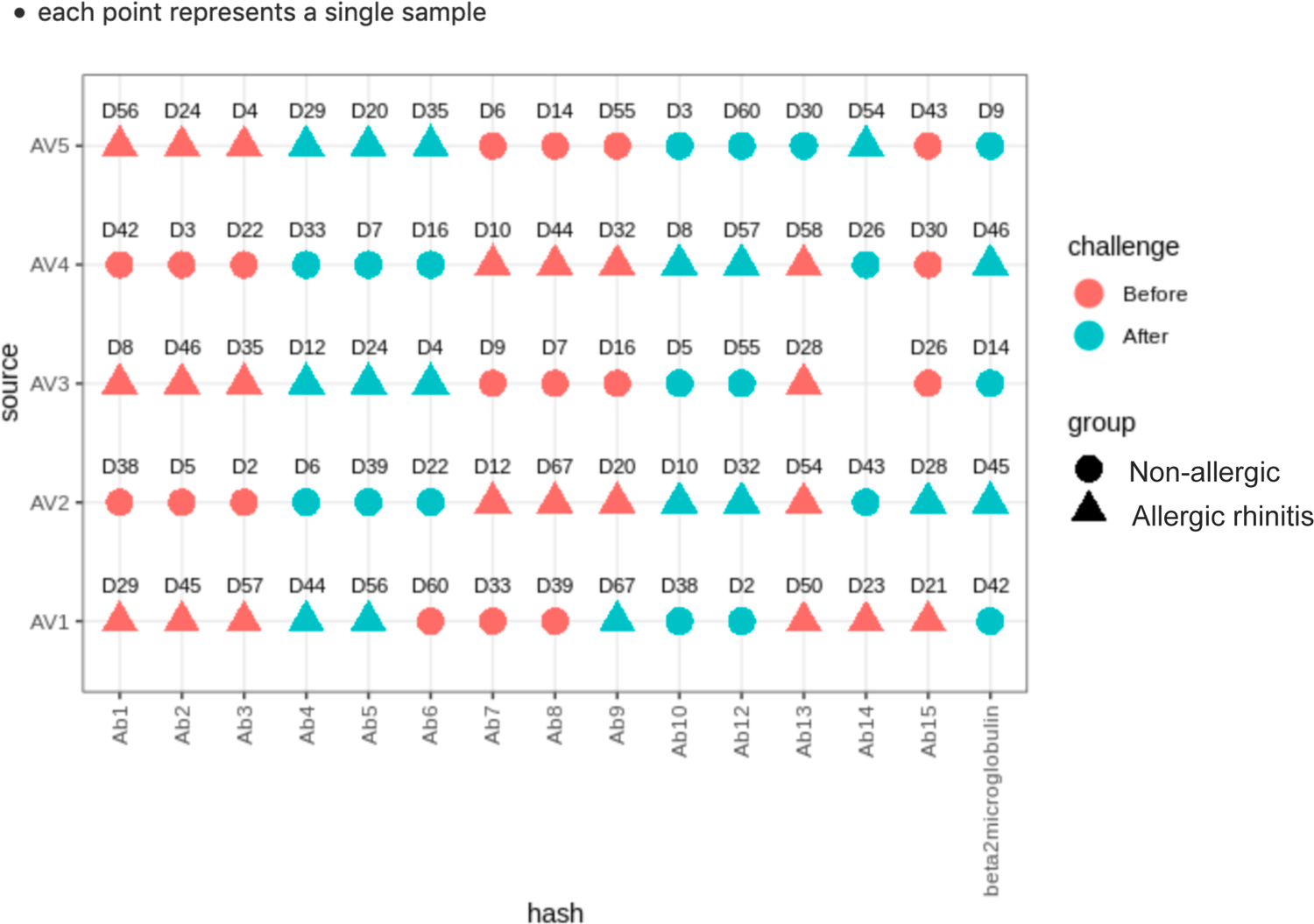
Experimental design for transcriptomic data-set generation All samples of the transcriptomic data, with donor number, group (allergic or non-allergic), challenge timepoint (before or after), 10x Genomics cell capture run (AV1-AV5) and hash-tag antibody (Ab1-15) indicated.

**Figure S11.**
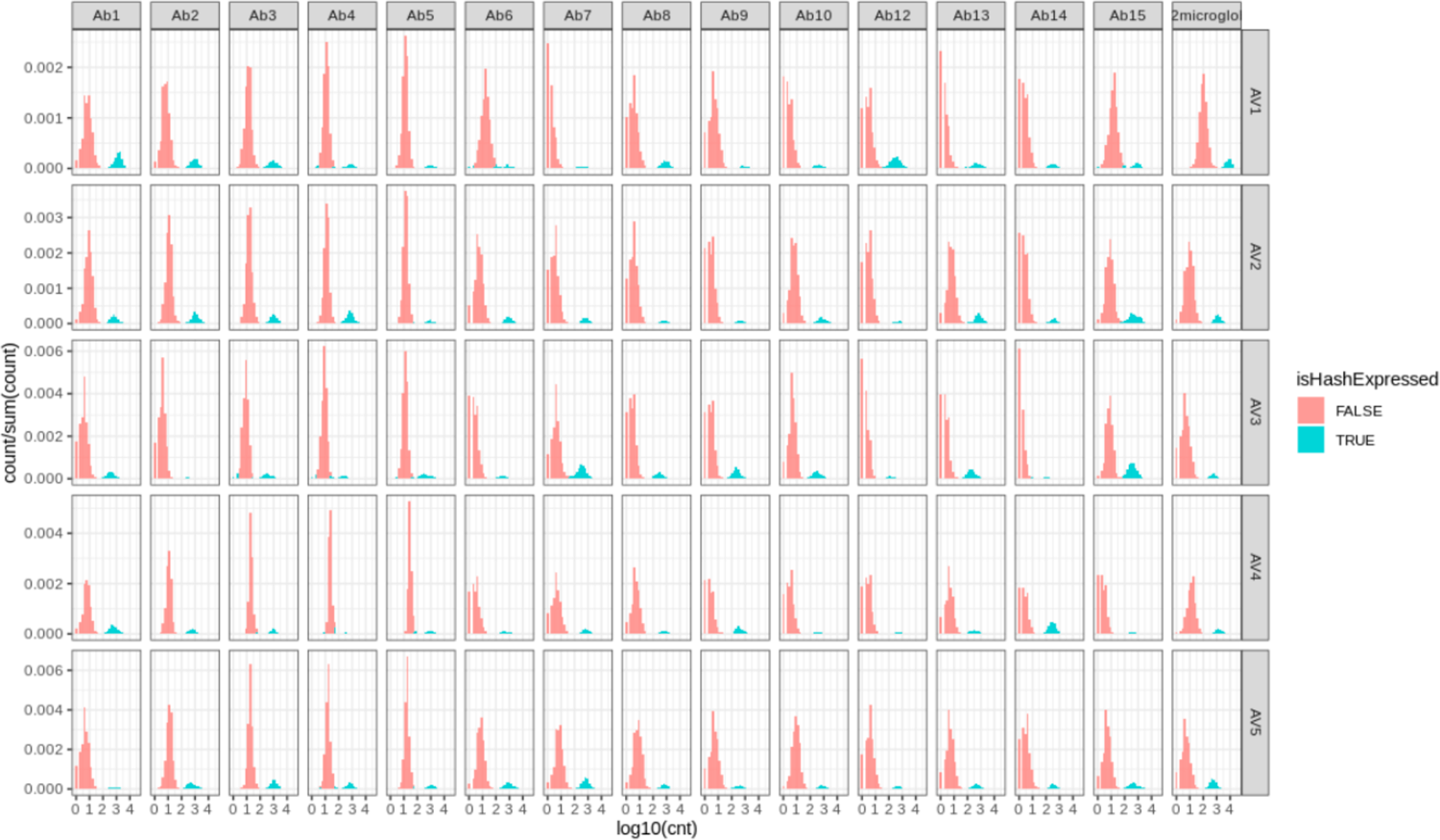
Hash-tag counts classified with mixtures of 2 gaussian models

**Figure S12.**
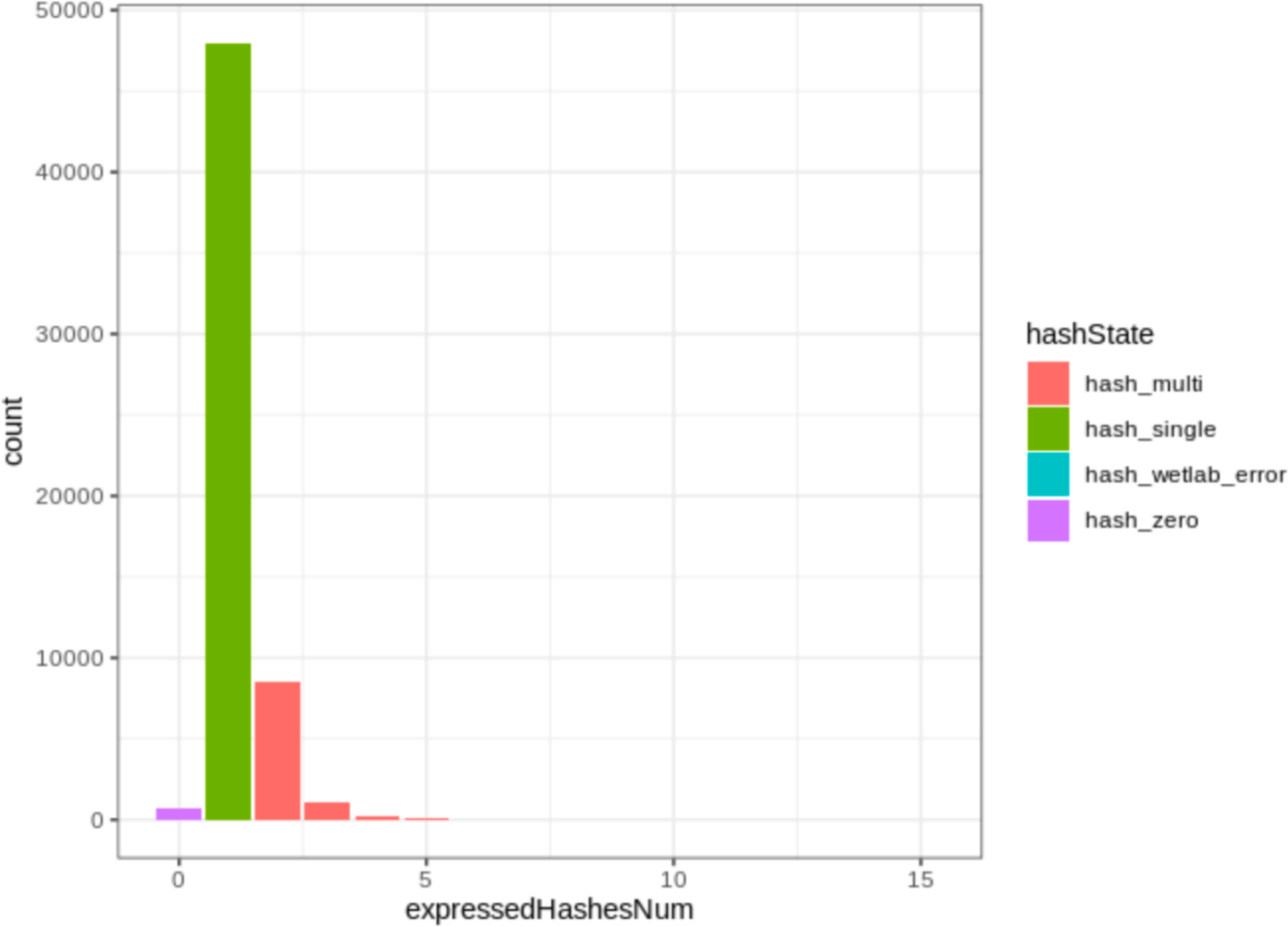
Counting numbers of hashes classified as expressed in each cell

**Figure S13.**
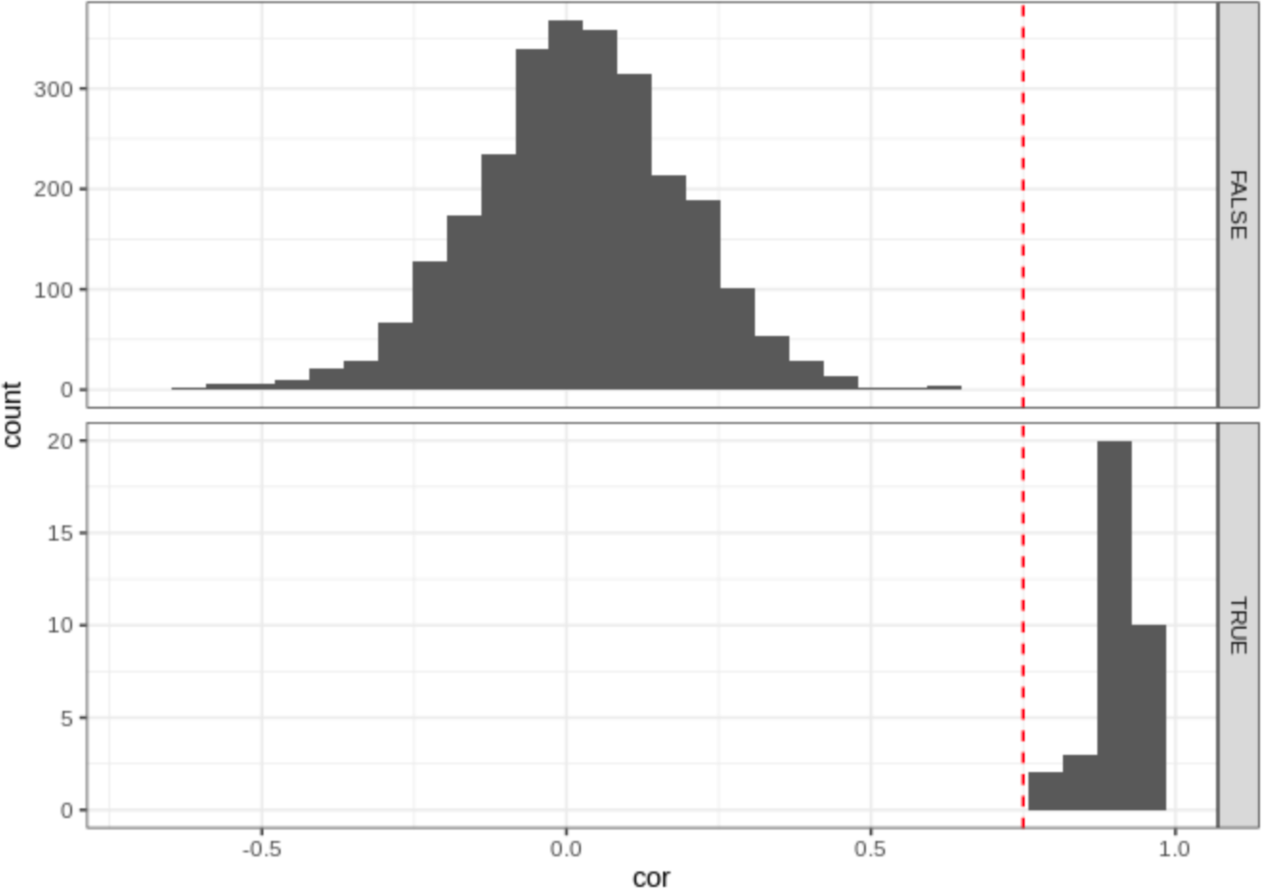
Genotype consistency between samples of the same donor

**Figure S14.**
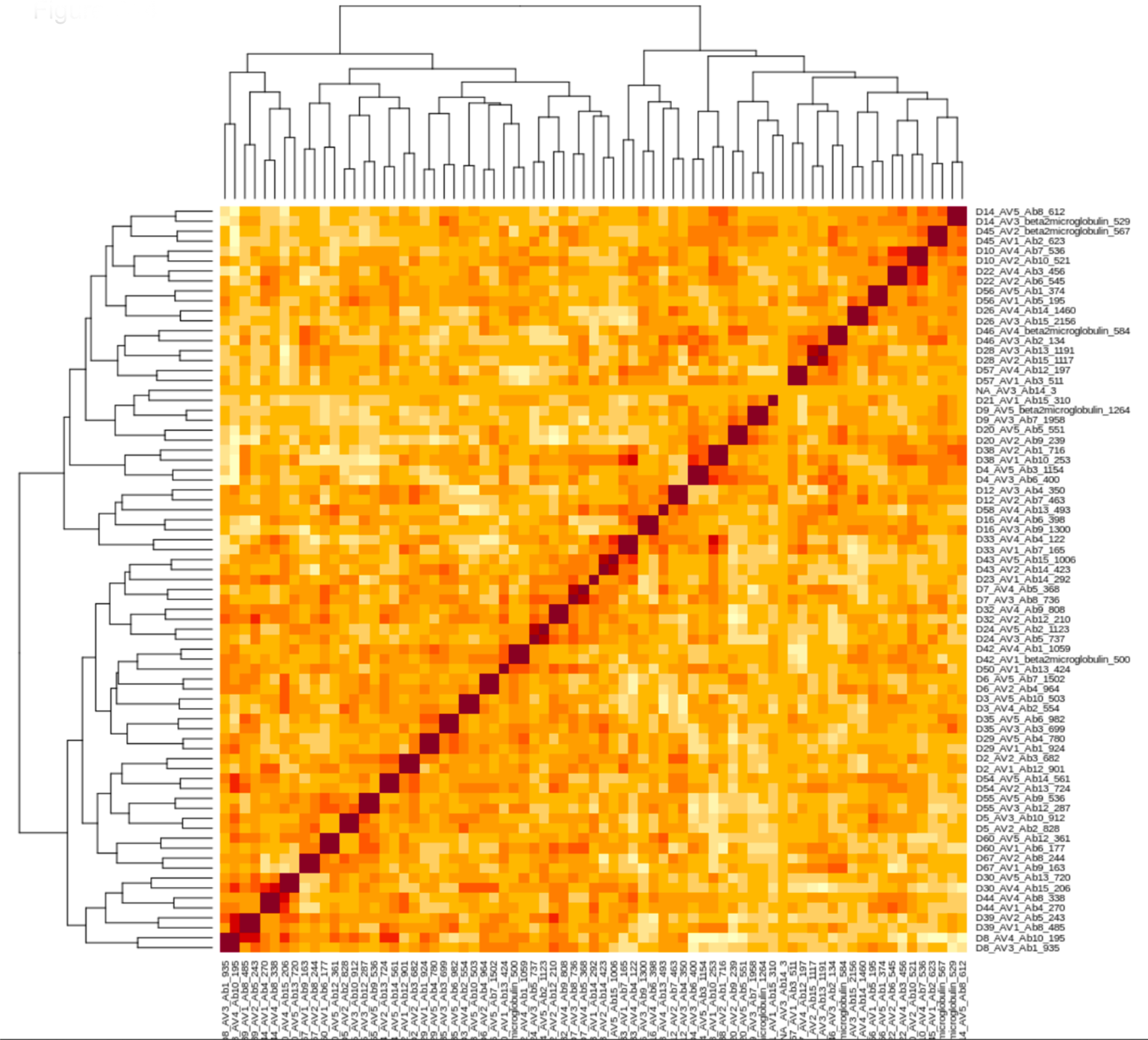
**Comparison of genotype profiles between samples defined by hashes**

**Figure S15.**
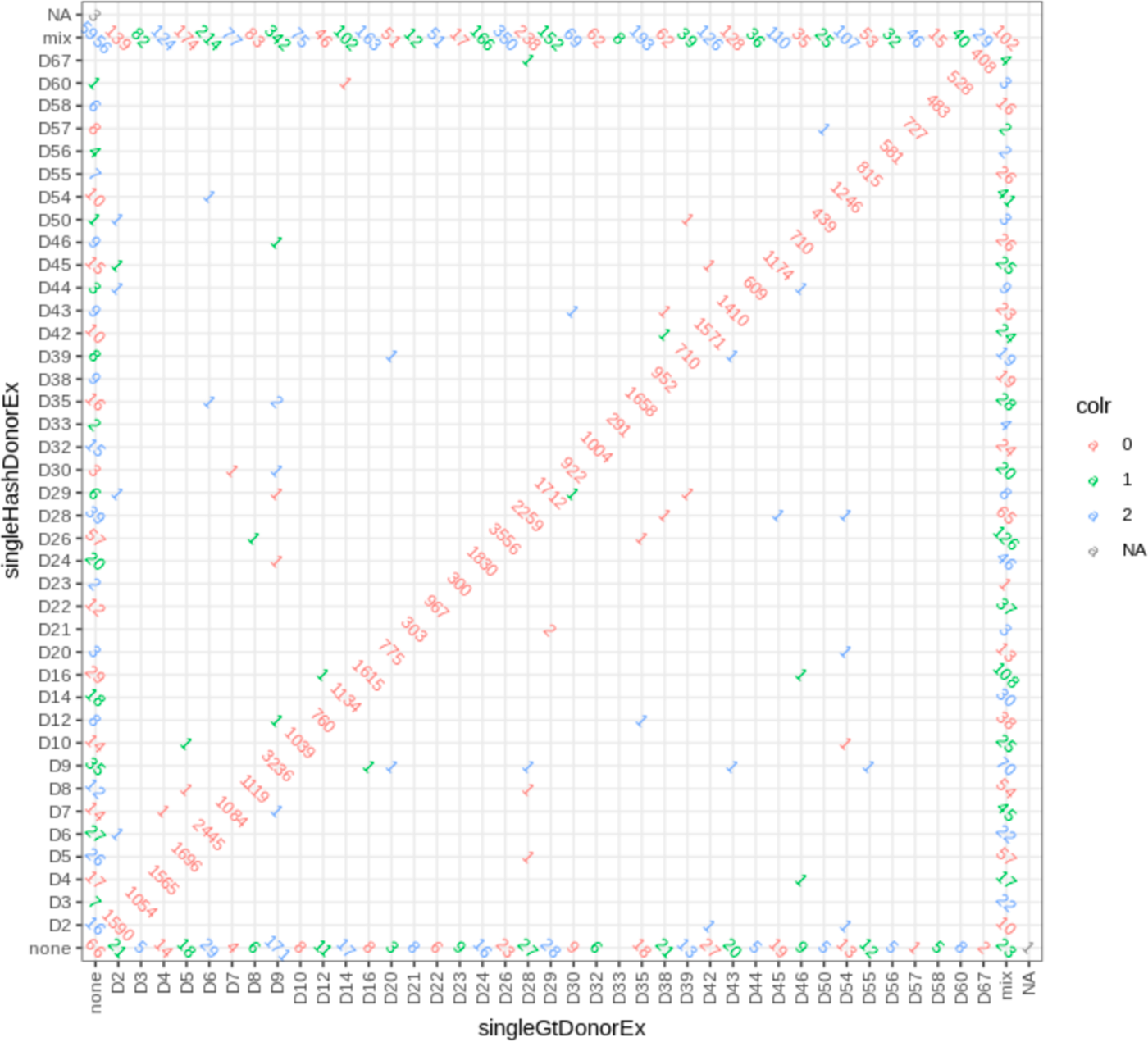
**Numbers of cells with predicted donors by both approaches**

**Table S1.**
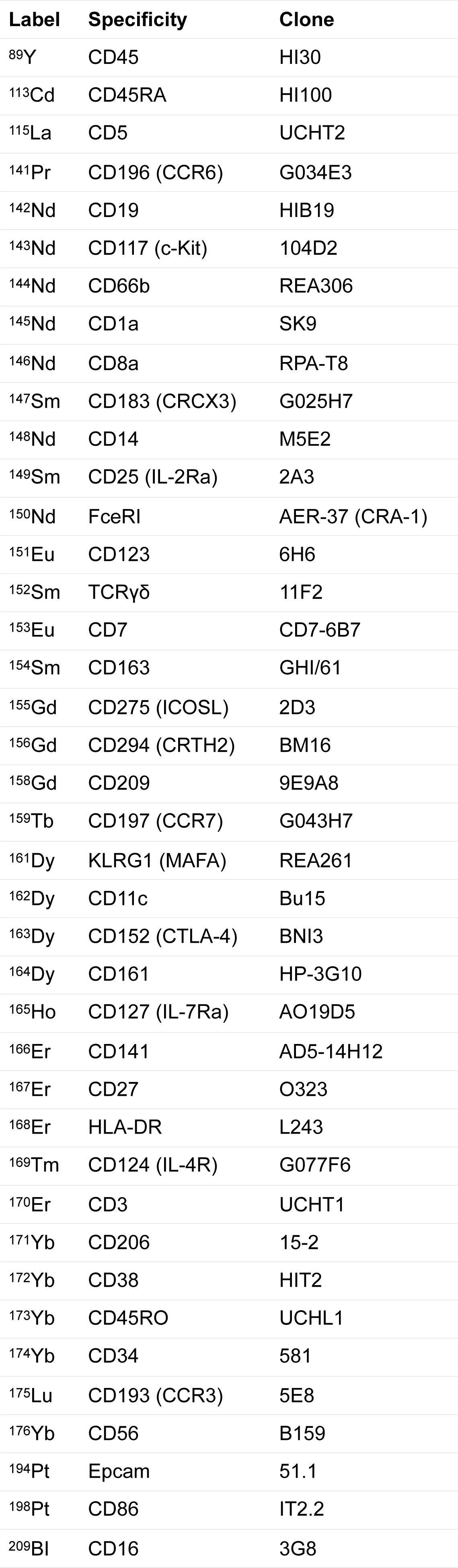
**Mass-cytometry antibody panel**

| Label | Specificity | Clone |
| --- | --- | --- |
| <sup>89</sup>Y | CD45 | HI30 |
| <sup>113</sup>Cd | CD45RA | HI100 |
| <sup>115</sup>La | CD5 | UCHT2 |
| <sup>141</sup>Pr | CD196 (CCR6) | G034E3 |
| <sup>142</sup>Nd | CD19 | HIB19 |
| <sup>143</sup>Nd | CD117 (c-Kit) | 104D2 |
| <sup>144</sup>Nd | CD66b | REA306 |
| <sup>145</sup>Nd | CD1a | SK9 |
| <sup>146</sup>Nd | CD8a | RPA-T8 |
| <sup>147</sup>Sm | CD183 (CRCX3) | G025H7 |
| <sup>148</sup>Nd | CD14 | M5E2 |
| <sup>149</sup>Sm | CD25 (IL-2Ra) | 2A3 |
| <sup>150</sup>Nd | FceRI | AER-37 (CRA-1) |
| <sup>151</sup>Eu | CD123 | 6H6 |
| <sup>152</sup>Sm | TCRγδ | 11F2 |
| <sup>153</sup>Eu | CD7 | CD7-6B7 |
| <sup>154</sup>Sm | CD163 | GHI/61 |
| <sup>155</sup>Gd | CD275 (ICOSL) | 2D3 |
| <sup>156</sup>Gd | CD294 (CRTH2) | BM16 |
| <sup>158</sup>Gd | CD209 | 9E9A8 |
| <sup>159</sup>Tb | CD197 (CCR7) | G043H7 |
| <sup>161</sup>Dy | KLRG1 (MAFA) | REA261 |
| <sup>162</sup>Dy | CD11c | Bu15 |
| <sup>163</sup>Dy | CD152 (CTLA-4) | BNI3 |
| <sup>164</sup>Dy | CD161 | HP-3G10 |
| <sup>165</sup>Ho | CD127 (IL-7Ra) | AO19D5 |
| <sup>166</sup>Er | CD141 | AD5-14H12 |
| <sup>167</sup>Er | CD27 | O323 |
| <sup>168</sup>Er | HLA-DR | L243 |
| <sup>169</sup>Tm | CD124 (IL-4R) | G077F6 |
| <sup>170</sup>Er | CD3 | UCHT1 |
| <sup>171</sup>Yb | CD206 | 15-2 |
| <sup>172</sup>Yb | CD38 | HIT2 |
| <sup>173</sup>Yb | CD45RO | UCHL1 |
| <sup>174</sup>Yb | CD34 | 581 |
| <sup>175</sup>Lu | CD193 (CCR3) | 5E8 |
| <sup>176</sup>Yb | CD56 | B159 |
| <sup>194</sup>Pt | Epcam | 51.1 |
| <sup>198</sup>Pt | CD86 | IT2.2 |
| <sup>209</sup>Bi | CD16 | 3G8 |

