## Supplemental Table for "Nasal systems immunology identifies inflammatory and tolerogenic myeloid cells that determine allergic outcome following challenge"

Table S2. MPS clusters (T4, T5, T13, T15, T18, T22, T23) top DEG

| | p_val | avg_logFC | pct.1 | pct.2 | p_val_adj | cluster | gene |
|---|---|---|---|---|---|---|---|
| 1 | 0 | 2,94181790817356 | 0,945 | 0,198 | 0 | 4 | S100A8 |
| 2 | 0 | 2,68554845391382 | 0,976 | 0,283 | 0 | 4 | S100A9 |
| 3 | 0 | 1,42062506227039 | 0,694 | 0,028 | 0 | 4 | S100A12 |
| 4 | 0 | 1,06178372307441 | 0,997 | 0,694 | 0 | 4 | LYZ |
| 5 | 0 | 1,05816024751746 | 0,936 | 0,288 | 0 | 4 | FCN1 |
| 6 | 0 | 0,881635846760492 | 0,916 | 0,373 | 0 | 4 | CSTA |
| 7 | 0 | 0,837505266544342 | 0,867 | 0,44 | 0 | 4 | MNDA |
| 8 | 0 | 0,814008581594069 | 0,993 | 0,682 | 0 | 4 | S100A4 |
| 9 | 0 | 0,758144680103419 | 0,777 | 0,163 | 0 | 4 | CD14 |
| 10 | 0 | 0,725301020826687 | 0,607 | 0,057 | 0 | 4 | VCAN |
| 11 | 0 | 0,683001330396441 | 0,869 | 0,373 | 0 | 4 | TSPO |
| 12 | 1,17169502726017E-288 | 0,556209147051608 | 0,745 | 0,376 | 1,97723535850154E-284 | 4 | AP1S2 |
| 13 | 1,02600521546594E-265 | 0,491617967991159 | 0,94 | 0,644 | 1,73138380109877E-261 | 4 | CTSS |
| 14 | 1,04727690848423E-224 | 0,488932942845677 | 0,954 | 0,699 | 1,76727978306714E-220 | 4 | LGALS1 |
| 15 | 8,12779607759468E-235 | 0,438264348768507 | 0,968 | 0,725 | 1,3715655880941E-230 | 4 | S100A11 |
| 16 | 8,14646862305862E-143 | 0,435752448976009 | 0,716 | 0,41 | 1,37471658014114E-138 | 4 | CD52 |
| 17 | 2,26157011463434E-282 | 0,434714888355903 | 0,719 | 0,255 | 3,81639956844544E-278 | 4 | CFD |
| 18 | 2,4814025206624E-98 | 0,430786303716304 | 0,349 | 0,144 | 4,1873667536178E-94 | 4 | AC020656.1 |
| 19 | 6,64398173702977E-200 | 0,40711628324242 | 0,836 | 0,575 | 1,12117191812377E-195 | 4 | ELOB |
| 20 | 2,67073962890416E-164 | 0,376682957182231 | 0,677 | 0,369 | 4,50687312377577E-160 | 4 | TXNIP |
| 21 | 2,60176849375222E-185 | 0,373124109258083 | 0,801 | 0,498 | 4,39048433320688E-181 | 4 | GMFG |
| 22 | 5,60415425491092E-228 | 0,364309351446056 | 0,51 | 0,169 | 9,45701030516218E-224 | 4 | BLVRB |
| 23 | 2,04525937509707E-152 | 0,357198040404979 | 0,503 | 0,222 | 3,45137519547631E-148 | 4 | NCF1 |
| 24 | 2,78002500682373E-173 | 0,350237454715217 | 0,62 | 0,302 | 4,69129219901505E-169 | 4 | TKT |
| 25 | 3,10327824293273E-170 | 0,347341885638502 | 0,648 | 0,322 | 5,23678203494898E-166 | 4 | CARD16 |
| 26 | 1,7443137622374E-139 | 0,343798895119498 | 0,654 | 0,366 | 2,9435294737756E-135 | 4 | HCST |
| 27 | 1,61935995932885E-204 | 0,330510595554877 | 0,418 | 0,12 | 2,73266993136744E-200 | 4 | CD36 |
| 28 | 5,85769094656971E-139 | 0,323006026836328 | 0,825 | 0,533 | 9,88485347233639E-135 | 4 | EMP3 |
| 29 | 1,35760916185061E-128 | 0,31637576440276 | 0,716 | 0,482 | 2,29096546062291E-124 | 4 | POLR2L |
| 30 | 3,40749184227634E-114 | 0,298527201890546 | 0,556 | 0,293 | 5,75014248384133E-110 | 4 | ANXA1 |
| 31 | 1,518311033072E-142 | 0,293307787143918 | 0,544 | 0,261 | 2,56214986830899E-138 | 4 | SDCBP |
| 32 | 2,83667103562329E-190 | 0,281767193458411 | 0,38 | 0,103 | 4,78688237261431E-186 | 4 | NUP214 |
| 33 | 6,51450645915483E-212 | 0,272985701588931 | 0,344 | 0,071 | 1,09932296498238E-207 | 4 | ASGR1 |
| 34 | 8,88475479521697E-200 | 0,266927709957105 | 0,274 | 0,042 | 1,49930237169286E-195 | 4 | RBP7 |
| 35 | 5,27874611469388E-181 | 0,26299254355044 | 0,757 | 0,33 | 8,90788406854593E-177 | 4 | SERPINA1 |
| 36 | 3,14154671741958E-173 | 0,260311393051118 | 0,366 | 0,102 | 5,30136008564555E-169 | 4 | AGTRAP |
| 37 | 1,85345532494779E-98 | 0,259654447400819 | 0,645 | 0,37 | 3,12770586084939E-94 | 4 | LGALS2 |
| 38 | 9,05487068218888E-139 | 0,251549704229062 | 0,982 | 0,718 | 1,52800942761937E-134 | 4 | AIF1 |
| 39 | 0 | 1,14602907337962 | 0,978 | 0,567 | 0 | 5 | HLA-DQB1 |
| 40 | 0 | 1,10951696873443 | 0,96 | 0,418 | 0 | 5 | HLA-DQA1 |
| 41 | 0 | 1,08354673322767 | 0,64 | 0,124 | 0 | 5 | FCER1A |
| 42 | 5,94472250894005E-158 | 0,718701006601099 | 0,284 | 0,063 | 1,00317192338363E-153 | 5 | C15orf48 |
| 43 | 8,95554653491727E-166 | 0,688374052059381 | 0,52 | 0,25 | 1,51124847776729E-161 | 5 | HLA-DQA2 |
| 44 | 0 | 0,671730153252914 | 0,672 | 0,235 | 0 | 5 | GSN |
| 45 | 0 | 0,623703892762301 | 0,458 | 0,049 | 0 | 5 | CD1C |
| 46 | 0 | 0,61494227488545 | 0,655 | 0,238 | 0 | 5 | PPA1 |
| 47 | 0 | 0,600628832392458 | 0,373 | 0,046 | 0 | 5 | CD1E |
| 48 | 5,69419933223333E-299 | 0,592219752419843 | 0,492 | 0,121 | 9,60896137314375E-295 | 5 | CLEC10A |
| 49 | 3,17659829183847E-303 | 0,52457672595851 | 0,704 | 0,3 | 5,36050961747742E-299 | 5 | SPINT2 |
| 50 | 2,29464225256746E-274 | 0,510054136207775 | 0,733 | 0,359 | 3,87220880120759E-270 | 5 | CTSH |
| 51 | 1,40064997563332E-268 | 0,489760490944801 | 0,905 | 0,605 | 2,36359683388123E-264 | 5 | HLA-DMA |
| 52 | 8,73783688369934E-281 | 0,487861159472378 | 0,387 | 0,066 | 1,47450997412426E-276 | 5 | PPP1R14A |
| 53 | 2,33158885700966E-250 | 0,485225430701546 | 0,797 | 0,459 | 3,9345561962038E-246 | 5 | HLA-DMB |
| 54 | 4,30915973942849E-158 | 0,485099500977446 | 0,541 | 0,258 | 7,27170706028558E-154 | 5 | YWHAH |
| 55 | 2,04552688943032E-159 | 0,437620060156631 | 0,558 | 0,276 | 3,45182662591366E-155 | 5 | LITAF |
| 56 | 0 | 0,433715746779933 | 0,394 | 0,056 | 0 | 5 | PKIB |
| 57 | 4,31499218659841E-200 | 0,414819151846946 | 0,597 | 0,27 | 7,28154931488482E-196 | 5 | ALDH2 |
| 58 | 7,69322508340461E-128 | 0,394024708021171 | 0,377 | 0,14 | 1,29823173282453E-123 | 5 | RGS1 |
| 59 | 4,48514999083926E-124 | 0,380690523955023 | 0,733 | 0,492 | 7,56869060954125E-120 | 5 | TUBA1B |
| 60 | 1,33154629358584E-149 | 0,37822885703608 | 0,819 | 0,598 | 2,24698437042611E-145 | 5 | LSP1 |
| 61 | 2,78725815566405E-174 | 0,355494952597838 | 0,489 | 0,193 | 4,70349813768308E-170 | 5 | CSF2RA |
| 62 | 4,02875139740015E-111 | 0,322129312072197 | 0,661 | 0,429 | 6,79851798311275E-107 | 5 | JAML |
| 63 | 7,41437399836229E-176 | 0,316714631603961 | 0,307 | 0,065 | 1,25117561222364E-171 | 5 | AXL |
| 64 | 3,12504160426729E-185 | 0,314103004253374 | 0,339 | 0,082 | 5,27350770720106E-181 | 5 | RUNX3 |

| | | | | | | | |
|---|---|---|---|---|---|---|---|
| 65 | 4,05012126398628E-212 | 0,310743569054977 | 0,365 | 0,081 | 6,83457963297685E-208 | 5 | BASP1 |
| 66 | 6,6579227949499E-250 | 0,304824230470796 | 0,366 | 0,065 | 1,1235244716478E-245 | 5 | NDRG2 |
| 67 | 2,40416889861556E-180 | 0,300763983912441 | 0,323 | 0,072 | 4,05703501641376E-176 | 5 | FCGR2B |
| 68 | 5,01576620823869E-96 | 0,294624082416643 | 0,575 | 0,361 | 8,46410547640278E-92 | 5 | SYNGR2 |
| 69 | 2,82457617955847E-68 | 0,293365650424134 | 0,599 | 0,391 | 4,76647230300491E-64 | 5 | LGALS2 |
| 70 | 1,45579886796442E-146 | 0,273207037632216 | 0,421 | 0,156 | 2,45666058968995E-142 | 5 | KCNK6 |
| 71 | 4,19052768008862E-201 | 0,250623566746464 | 0,236 | 0,025 | 7,07151546014955E-197 | 5 | ENHO |
| 72 | 0 | 1,43392642310962 | 0,919 | 0,083 | 0 | 13 | FCGR3A |
| 73 | 0 | 1,30946847808565 | 0,997 | 0,769 | 0 | 13 | LST1 |
| 74 | 0 | 1,1179620328285 | 0,918 | 0,373 | 0 | 13 | IFITM2 |
| 75 | 0 | 1,07355559567277 | 0,877 | 0,157 | 0 | 13 | SMIM25 |
| 76 | 0 | 1,00669601371601 | 0,986 | 0,764 | 0 | 13 | AIF1 |
| 77 | 0 | 0,959958456565213 | 0,953 | 0,374 | 0 | 13 | SERPINA1 |
| 78 | 0 | 0,958294084469287 | 0,991 | 0,755 | 0 | 13 | SAT1 |
| 79 | 0 | 0,937885906673504 | 0,875 | 0,374 | 0 | 13 | IFITM3 |
| 80 | 3,47642315650497E-247 | 0,936518990743884 | 0,85 | 0,443 | 5,86646407660213E-243 | 13 | CD52 |
| 81 | 0 | 0,779123809130126 | 0,875 | 0,313 | 0 | 13 | CFD |
| 82 | 1,71139436327876E-304 | 0,775743960933186 | 0,987 | 0,689 | 2,8879779880329E-300 | 13 | CTSS |
| 83 | 0 | 0,769929795442381 | 0,804 | 0,219 | 0 | 13 | MS4A7 |
| 84 | 1,00194262391999E-221 | 0,713896670397832 | 0,996 | 0,737 | 1,69077817786498E-217 | 13 | S100A4 |
| 85 | 0 | 0,683067793029035 | 0,684 | 0,147 | 0 | 13 | RHOC |
| 86 | 1,63519500861211E-228 | 0,656971500247463 | 0,681 | 0,249 | 2,75939157703294E-224 | 13 | CEBPB |
| 87 | 7,03167624890477E-245 | 0,635433888089255 | 0,731 | 0,274 | 1,18659536700268E-240 | 13 | LY6E |
| 88 | 1,69807468608788E-291 | 0,62577685928553 | 0,793 | 0,294 | 2,86550103277329E-287 | 13 | STXBP2 |
| 89 | 0 | 0,591844943356798 | 0,244 | 0,003 | 0 | 13 | LYPD2 |
| 90 | 1,13574457953661E-199 | 0,576519721544852 | 0,831 | 0,506 | 1,91656897796804E-195 | 13 | NAP1L1 |
| 91 | 8,86361463303461E-164 | 0,537352565183009 | 0,942 | 0,699 | 1,49573496932459E-159 | 13 | COTL1 |
| 92 | 1,35435517982322E-184 | 0,532851072660341 | 0,972 | 0,768 | 2,28547436595168E-180 | 13 | S100A11 |
| 93 | 1,90160766718951E-176 | 0,523031831321109 | 0,843 | 0,494 | 3,2089629383823E-172 | 13 | HLA-E |
| 94 | 1,92851746187013E-160 | 0,518509629248683 | 0,895 | 0,574 | 3,25437321690584E-156 | 13 | EMP3 |
| 95 | 7,9987322837199E-217 | 0,507506323031764 | 0,777 | 0,339 | 1,34978607287773E-212 | 13 | ASAH1 |
| 96 | 3,80813030260439E-213 | 0,505309210222172 | 0,703 | 0,289 | 6,42621988564491E-209 | 13 | PTPN6 |
| 97 | 5,60809554329605E-237 | 0,498571827778087 | 0,68 | 0,216 | 9,46366122931209E-233 | 13 | MAFB |
| 98 | 1,67289815145503E-290 | 0,495542606086777 | 0,69 | 0,198 | 2,82301563058036E-286 | 13 | TNFRSF1B |
| 99 | 9,94045610207566E-143 | 0,487972580486967 | 0,903 | 0,608 | 1,67745196722527E-138 | 13 | NEAT1 |
| 100 | 1,49250305745804E-134 | 0,484256564052365 | 0,538 | 0,208 | 2,51859890946045E-130 | 13 | MT2A |
| 101 | 7,17287508481437E-269 | 0,474827527103985 | 0,683 | 0,208 | 1,21042267056243E-264 | 13 | LRRC25 |
| 102 | 3,28695087144831E-170 | 0,473468056984398 | 0,837 | 0,487 | 5,54672959556903E-166 | 13 | CD68 |
| 103 | 3,06106381263472E-281 | 0,464981861725148 | 0,664 | 0,182 | 5,16554518382109E-277 | 13 | PILRA |
| 104 | 3,59863716275658E-90 | 0,445019569109305 | 0,86 | 0,644 | 6,07270021215173E-86 | 13 | JUNB |
| 105 | 1,82340076617278E-177 | 0,440910534683912 | 0,588 | 0,203 | 3,07698879291656E-173 | 13 | VAMP5 |
| 106 | 7,97526829999411E-148 | 0,438730144868186 | 0,882 | 0,632 | 1,34582652562401E-143 | 13 | ARHGDIB |
| 107 | 9,35988134737611E-173 | 0,435039570652641 | 0,763 | 0,369 | 1,57947997736972E-168 | 13 | HCK |
| 108 | 9,32763333785543E-155 | 0,421376585030211 | 0,752 | 0,363 | 1,5740381257631E-150 | 13 | CARD16 |
| 109 | 3,92922789807072E-132 | 0,420034076883538 | 0,887 | 0,602 | 6,63057207799435E-128 | 13 | SPI1 |
| 110 | 5,50898892816121E-135 | 0,414506531355501 | 0,345 | 0,087 | 9,29641881627205E-131 | 13 | IFITM1 |
| 111 | 1,03582089471308E-234 | 0,413857674127046 | 0,593 | 0,166 | 1,74794775982832E-230 | 13 | LILRB2 |
| 112 | 0 | 2,22589836465038 | 0,965 | 0,041 | 0 | 15 | GZMB |
| 113 | 0 | 2,04563886276914 | 0,963 | 0,061 | 0 | 15 | JCHAIN |
| 114 | 0 | 1,53713635482852 | 0,447 | 0,012 | 0 | 15 | PTGDS |
| 115 | 0 | 1,41246722298492 | 0,911 | 0,099 | 0 | 15 | ITM2C |
| 116 | 0 | 1,27536682422563 | 0,895 | 0,197 | 0 | 15 | IGKC |
| 117 | 0 | 1,25773552343251 | 0,866 | 0,131 | 0 | 15 | IRF7 |
| 118 | 0 | 1,21113596832395 | 0,886 | 0,188 | 0 | 15 | IRF8 |
| 119 | 0 | 1,17451382118443 | 0,891 | 0,196 | 0 | 15 | ALOX5AP |
| 120 | 0 | 1,0642228948237 | 0,802 | 0,076 | 0 | 15 | PPP1R14B |
| 121 | 0 | 0,985701063814572 | 0,769 | 0,055 | 0 | 15 | C12orf75 |
| 122 | 0 | 0,944364267019388 | 0,893 | 0,354 | 0 | 15 | PLAC8 |
| 123 | 0 | 0,916081659016699 | 0,68 | 0,03 | 0 | 15 | CLIC3 |
| 124 | 0 | 0,902631787044511 | 0,803 | 0,09 | 0 | 15 | SERPINF1 |
| 125 | 0 | 0,882706638171575 | 0,689 | 0,044 | 0 | 15 | LILRA4 |
| 126 | 0 | 0,857152680939864 | 0,868 | 0,373 | 0 | 15 | HERPUD1 |
| 127 | 0 | 0,854011293641436 | 0,892 | 0,447 | 0 | 15 | SEC61B |
| 128 | 0 | 0,845201250705301 | 0,829 | 0,244 | 0 | 15 | PLD4 |
| 129 | 0 | 0,839529790362192 | 0,783 | 0,079 | 0 | 15 | TCF4 |
| 130 | 0 | 0,797750745469631 | 0,664 | 0,007 | 0 | 15 | MZB1 |

| | | | | | | | |
|---|---|---|---|---|---|---|---|
| 131 | 0 | 0,766814856609796 | 0,741 | 0,088 | 0 | 15 | CYB561A3 |
| 132 | 0 | 0,758677300705738 | 0,744 | 0,108 | 0 | 15 | APP |
| 133 | 0 | 0,751265413000711 | 0,633 | 0,003 | 0 | 15 | PTCRA |
| 134 | 0 | 0,748835759909592 | 0,726 | 0,098 | 0 | 15 | CCDC50 |
| 135 | 0 | 0,728747181215155 | 0,833 | 0,352 | 0 | 15 | SPCS1 |
| 136 | 0 | 0,722939323315561 | 0,664 | 0,007 | 0 | 15 | DERL3 |
| 137 | 0 | 0,710084000220766 | 0,748 | 0,163 | 0 | 15 | SELENOS |
| 138 | 0 | 0,693459449806075 | 0,696 | 0,1 | 0 | 15 | BCL11A |
| 139 | 0 | 0,693397087770158 | 0,66 | 0,046 | 0 | 15 | UGCG |
| 140 | 0 | 0,686585822695133 | 0,258 | 0,002 | 0 | 15 | TCL1A |
| 141 | 2,13985762792034E-138 | 0,662299652266708 | 0,543 | 0,209 | 3,61100974711558E-134 | 15 | LTB |
| 142 | 0 | 0,646641605897249 | 0,586 | 0,02 | 0 | 15 | TSPAN13 |
| 143 | 0 | 0,632031979656362 | 0,585 | 0,004 | 0 | 15 | LRRC26 |
| 144 | 0 | 0,617590211140189 | 0,575 | 0,031 | 0 | 15 | SPIB |
| 145 | 3,30463672263704E-104 | 0,614701279895416 | 0,287 | 0,078 | 5,57657446945E-100 | 15 | IGLC2 |
| 146 | 0 | 0,606230330648473 | 0,615 | 0,048 | 0 | 15 | IL3RA |
| 147 | 1,87848907330777E-296 | 0,597538237348262 | 0,72 | 0,22 | 3,16995031120685E-292 | 15 | PLP2 |
| 148 | 0 | 0,597110451141927 | 0,606 | 0,063 | 0 | 15 | STMN1 |
| 149 | 0 | 0,573763761523883 | 0,493 | 0,007 | 0 | 15 | SCT |
| 150 | 1,46496858183749E-194 | 0,555077350952349 | 0,863 | 0,625 | 2,47213448185076E-190 | 15 | SRP14 |
| 151 | 6,5929256496841E-192 | 0,539714782100013 | 0,814 | 0,465 | 1,11255620338419E-187 | 15 | SSR4 |
| 152 | 0 | 1,82159793931468 | 0,965 | 0,105 | 0 | 18 | C1QA |
| 153 | 0 | 1,81965637241747 | 0,913 | 0,035 | 0 | 18 | C1QC |
| 154 | 0 | 1,7863435332739 | 0,908 | 0,051 | 0 | 18 | C1QB |
| 155 | 1,14517416399987E-130 | 0,699337100794721 | 0,941 | 0,64 | 1,93248140174978E-126 | 18 | ITM2B |
| 156 | 2,6928342539932E-97 | 0,638885702241315 | 0,953 | 0,665 | 4,54415780361352E-93 | 18 | HLA-DQB1 |
| 157 | 5,29631509204625E-54 | 0,612769571435498 | 0,58 | 0,332 | 8,93753171782805E-50 | 18 | CTSD |
| 158 | 2,2918770606463E-102 | 0,589896497045587 | 0,953 | 0,546 | 3,86754253984062E-98 | 18 | HLA-DQA1 |
| 159 | 2,03058070629593E-114 | 0,589879005935443 | 0,864 | 0,486 | 3,42660494187438E-110 | 18 | CD63 |
| 160 | 4,99066990159062E-128 | 0,570881428900205 | 0,961 | 0,671 | 8,42175545893417E-124 | 18 | HLA-DMA |
| 161 | 2,46493015070005E-54 | 0,560204735211477 | 0,536 | 0,254 | 4,15956962930633E-50 | 18 | FCER1A |
| 162 | 4,00801954264885E-113 | 0,536173595215775 | 0,496 | 0,15 | 6,76353297821994E-109 | 18 | ACP5 |
| 163 | 1,86119852954374E-84 | 0,535918295453096 | 0,862 | 0,569 | 3,14077251860506E-80 | 18 | MS4A6A |
| 164 | 6,04977792962677E-75 | 0,523451018695978 | 0,326 | 0,09 | 1,02090002562452E-70 | 18 | HLA-DQB2 |
| 165 | 6,00803105948161E-66 | 0,479471655799858 | 0,472 | 0,188 | 1,01385524128752E-61 | 18 | RGS1 |
| 166 | 6,29398120352664E-94 | 0,47293511414538 | 0,872 | 0,533 | 1,06210932809512E-89 | 18 | HLA-DMB |
| 167 | 9,55568165457013E-120 | 0,455854928159353 | 0,445 | 0,11 | 1,61252127920871E-115 | 18 | AXL |
| 168 | 5,78117494096885E-55 | 0,446097861467555 | 0,597 | 0,32 | 9,75573271288493E-51 | 18 | YWHAH |
| 169 | 7,86883717411573E-90 | 0,444767104634831 | 0,622 | 0,256 | 1,32786627313203E-85 | 18 | MAFB |
| 170 | 4,17875938636907E-61 | 0,431059311249947 | 0,765 | 0,501 | 7,05165646449781E-57 | 18 | CTSB |
| 171 | 7,27680639205466E-74 | 0,429487498671604 | 0,81 | 0,516 | 1,22796107865922E-69 | 18 | CD68 |
| 172 | 2,7783507748263E-100 | 0,416710001522469 | 0,659 | 0,271 | 4,68846693251938E-96 | 18 | CSF1R |
| 173 | 4,23949505645733E-51 | 0,384536340418837 | 0,792 | 0,544 | 7,15414790777174E-47 | 18 | TUBA1B |
| 174 | 1,26646459337295E-57 | 0,37718978475405 | 0,573 | 0,289 | 2,13715900131686E-53 | 18 | CALHM6 |
| 175 | 7,31593709511716E-39 | 0,372892834798236 | 0,588 | 0,327 | 1,23456438480102E-34 | 18 | CD14 |
| 176 | 1,68747630322996E-87 | 0,353520040668145 | 0,417 | 0,124 | 2,84761626170055E-83 | 18 | FCGR2B |
| 177 | 3,02776609528088E-196 | 0,350418340507189 | 0,308 | 0,029 | 5,10935528578648E-192 | 18 | A2M |
| 178 | 3,51932759313462E-60 | 0,348116822465892 | 0,467 | 0,189 | 5,93886531341468E-56 | 18 | TMEM176B |
| 179 | 3,51835586167047E-37 | 0,337786797706115 | 0,608 | 0,372 | 5,93722551656892E-33 | 18 | CTSC |
| 180 | 3,11950541494477E-49 | 0,334509552424066 | 0,77 | 0,496 | 5,26416538771931E-45 | 18 | CPVL |
| 181 | 1,22168240370874E-44 | 0,319579711056474 | 0,709 | 0,449 | 2,0615890562585E-40 | 18 | CTSH |
| 182 | 1,99143890187177E-36 | 0,31188001667287 | 0,534 | 0,304 | 3,36055314690862E-32 | 18 | CEBPD |
| 183 | 4,05659804383749E-37 | 0,302559232236215 | 0,708 | 0,502 | 6,84550919897576E-33 | 18 | ATP6V1F |
| 184 | 9,49869085309694E-44 | 0,298824249005023 | 0,462 | 0,211 | 1,60290408146011E-39 | 18 | CLEC10A |
| 185 | 3,06175680565339E-40 | 0,295289784330822 | 0,697 | 0,45 | 5,1667146095401E-36 | 18 | FGL2 |
| 186 | 1,60120562172125E-80 | 0,289852272149423 | 0,402 | 0,125 | 2,70203448665461E-76 | 18 | AKR1B1 |
| 187 | 1,04327948648112E-39 | 0,285747139570718 | 0,576 | 0,335 | 1,76053413343688E-35 | 18 | IGSF6 |
| 188 | 1,30003414766398E-69 | 0,266860029647897 | 0,388 | 0,129 | 2,19380762418297E-65 | 18 | RB1 |
| 189 | 1,121295085102E-39 | 0,259504826476738 | 0,536 | 0,295 | 1,89218545610963E-35 | 18 | TGFBI |
| 190 | 1,27270910986488E-43 | 0,252335282038921 | 0,471 | 0,227 | 2,14769662289699E-39 | 18 | RAB32 |
| 191 | 0 | 1,36371543712798 | 0,958 | 0,099 | 0 | 22 | C1orf54 |
| 192 | 2,21418620940386E-114 | 1,28033768218595 | 0,933 | 0,504 | 3,73643922836901E-110 | 22 | CPVL |
| 193 | 9,43907092824985E-126 | 1,20606607898262 | 0,95 | 0,523 | 1,59284321914216E-121 | 22 | SNX3 |
| 194 | 0 | 1,07740373654784 | 0,721 | 0,051 | 0 | 22 | RGCC |
| 195 | 1,12480751562141E-153 | 1,05257488776624 | 0,95 | 0,263 | 1,89811268261113E-149 | 22 | IRF8 |
| 196 | 8,57492534091269E-184 | 1,04620934143191 | 0,833 | 0,158 | 1,44701865127902E-179 | 22 | DNASE1L3 |

| | | | | | | | |
|---|---|---|---|---|---|---|---|
| 197 | 3,53616783388692E-137 | 0,986810861485166 | 0,883 | 0,265 | 5,96728321968417E-133 | 22 | ID2 |
| 198 | 2,80332882310252E-79 | 0,939619455543022 | 0,992 | 0,678 | 4,7306173889855E-75 | 22 | HLA-DQB1 |
| 199 | 1,21797174775315E-83 | 0,914897961392797 | 1 | 0,564 | 2,05532732433345E-79 | 22 | HLA-DQA1 |
| 200 | 1,43518699123508E-24 | 0,774256296451502 | 0,583 | 0,352 | 2,4218780477092E-20 | 22 | HLA-DRB5 |
| 201 | 0 | 0,768496518269291 | 0,713 | 0,003 | 0 | 22 | CLEC9A |
| 202 | 5,90962495733878E-76 | 0,728606825699989 | 0,863 | 0,41 | 9,9724921155092E-72 | 22 | PPT1 |
| 203 | 0 | 0,706986250887686 | 0,596 | 0,022 | 0 | 22 | IDO1 |
| 204 | 0 | 0,599250669255018 | 0,429 | 0,004 | 0 | 22 | TACSTD2 |
| 205 | 1,36084910325808E-45 | 0,578726430035889 | 0,833 | 0,44 | 2,29643286174801E-41 | 22 | LGALS2 |
| 206 | 6,21354578159309E-15 | 0,578008810909464 | 0,6 | 0,39 | 1,04853585064383E-10 | 22 | TXN |
| 207 | 1,10378078655243E-71 | 0,564709510174629 | 0,825 | 0,339 | 1,86263007730723E-67 | 22 | RGS10 |
| 208 | 6,39945523590274E-86 | 0,559595946468741 | 0,621 | 0,163 | 1,07990807105859E-81 | 22 | TAP1 |
| 209 | 9,80852366325607E-61 | 0,542811269444766 | 0,283 | 0,046 | 1,65518836817446E-56 | 22 | S100B |
| 210 | 1,41124305130023E-47 | 0,541419820478045 | 0,971 | 0,752 | 2,38147264906914E-43 | 22 | VIM |
| 211 | 9,05686671635002E-73 | 0,538333775136183 | 0,692 | 0,232 | 1,52834625838407E-68 | 22 | RAB32 |
| 212 | 6,45607432588447E-304 | 0,517415910103593 | 0,475 | 0,022 | 1,089462542493E-299 | 22 | CCND1 |
| 213 | 6,10427550842271E-119 | 0,508251968434121 | 0,55 | 0,094 | 1,03009649204633E-114 | 22 | CPNE3 |
| 214 | 4,56394364613604E-40 | 0,499390574386018 | 0,867 | 0,658 | 7,70165490285457E-36 | 22 | EEF1B2 |
| 215 | 6,69015110954839E-48 | 0,487048155411301 | 0,933 | 0,654 | 1,12896299973629E-43 | 22 | LSP1 |
| 216 | 3,00405151403043E-127 | 0,463599429922657 | 0,471 | 0,063 | 5,06933692992635E-123 | 22 | ASB2 |
| 217 | 3,65877397667397E-28 | 0,45281565570846 | 0,854 | 0,592 | 6,17418108563733E-24 | 22 | S100A10 |
| 218 | 0 | 0,451105505644995 | 0,412 | 0,006 | 0 | 22 | RAB7B |
| 219 | 3,27736015690954E-72 | 0,427940014281252 | 0,575 | 0,151 | 5,53054526478485E-68 | 22 | BASP1 |
| 220 | 4,73188844631325E-30 | 0,427142982216962 | 0,471 | 0,196 | 7,98506175315362E-26 | 22 | SERPINB9 |
| 221 | 4,66661623484475E-39 | 0,426639085228253 | 0,954 | 0,685 | 7,87491489630052E-35 | 22 | HLA-DMA |
| 222 | 2,36451103364712E-166 | 0,424823901147971 | 0,5 | 0,053 | 3,99011236927951E-162 | 22 | WDFY4 |
| 223 | 3,03103273626472E-76 | 0,422497058316155 | 0,579 | 0,151 | 5,11486774244671E-72 | 22 | CYB5R3 |
| 224 | 6,45220073205221E-33 | 0,391771589403535 | 0,821 | 0,492 | 1,08880887353381E-28 | 22 | TAGLN2 |
| 225 | 0 | 0,371532917734784 | 0,358 | 0,003 | 0 | 22 | CADM1 |
| 226 | 3,75928635351186E-75 | 0,365792288488009 | 0,408 | 0,075 | 6,34379572155126E-71 | 22 | LMNA |
| 227 | 3,35300393907886E-37 | 0,350956466442214 | 0,583 | 0,245 | 5,65819414719558E-33 | 22 | NAAA |
| 228 | 2,98269215130008E-83 | 0,338028309009676 | 0,446 | 0,082 | 5,03329300531889E-79 | 22 | SHTN1 |
| 229 | 3,10133090822414E-25 | 0,337833857935605 | 0,35 | 0,121 | 5,23349590762823E-21 | 22 | C15orf48 |
| 230 | 3,06157844487259E-57 | 0,337175402243609 | 0,483 | 0,129 | 5,1664136257225E-53 | 22 | FNBP1 |
| 231 | 9,73901424446881E-36 | 0,862973165477557 | 0,879 | 0,647 | 1,64345865375411E-31 | 23 | NEAT1 |
| 232 | 6,86125509866749E-29 | 0,751013960229614 | 0,714 | 0,34 | 1,15783679790014E-24 | 23 | CD14 |
| 233 | 1,67880298011891E-25 | 0,687220154724265 | 0,857 | 0,475 | 2,83298002895066E-21 | 23 | FCN1 |
| 234 | 3,65308814207421E-134 | 0,615727168819534 | 0,821 | 0,115 | 6,16458623975023E-130 | 23 | CCL5 |
| 235 | 6,48146296914655E-26 | 0,576707452705192 | 0,957 | 0,494 | 1,09374687604348E-21 | 23 | CD52 |
| 236 | 1,11238086591801E-14 | 0,510777863484387 | 0,557 | 0,35 | 1,87714271123664E-10 | 23 | MT-ND4L |
| 237 | 5,13381992153067E-17 | 0,503580681566182 | 0,671 | 0,448 | 8,663321117583E-13 | 23 | HCST |
| 238 | 7,4627824025844E-22 | 0,500908285636447 | 0,521 | 0,219 | 1,25934453043612E-17 | 23 | CSF3R |
| 239 | 1,65609239710607E-22 | 0,488372700238446 | 0,55 | 0,216 | 2,79465592011649E-18 | 23 | VCAN |
| 240 | 9,54297111357966E-25 | 0,484587151614849 | 0,871 | 0,539 | 1,61037637541657E-20 | 23 | HLA-E |
| 241 | 6,0090639483519E-15 | 0,44993607473818 | 0,686 | 0,419 | 1,01402954128438E-10 | 23 | JUN |
| 242 | 1,44859830525269E-72 | 0,399493506175998 | 0,479 | 0,07 | 2,44450964011391E-68 | 23 | CD3D |
| 243 | 8,02229713773067E-10 | 0,329492513967532 | 0,621 | 0,39 | 1,35376264199205E-05 | 23 | CFD |
| 244 | 8,02251940471493E-35 | 0,31241432193449 | 0,314 | 0,06 | 1,35380014954564E-30 | 23 | IL32 |

| Table S3. CD14+ monocytes (T4) sub-clusters top DEG | | | | | | | |
|---|---|---|---|---|---|---|---|
| | **p_val** | **avg_logFC** | **pct.1** | **pct.2** | **p_val_adj** | **cluster** | **gene** |
| **1** | 1,0914938443678E-6 | 0,893529459171005 | 0,823 | 0,565 | 1,30062406494867E-61 | 0 | CRIP1 |
| **2** | 3,77005693712639E- | 0,406223042974106 | 0,427 | 0,301 | 4,49239984627981E-07 | 0 | AC020656.1 |
| **3** | 6,2680349353875E-4 | 0,358410294665103 | 0,971 | 0,874 | 7,46899042900775E-44 | 0 | RPL17 |
| **4** | 1,28226836975208E- | 0,283083753507702 | 0,925 | 0,803 | 1,52795098939658E-19 | 0 | S100A10 |
| **5** | 4,78045555207056E- | 0,719501040087853 | 0,711 | 0,387 | 5,69639083584728E-58 | 1 | IFITM3 |
| **6** | 1,8136742210748E-1 | 0,692406494147901 | 0,981 | 0,798 | 2,16117420183273E-112 | 1 | HLA-DPA1 |
| **7** | 1,39631746979292E- | 0,62656311701649 | 0,986 | 0,841 | 1,66385189700524E-109 | 1 | HLA-DPB1 |
| **8** | 3,80604253990361E- | 0,588748104367079 | 0,651 | 0,328 | 4,53528029054914E-53 | 1 | LY6E |
| **9** | 3,10425495840295E- | 0,573293898220459 | 1 | 0,991 | 3,69903020843295E-104 | 1 | HLA-DRA |
| **10** | 1,18852015110079E- | 0,555895040186162 | 0,997 | 0,954 | 1,4162406120517E-84 | 1 | HLA-DRB1 |
| **11** | 3,24376908229801E- | 0,547986932567587 | 0,464 | 0,254 | 3,86527523846631E-22 | 1 | ISG15 |
| **12** | 9,71632005475953E- | 0,542157910265804 | 1 | 0,995 | 1,15779669772515E-105 | 1 | CD74 |
| **13** | 3,07532355015356E- | 0,48290547117437 | 0,553 | 0,258 | 3,66455554236298E-43 | 1 | PLAC8 |
| **14** | 2,79888978274615E- | 0,447349486939218 | 0,817 | 0,535 | 3,33515706512031E-48 | 1 | PSME2 |
| **15** | 2,61591198895123E- | 0,38302880346664 | 0,315 | 0,098 | 3,11712072603429E-34 | 1 | APOBEC3A |
| **16** | 1,34006262790954E- | 0,382622148808322 | 0,817 | 0,543 | 1,59681862741701E-46 | 1 | HLA-DMA |
| **17** | 1,34194195214841E- | 0,379399876193427 | 0,594 | 0,314 | 1,59905803018004E-35 | 1 | TUBA1B |
| **18** | 4,41120032676865E- | 0,365824778528217 | 0,577 | 0,231 | 5,25638630937753E-50 | 1 | HLA-DQA1 |
| **19** | 1,03816507180879E- | 0,357926019048006 | 0,871 | 0,83 | 1,23707749956735E-12 | 1 | CD52 |
| **20** | 8,28717045372324E- | 0,346589559834607 | 1 | 0,999 | 9,87499231265661E-57 | 1 | CST3 |
| **21** | 8,71139705732846E- | 0,346063790428407 | 0,374 | 0,252 | 1,03805007335126E-07 | 1 | HLA-DRB5 |
| **22** | 4,1283871336658E-6 | 0,331480944595117 | 0,316 | 0,053 | 4,91938610847617E-62 | 1 | GBP1 |
| **23** | 1,71249656478285E- | 0,316074668669537 | 0,438 | 0,162 | 2,04061090659525E-41 | 1 | LIPA |
| **24** | 1,89580856296577E- | 0,314429123372949 | 0,386 | 0,208 | 2,25904548363001E-16 | 1 | IFI6 |
| **25** | 8,36514237636917E- | 0,313064002075417 | 0,804 | 0,518 | 9,9679036556815E-36 | 1 | HLA-DQB1 |
| **26** | 1,2042934891569E-2 | 0,311397728081417 | 0,932 | 0,881 | 1,43503612167937E-17 | 1 | SAT1 |
| **27** | 4,21373177574967E- | 0,306752115037487 | 0,942 | 0,857 | 5,02108278398331E-34 | 1 | YBX1 |
| **28** | 5,13785997640462E- | 0,305003445290153 | 1 | 1 | 6,12227394788374E-46 | 1 | ACTB |
| **29** | 2,69163982227109E- | 0,292068659864552 | 0,783 | 0,556 | 3,20735801221823E-25 | 1 | C1orf162 |
| **30** | 3,19571074485174E- | 0,280935847226779 | 0,508 | 0,283 | 3,80800892356534E-20 | 1 | VAMP5 |
| **31** | 3,77042126457475E- | 0,277378804800587 | 0,909 | 0,864 | 4,49283397886727E-16 | 1 | COTL1 |
| **32** | 6,37343890776919E- | 0,273733698942762 | 0,443 | 0,198 | 7,59458980249777E-29 | 1 | MARCKS |
| **33** | 1,50717645616149E- | 0,273287701137665 | 0,707 | 0,479 | 1,79595146516203E-22 | 1 | RNASET2 |
| **34** | 4,95332974146641E- | 0,269639620892948 | 0,963 | 0,885 | 5,90238771993138E-30 | 1 | NPC2 |
| **35** | 2,09910024731829E- | 0,265424523617959 | 0,993 | 0,987 | 2,50128785470447E-30 | 1 | AIF1 |
| **36** | 9,68850281733665E- | 0,253038285065203 | 0,298 | 0,137 | 1,15448199571384E-15 | 1 | HLA-DQA2 |
| **37** | 4,75833487105374E- | 0,252602345755062 | 0,426 | 0,164 | 5,67003183234764E-35 | 1 | ABI3 |
| **38** | 8,07615769454873E- | 1,38117447876285 | 0,997 | 0,63 | 9,62354950882426E-152 | 2 | S100A12 |
| **39** | 1,16365154039091E- | 1,33851683422664 | 1 | 0,921 | 1,38660717552981E-188 | 2 | S100A8 |
| **40** | 2,26491185222434E- | 1,16129883295754 | 1 | 0,96 | 2,69886896311053E-163 | 2 | S100A9 |
| **41** | 2,4350274843095E-6 | 0,562715757626955 | 0,997 | 0,866 | 2,9015787503032E-57 | 2 | MNDA |
| **42** | 2,57198237955293E- | 0,498153486385119 | 0,827 | 0,553 | 3,06477420347527E-42 | 2 | VCAN |
| **43** | 5,21069533289879E- | 0,376490368048936 | 0,367 | 0,138 | 6,2090645586822E-26 | 2 | FOLR3 |
| **44** | 1,34186277925534E- | 0,372927623122344 | 0,555 | 0,282 | 1,59896368776066E-28 | 2 | SELL |
| **45** | 3,35251819707068E- | 0,352635420593311 | 0,482 | 0,228 | 3,99486068362942E-25 | 2 | RBP7 |
| **46** | 4,14484687422636E- | 0,350707295712644 | 0,445 | 0,183 | 4,93899953532813E-29 | 2 | ALOX5AP |
| **47** | 8,02023138712988E- | 0,336216930761464 | 0,307 | 0,098 | 9,55690772090397E-27 | 2 | CYP1B1 |
| **48** | 2,50114370060388E- | 0,335409139582606 | 0,997 | 0,93 | 2,98036283363959E-31 | 2 | CSTA |
| **49** | 2,49977422617074E- | 0,331537953641153 | 1 | 0,992 | 2,97873096790505E-49 | 2 | S100A6 |
| **50** | 1,3120345294169E-2 | 0,324833632634993 | 0,503 | 0,306 | 1,56342034525318E-16 | 2 | HMGB2 |
| **51** | 1,54883132504243E- | 0,324202426323928 | 0,467 | 0,253 | 1,84558740692056E-18 | 2 | PLBD1 |
| **52** | 5,40252834690189E- | 0,293844224881182 | 0,96 | 0,896 | 6,43765277816829E-22 | 2 | TSPO |
| **53** | 1,05400684993731E- | 0,289596721426304 | 1 | 0,998 | 1,2559545623853E-22 | 2 | LYZ |
| **54** | 1,18870033016446E- | 0,281715465245517 | 0,653 | 0,469 | 1,41645531342397E-11 | 2 | RGS2 |
| **55** | 2,82745477329996E- | 0,268666048991524 | 0,982 | 0,924 | 3,36919510786423E-18 | 2 | ATP5MPL |
| **56** | 2,42046545070667E- | 0,260937775091137 | 0,417 | 0,233 | 2,88422663106207E-13 | 2 | MGST1 |
| **57** | 9,22188173316528E- | 0,256539807112472 | 0,995 | 0,939 | 1,09887942732398E-14 | 2 | FCN1 |
| **58** | 6,90106015160226E- | 0,251518553174646 | 0,307 | 0,178 | 8,22330327664925E-06 | 2 | RETN |

| | | | | | | | |
|---|---|---|---|---|---|---|---|
| 59 | 3,89390016879951E- | 0,502948189008066 | 0,909 | 0,728 | 4,6399714411415E-36 | 3 | RPL36A |
| 60 | 1,01268794675894E- | 0,406827397106284 | 0,956 | 0,748 | 1,20671895735795E-29 | 3 | AP1S2 |
| 61 | 3,34074913944069E- | 0,393994265608835 | 0,984 | 0,873 | 3,98083667455752E-28 | 3 | MNDA |
| 62 | 1,85603423124625E- | 0,375424403745158 | 0,862 | 0,615 | 2,21165038995304E-28 | 3 | EIF3E |
| 63 | 1,25362596831576E- | 0,33801880187084 | 0,789 | 0,567 | 1,49382070384506E-14 | 3 | ANXA1 |
| 64 | 3,29468050106143E- | 0,33424563459336 | 0,994 | 0,96 | 3,9259412850648E-26 | 3 | CTSS |
| 65 | 5,73629920849873E- | 0,333490829147186 | 1 | 0,998 | 6,83537413684709E-21 | 3 | LYZ |
| 66 | 1,94445926693156E- | 0,317494001074081 | 0,965 | 0,885 | 2,31701766247565E-22 | 3 | ITM2B |
| 67 | 1,14420960054843E- | 0,314920774171915 | 0,953 | 0,886 | 1,36344016001351E-20 | 3 | PABPC1 |
| 68 | 2,82427885670209E- | 0,310976661276726 | 0,717 | 0,481 | 3,36541068564621E-16 | 3 | ZFP36L2 |
| 69 | 2,95725650671222E- | 0,309181766575406 | 0,739 | 0,525 | 3,52386685339828E-15 | 3 | CCNI |
| 70 | 9,53486173695634E- | 0,304336384336885 | 0,965 | 0,917 | 1,13617412457572E-14 | 3 | CALM2 |
| 71 | 6,04372014685651E- | 0,283224671554667 | 0,607 | 0,376 | 7,20169692699421E-14 | 3 | CD36 |
| 72 | 5,52514444052175E- | 0,279833114375052 | 0,623 | 0,347 | 6,58376211532572E-19 | 3 | HSD17B11 |
| 73 | 3,8155703938942E-1 | 0,275075092435599 | 0,761 | 0,531 | 4,54663368136433E-13 | 3 | EVI2B |
| 74 | 2,48900553954703E- | 0,274529317310651 | 0,918 | 0,772 | 2,96589900092425E-12 | 3 | MS4A6A |
| 75 | 5,22507927952674E- | 0,266494435208175 | 0,814 | 0,632 | 6,22620446948407E-10 | 3 | LGALS2 |
| 76 | 3,40003101507596E- | 0,264619420062425 | 0,66 | 0,418 | 4,05147695756452E-14 | 3 | EIF3M |
| 77 | 4,33110360996802E- | 1,63710138060323 | 0,91 | 0,625 | 5,16094306163789E-81 | 4 | TIMP1 |
| 78 | 2,63147901475329E- | 1,27144916639156 | 0,373 | 0,014 | 3,13567039398002E-131 | 4 | C15orf48 |
| 79 | 2,20964276937607E- | 0,902143924372953 | 0,967 | 0,865 | 2,63301032398853E-48 | 4 | HLA-DPB1 |
| 80 | 2,21921459022179E- | 0,835965613152409 | 0,993 | 0,994 | 2,64441610570828E-61 | 4 | HLA-DRA |
| 81 | 6,60384312015535E- | 0,77704214305253 | 0,64 | 0,271 | 7,86913946197711E-47 | 4 | HLA-DQA1 |
| 82 | 2,31036809622299E- | 0,763255206812364 | 0,863 | 0,595 | 2,75303462345932E-50 | 4 | CD63 |
| 83 | 3,07865194608286E- | 0,746505890177086 | 1 | 0,996 | 3,66852165895233E-48 | 4 | CD74 |
| 84 | 3,8718462848918E-4 | 0,739145377463638 | 0,98 | 0,962 | 4,61369203307707E-41 | 4 | HLA-DRB1 |
| 85 | 8,73819025432478E- | 0,73762207746757 | 0,96 | 0,828 | 1,04124275070534E-36 | 4 | HLA-DPA1 |
| 86 | 4,43320491951794E- | 0,713097363748748 | 0,82 | 0,556 | 5,28260698209758E-37 | 4 | HLA-DQB1 |
| 87 | 1,13839919601117E- | 0,701767691031956 | 0,38 | 0,134 | 1,35651648196691E-26 | 4 | FABP5 |
| 88 | 1,03461767564923E- | 0,617739577150377 | 1 | 1 | 1,23285042230362E-19 | 4 | FTH1 |
| 89 | 6,46440919728061E- | 0,612858049063001 | 0,6 | 0,223 | 7,70298999947957E-50 | 4 | LITAF |
| 90 | 1,89831913553912E- | 0,564772690467471 | 0,693 | 0,473 | 2,26203708190842E-22 | 4 | CTSB |
| 91 | 9,18246992925967E- | 0,538259018818429 | 0,107 | 0,016 | 1,09418311677058E-15 | 4 | CCL2 |
| 92 | 1,02212693756064E- | 0,521262998766289 | 0,953 | 0,897 | 1,21796645879726E-40 | 4 | NPC2 |
| 93 | 3,00258795178682E- | 0,513811663144899 | 0,427 | 0,072 | 3,57788380334918E-70 | 4 | ACP5 |
| 94 | 1,18931495381024E- | 0,487723378897718 | 0,517 | 0,254 | 1,41718769896028E-23 | 4 | YWHAH |
| 95 | 3,12975540669385E- | 0,486302532193698 | 0,93 | 0,888 | 3,72941654261639E-20 | 4 | SAT1 |
| 96 | 1,5636354352169E-1 | 0,467040491116431 | 0,227 | 0,078 | 1,86322798460445E-13 | 4 | IL1B |
| 97 | 6,44308081664172E- | 0,462780049679257 | 0,39 | 0,146 | 7,67757510111028E-23 | 4 | HLA-DQA2 |
| 98 | 1,50597304382865E- | 0,461430498487485 | 0,277 | 0,004 | 1,79451747902622E-111 | 4 | SDS |
| 99 | 2,94790843311143E- | 0,448171306916832 | 0,227 | 0,094 | 3,51272768889558E-08 | 4 | CCL3 |
| 100 | 4,81588362943304E- | 0,430674725589832 | 0,4 | 0,138 | 5,73860693283241E-31 | 4 | GSN |
| 101 | 5,52315737774852E- | 0,422123930106831 | 0,14 | 0,05 | 6,58139433132514E-06 | 4 | CCL4 |
| 102 | 5,17186941017899E- | 0,419522048024996 | 0,92 | 0,965 | 6,16279958916929E-05 | 4 | SRGN |
| 103 | 2,72503237651826E- | 0,375579548297061 | 0,247 | 0,04 | 3,24714857985916E-38 | 4 | CTSL |
| 104 | 4,26728477464533E- | 0,373448457456087 | 0,483 | 0,25 | 5,08489653746737E-18 | 4 | GLUL |
| 105 | 1,03492644419186E- | 0,367906013027352 | 0,337 | 0,042 | 1,23321835089903E-66 | 4 | GPR183 |
| 106 | 5,69233980096756E- | 0,367305111657243 | 0,33 | 0,113 | 6,78299210683294E-22 | 4 | MARCKSL1 |
| 107 | 1,09372508529594E- | 0,360144977828512 | 0,35 | 0,206 | 1,30328281163865E-06 | 4 | SOD2 |
| 108 | 1,00414070396891E- | 0,355847316035862 | 0,38 | 0,134 | 1,19653406284935E-25 | 4 | CXCR4 |
| 109 | 6,32493945162127E- | 0,352728950345537 | 0,327 | 0,115 | 7,53679785055191E-21 | 4 | SERPINB9 |
| 110 | 1,74456522970849E- | 0,347304444950573 | 0,777 | 0,588 | 2,07882392772064E-13 | 4 | HLA-DMA |
| 111 | 1,46890761598431E- | 0,344248812124607 | 0,573 | 0,39 | 1,7503503152069E-10 | 4 | LDHA |
| 112 | 3,32516610154728E- | 0,343657048792347 | 0,82 | 0,698 | 3,96226792660374E-14 | 4 | GPX4 |
| 113 | 1,12304365303076E- | 0,342520116046805 | 0,2 | 0,048 | 1,33821881695146E-19 | 4 | CD9 |
| 114 | 5,0633371709976E-1 | 0,33947484837687 | 0,343 | 0,192 | 6,03347257296074E-08 | 4 | NAMPT |
| 115 | 3,02388668807582E- | 0,334388296644101 | 0,443 | 0,184 | 3,60326337751115E-24 | 4 | SPINT2 |
| 116 | 8,51903674122894E- | 0,328094467679196 | 0,35 | 0,098 | 1,01512841808484E-31 | 4 | HIF1A |

| Table S4. DEG in response to allergen challenge (MPS clusters) | | | | | | | |
|---|---|---|---|---|---|---|---|
| | **p_val** | **avg_logFC** | **pct.1** | **pct.2** | **p_val_adj** | **Cluster** | **Group** |
| **IFITM3** | 2,76597669791102E- | 0,502634238639332 | 0,561 | 0,445 | 0,0054553358412899 | 4 | Non-allergic |
| **ISG15** | 6,16927139032746E- | 0,628148467929685 | 0,424 | 0,305 | 0,0121676539631429 | 4 | Non-allergic |
| **IL1B** | 1,13150909702895E- | -0,598884416686376 | 0,138 | 0,282 | 2,23167539207021E-06 | 5 | Non-allergic |
| **SRGN** | 5,21129936654354E- | -0,335895129170165 | 0,872 | 0,935 | 1,02782457406338E-05 | 5 | Non-allergic |
| **NR4A1** | 1,20695590396027E- | -0,229837287149199 | 0,062 | 0,176 | 2,38047912938084E-05 | 5 | Non-allergic |
| **ANXA2** | 1,23373315026236E- | -0,269344878948671 | 0,625 | 0,786 | 2,43329189226246E-05 | 5 | Non-allergic |
| **TPM4** | 1,65728080898087E- | -0,309081365261008 | 0,4 | 0,545 | 0,00326865493955298 | 5 | Non-allergic |
| **TYMP** | 2,53792027753222E- | -0,225224586383707 | 0,807 | 0,874 | 0,00500554016337679 | 5 | Non-allergic |
| **FTH1** | 4,63331093097859E- | -0,202767122561551 | 0,969 | 0,991 | 0,00913827914916907 | 5 | Non-allergic |
| **AP1S2** | 5,18874042121621E- | -0,261559713517797 | 0,406 | 0,547 | 0,0102337527327647 | 5 | Non-allergic |
| **TPM3** | 5,95714509072693E- | -0,219184704252776 | 0,714 | 0,836 | 0,0117492772624407 | 5 | Non-allergic |
| **IL4I1** | 7,76369256133017E- | -0,45693403534761 | 0,156 | 0,268 | 0,0153123308387115 | 5 | Non-allergic |
| **TIMP1** | 8,15804182448836E- | -0,647312368892224 | 0,622 | 0,712 | 0,0160901058904384 | 5 | Non-allergic |
| **HSPA8** | 9,45248068142556E- | -0,233921142302165 | 0,675 | 0,788 | 0,0186431276479756 | 5 | Non-allergic |
| **PLAUR** | 1,01869275314637E- | -0,377302630771574 | 0,235 | 0,372 | 0,0200916771703059 | 5 | Non-allergic |
| **EMP3** | 1,90820904157109E- | -0,254919265003347 | 0,678 | 0,777 | 0,0376356069269065 | 5 | Non-allergic |
| **ISG15** | 3,94503884778803E- | 0,69881433094676 | 0,542 | 0,4 | 0,00778080011949234 | 13 | Non-allergic |
| **MX1** | 4,45476440599284E- | 0,441873778671678 | 0,238 | 0,108 | 0,00878613183793969 | 13 | Non-allergic |
| **DUSP1** | 9,63702923129654E- | 0,208296224018296 | 0,754 | 0,688 | 0,0190071127528862 | 13 | Non-allergic |
| **RPS29** | 5,57834462192379E- | 0,297095288737164 | 1 | 0,993 | 0,00110021690978203 | 18 | Non-allergic |
| **SRGN** | 3,57566526285166E- | -0,409341395565838 | 0,9 | 0,97 | 0,00705228459792233 | 18 | Non-allergic |
| **MVP** | 5,64932730528095E- | -0,424833915447665 | 0,212 | 0,422 | 0,0111421682442056 | 18 | Non-allergic |
| **RPL17** | 1,23838061506838E- | 0,489879737827565 | 0,931 | 0,904 | 0,0244245808709936 | 18 | Non-allergic |
| **RPL35** | 1,34961214915804E- | 0,231991672266978 | 1 | 0,978 | 0,0266184004178439 | 18 | Non-allergic |
| **CDC42** | 2,67264611893917E- | -0,688818118269134 | 0,452 | 0,897 | 0,00527125994038373 | 22 | Non-allergic |
| **DUSP1** | 8,93220310426994E- | 0,265160273535466 | 0,632 | 0,558 | 0,0176169841825516 | 4 | Allergic |
| **CEBPB** | 1,2928270128541E-( | 0,371907930197425 | 0,541 | 0,39 | 0,0254984271745215 | 4 | Allergic |
| **ALOX15** | 4,08565844055064E- | -0,407246471502403 | 0,086 | 0,234 | 8,05814414229803E-08 | 5 | Allergic |
| **HSPA1B** | 1,35325705198578E- | -0,355885636005472 | 0,157 | 0,28 | 0,000266902888363155 | 5 | Allergic |
| **CD1B** | 1,28006244123184E- | -0,37175951298942 | 0,077 | 0,155 | 0,00252466715284156 | 5 | Allergic |
| **CD1E** | 2,33554923365071E- | -0,384868000296656 | 0,439 | 0,553 | 0,00460640375352929 | 5 | Allergic |
| **CD1D** | 3,32942782578929E- | -0,24068578051583 | 0,336 | 0,458 | 0,00656663050080421 | 5 | Allergic |
| **HSPA1A** | 1,73214930210405E- | -0,378840414145196 | 0,202 | 0,317 | 0,0341631806853981 | 5 | Allergic |
| **CKB** | 2,09830093784764E- | -0,25500799273176 | 0,102 | 0,199 | 0,041384789397169 | 5 | Allergic |
| **ARF5** | 2,52548697504037E- | -0,408254065023407 | 0,643 | 0,939 | 0,00498101796087213 | 22 | Allergic |

Table S5. cDC2 (T5) sub-clusters top DEG

| | p_val | avg_logFC | pct.1 | pct.2 | p_val_adj | cluster | gene |
|---|---|---|---|---|---|---|---|
| 1 | 2,10117618430684E- | 1,18480854143705 | 0,568 | 0,255 | 2,86474360968395E-55 | 0 | C15orf48 |
| 2 | 2,72639698233204E- | 1,093101597786 | 0,907 | 0,698 | 3,71716964571151E-75 | 0 | TIMP1 |
| 3 | 2,16097726387994E- | 0,722749254220669 | 0,238 | 0,054 | 2,94627640157391E-34 | 0 | CCL17 |
| 4 | 5,88476605448684E- | 0,69172088853557 | 0,781 | 0,521 | 8,02329003868736E-46 | 0 | IFITM3 |
| 5 | 1,60936304194434E- | 0,686914263082815 | 0,63 | 0,387 | 2,19420557138691E-36 | 0 | CD1E |
| 6 | 5,2464598634665E-6 | 0,674722720053185 | 0,888 | 0,709 | 7,15302337785023E-62 | 0 | YWHAH |
| 7 | 4,51201609382751E- | 0,601784555785118 | 0,951 | 0,738 | 6,15168274232442E-76 | 0 | LITAF |
| 8 | 9,04834073207423E- | 0,60054105302323 | 0,983 | 0,91 | 1,233650775411E-85 | 0 | NPC2 |
| 9 | 9,55796708032899E- | 0,566970812263283 | 0,953 | 0,848 | 1,30313323173205E-67 | 0 | CD63 |
| 10 | 4,24442654921692E- | 0,561197124168857 | 0,123 | 0,02 | 5,78685115720235E-21 | 0 | MMP12 |
| 11 | 1,99895755446729E- | 0,491510664054717 | 0,349 | 0,147 | 2,7253787297607E-25 | 0 | CD207 |
| 12 | 5,28030963662971E- | 0,452891133399027 | 0,481 | 0,235 | 7,19917415858094E-32 | 0 | PLAUR |
| 13 | 2,72920635482294E- | 0,45170009795925 | 1 | 0,992 | 3,7209999441656E-61 | 0 | HLA-DQB1 |
| 14 | 9,33971128334972E- | 0,445855300276676 | 0,442 | 0,171 | 1,2733762363719E-43 | 0 | FPR1 |
| 15 | 1,30421121381832E- | 0,444049987506336 | 0,689 | 0,51 | 1,77816156891989E-21 | 0 | CD1C |
| 16 | 3,3033101003934E-1 | 0,439448697603327 | 0,693 | 0,604 | 4,50373299087637E-12 | 0 | HLA-DQA2 |
| 17 | 1,77295561145598E- | 0,435271513556216 | 1 | 0,999 | 2,41724768065908E-69 | 0 | HLA-DRB1 |
| 18 | 2,67435228687665E- | 0,427674875139931 | 0,994 | 0,94 | 3,64621190792763E-53 | 0 | S100A11 |
| 19 | 1,64071444907757E- | 0,423566410575756 | 0,403 | 0,084 | 2,23695007987236E-72 | 0 | FPR3 |
| 20 | 3,84376336522797E- | 0,421700289279278 | 0,266 | 0,081 | 5,24058697215182E-30 | 0 | CD1A |
| 21 | 1,99727815395805E- | 0,419583413265561 | 0,566 | 0,366 | 2,7230890351064E-20 | 0 | LGALS3 |
| 22 | 1,1257637247515E-4 | 0,407913451101376 | 0,614 | 0,34 | 1,53486626232619E-38 | 0 | SLC16A3 |
| 23 | 2,07212407035414E- | 0,40589816486662 | 1 | 1 | 2,82513395752084E-84 | 0 | HLA-DRA |
| 24 | 2,24756170500227E- | 0,404191398018288 | 0,824 | 0,643 | 3,0643256286001E-33 | 0 | SERPINB1 |
| 25 | 1,09474625455322E- | 0,396526547602006 | 0,632 | 0,411 | 1,49257704345787E-23 | 0 | CXCR4 |
| 26 | 4,53977521093956E- | 0,396507479916489 | 0,353 | 0,156 | 6,18952952259499E-24 | 0 | IL4I1 |
| 27 | 6,02433178627902E- | 0,395680247375623 | 0,598 | 0,512 | 0,000821357395741282 | 0 | FABP5 |
| 28 | 5,97220237788001E- | 0,395296204300906 | 1 | 1 | 8,1425007220016E-91 | 0 | HLA-DPB1 |
| 29 | 1,17521431472773E- | 0,394870138089431 | 0,812 | 0,634 | 1,60228719669978E-29 | 0 | CTSB |
| 30 | 4,69361694800911E- | 0,372925350768953 | 1 | 0,987 | 6,39927734691562E-44 | 0 | HLA-DQA1 |
| 31 | 3,50419348454655E- | 0,35890670549413 | 0,537 | 0,269 | 4,77761739683077E-32 | 0 | ACP5 |
| 32 | 1,58890618538549E- | 0,358422802356735 | 0,209 | 0,025 | 2,16631469315457E-46 | 0 | ALOX15 |
| 33 | 1,60385074118627E- | 0,356620223822742 | 0,925 | 0,858 | 2,18669010053336E-22 | 0 | PPA1 |
| 34 | 1,59948916715063E- | 0,356169107736922 | 0,89 | 0,762 | 2,18074353049317E-23 | 0 | LDHA |
| 35 | 8,97670726065406E- | 0,343111298509249 | 0,121 | 0,047 | 1,22388426791757E-06 | 0 | G0S2 |
| 36 | 2,71530044293785E- | 0,332492714453303 | 0,654 | 0,423 | 3,70204062390147E-28 | 0 | LIMS1 |
| 37 | 2,20587046326134E- | 0,32902484344471 | 0,957 | 0,883 | 3,00748378961051E-29 | 0 | TYMP |
| 38 | 2,23778628168484E- | 0,328633911173189 | 1 | 1 | 3,05099781644912E-65 | 0 | HLA-DPA1 |
| 39 | 5,33931646122069E- | 0,327275413277439 | 0,947 | 0,845 | 7,27962406322829E-32 | 0 | SPINT2 |
| 40 | 4,85901256291378E- | 0,325188766653359 | 0,453 | 0,222 | 6,62477772827665E-26 | 0 | CST7 |
| 41 | 9,98977581036823E- | 1,27911179819212 | 0,568 | 0,195 | 1,3620060339856E-66 | 1 | S100A9 |
| 42 | 2,84481703953486E- | 1,17670054258359 | 0,563 | 0,086 | 3,87862355170183E-125 | 1 | FCN1 |
| 43 | 9,83057188868632E- | 1,04950522008333 | 0,398 | 0,15 | 1,34030017130349E-32 | 1 | S100A8 |
| 44 | 1,12878961300927E- | 0,935916006666967 | 0,944 | 0,682 | 1,53899175837684E-83 | 1 | CD52 |
| 45 | 4,0806071222934E-8 | 0,871207241396772 | 0,998 | 0,927 | 5,56349975053482E-85 | 1 | LYZ |
| 46 | 3,46712681712956E- | 0,821040767673401 | 0,867 | 0,42 | 4,72708070247444E-93 | 1 | CSTA |
| 47 | 5,80075259775983E- | 0,820097944506791 | 0,908 | 0,489 | 7,90874609178576E-101 | 1 | C1orf162 |
| 48 | 2,79643951021194E- | 0,818742267124 | 0,837 | 0,411 | 3,81266562822296E-91 | 1 | AP1S2 |
| 49 | 6,52491991186071E- | 0,757054839797725 | 0,859 | 0,43 | 8,8960758078309E-89 | 1 | TSPO |
| 50 | 1,14893455027772E- | 0,708797223349678 | 0,922 | 0,676 | 1,56645736584864E-57 | 1 | ANXA1 |
| 51 | 1,83606081737141E- | 0,696002064892051 | 0,646 | 0,136 | 2,50328531840419E-114 | 1 | SELL |
| 52 | 8,73519002426213E- | 0,685089705478686 | 0,998 | 0,869 | 1,1909558079079E-64 | 1 | S100A4 |
| 53 | 1,53563193408957E- | 0,621712684319274 | 0,93 | 0,621 | 2,09368057893772E-60 | 1 | MNDA |
| 54 | 1,84626856520108E- | 0,583099804411518 | 0,85 | 0,499 | 2,51720256179515E-54 | 1 | CFP |
| 55 | 4,73811487655851E- | 0,49517303051811 | 0,864 | 0,505 | 6,45994582269987E-54 | 1 | CD37 |
| 56 | 2,07322771383537E- | 0,482340021013274 | 0,981 | 0,914 | 2,82663866504314E-43 | 1 | CALM2 |
| 57 | 3,03768874494988E- | 0,476624087785468 | 0,998 | 0,853 | 4,14158483486466E-50 | 1 | FCER1G |
| 58 | 1,08849289360671E- | 0,474379500019905 | 0,947 | 0,758 | 1,48405121114339E-38 | 1 | EMP3 |

| | | | | | | | |
|---|---|---|---|---|---|---|---|
| **59** | 9,19401002153862E- | 0,473742552067561 | 0,983 | 0,864 | 1,25351132633658E-36 | 1 | S100A10 |
| **60** | 1,7456354038991E-5 | 0,471001215529486 | 0,964 | 0,781 | 2,37999930967603E-46 | 1 | ANXA2 |
| **61** | 5,14596168018559E- | 0,461554794150815 | 0,925 | 0,753 | 7,01600415476503E-38 | 1 | CTSS |
| **62** | 6,41316592671989E- | 0,454168179223561 | 0,667 | 0,32 | 8,74371042448989E-46 | 1 | CASP1 |
| **63** | 1,14709816363549E- | 0,453071950584025 | 0,65 | 0,315 | 1,56395363630062E-43 | 1 | TKT |
| **64** | 1,14536821099365E- | 0,446137837219463 | 0,782 | 0,575 | 1,56159501886874E-20 | 1 | CLEC10A |
| **65** | 1,58542589046338E- | 0,443921860757393 | 0,927 | 0,801 | 2,16156965905777E-40 | 1 | EIF3E |
| **66** | 2,61167345553208E- | 0,441250211618858 | 0,667 | 0,302 | 3,56075558927244E-47 | 1 | ICAM3 |
| **67** | 2,43049097462908E- | 0,436182857103619 | 0,728 | 0,41 | 3,31373139480929E-39 | 1 | CARD16 |
| **68** | 1,78427763855373E- | 0,428821406245425 | 0,791 | 0,508 | 2,43268413240416E-35 | 1 | SAMHD1 |
| **69** | 7,2597706626237E-3 | 0,421850647624596 | 0,993 | 0,923 | 9,89797132142115E-32 | 1 | LGALS1 |
| **70** | 4,36764829682441E- | 0,399378497474148 | 0,566 | 0,235 | 5,95485168789041E-43 | 1 | GLRX |
| **71** | 3,87060045182062E- | 0,380441185342771 | 0,961 | 0,82 | 5,27717665601223E-39 | 1 | GMFG |
| **72** | 6,14963502983345E- | 0,370407436405968 | 0,473 | 0,176 | 8,38441239967492E-38 | 1 | CAPN2 |
| **73** | 2,46106395824241E- | 0,36678806844523 | 1 | 0,989 | 3,3554146006677E-43 | 1 | SLC25A6 |
| **74** | 4,43895291405918E- | 0,357023750064547 | 0,985 | 0,962 | 6,05206840302829E-31 | 1 | EEF2 |
| **75** | 2,84777430854286E- | 0,356509784661323 | 0,993 | 0,979 | 3,88265549226734E-33 | 1 | PABPC1 |
| **76** | 6,15238393278492E- | 0,35406590023019 | 0,706 | 0,467 | 8,38816025395896E-24 | 1 | LY86 |
| **77** | 3,49087255750632E- | 0,348235798788097 | 0,859 | 0,68 | 4,75945564490412E-20 | 1 | TXNIP |
| **78** | 1,12093450543556E- | 0,329506296101465 | 0,988 | 0,943 | 1,52828210471085E-29 | 1 | ARPC1B |
| **79** | 3,07307052301564E- | 0,324109907315836 | 0,728 | 0,447 | 4,18982435107952E-26 | 1 | PTPRE |
| **80** | 2,78864075383516E- | 0,319699110042992 | 0,447 | 0,161 | 3,80203280377886E-35 | 1 | AHNAK |
| **81** | 2,48935686993693E- | 0,77504543642927 | 0,864 | 0,527 | 3,39398915647201E-52 | 2 | LTB |
| **82** | 5,48641275244501E- | 0,605797892126745 | 0,995 | 0,986 | 7,48017514668352E-41 | 2 | COTL1 |
| **83** | 1,28095340646917E- | 0,581118789827392 | 0,71 | 0,44 | 1,74645187438007E-30 | 2 | PKIB |
| **84** | 2,29597545139147E- | 0,474639095417904 | 0,206 | 0,014 | 3,13033293042712E-56 | 2 | IL22RA2 |
| **85** | 7,48590447633635E- | 0,473877698217325 | 0,479 | 0,153 | 1,0206282163037E-49 | 2 | TNNI2 |
| **86** | 2,5509510776817E-1 | 0,458787162473085 | 0,4 | 0,242 | 3,47796669931122E-09 | 2 | HLA-DQB2 |
| **87** | 1,85852238948388E- | 0,398854920990711 | 0,397 | 0,17 | 2,53390942582232E-20 | 2 | CD207 |
| **88** | 9,42189193917845E- | 0,388492265619959 | 0,707 | 0,465 | 1,28458074698759E-19 | 2 | RUNX3 |
| **89** | 1,41302802537895E- | 0,385734255344063 | 0,462 | 0,191 | 1,92652240980167E-31 | 2 | SUSD3 |
| **90** | 6,06251363164846E- | 0,36508223544684 | 0,449 | 0,204 | 8,26563108538951E-26 | 2 | KCNMB1 |
| **91** | 1,32705871919168E- | 0,359644401472452 | 0,568 | 0,374 | 1,80931185774593E-16 | 2 | SMCO4 |
| **92** | 3,24974057671674E- | 0,347479386450067 | 0,412 | 0,176 | 4,43069630229561E-24 | 2 | ASB2 |
| **93** | 1,06176952315188E- | 0,345715233702198 | 0,63 | 0,413 | 1,44761656786528E-14 | 2 | SERPINB9 |
| **94** | 4,00620679728946E- | 0,344702398770734 | 0,288 | 0,039 | 5,46206234742445E-57 | 2 | CDH17 |
| **95** | 1,2722341915685E-3 | 0,343899378611691 | 0,809 | 0,48 | 1,7345640967845E-33 | 2 | PLAC8 |
| **96** | 1,48527896233481E- | 0,339389086676969 | 0,707 | 0,582 | 2,02502933724728E-07 | 2 | AC004687.1 |
| **97** | 1,51392997118598E- | 0,336504698521457 | 0,434 | 0,263 | 2,06409212271497E-11 | 2 | ITGAX |
| **98** | 5,80596436232295E- | 0,31690163719403 | 0,792 | 0,642 | 7,91585181159111E-18 | 2 | PRDX5 |
| **99** | 9,40541450841355E- | 0,314744584370808 | 0,553 | 0,359 | 1,2823342140771E-13 | 2 | PTMS |
| **100** | 4,37459405734932E- | 0,298608448656875 | 0,898 | 0,764 | 5,96432153779006E-19 | 2 | SELENOH |
| **101** | 8,5494430288831E-1 | 0,28992800446355 | 0,345 | 0,163 | 1,16563106255792E-14 | 2 | LAMP1 |
| **102** | 7,30593822444112E- | 0,285434630871694 | 1 | 1 | 9,96091617520303E-06 | 2 | MT-CO3 |
| **103** | 1,77656378594281E- | 0,274807505481638 | 0,275 | 0,058 | 2,42216706575442E-38 | 2 | PIK3R6 |
| **104** | 6,00632584402814E- | 0,2720227746059 | 0,412 | 0,175 | 8,18902465574796E-23 | 2 | HIC1 |
| **105** | 8,52131343795768E- | 0,272011307803289 | 1 | 1 | 1,16179587413115E-29 | 2 | TMSB4X |
| **106** | 3,32818868483227E- | 0,269749600655863 | 0,315 | 0,122 | 4,53765245290031E-19 | 2 | CCR6 |
| **107** | 1,47815391882117E- | 0,260232800613454 | 0,66 | 0,598 | 0,201531505292078 | 2 | NME2 |
| **108** | 9,89707240145761E- | 0,2596101937266 | 0,769 | 0,62 | 1,34936685121473E-10 | 2 | VASP |
| **109** | 1,11318579850824E- | 0,257408449364381 | 0,727 | 0,595 | 1,51771751768613E-08 | 2 | TMEM14C |
| **110** | 4,56092455572955E- | 0,256407239485576 | 0,419 | 0,219 | 6,21836453928166E-15 | 2 | ITGB7 |
| **111** | 1,26331892687698E- | 0,256083126228307 | 0,439 | 0,236 | 1,72240902490407E-14 | 2 | TLR10 |
| **112** | 5,56198714144338E- | 0,254003490827188 | 0,998 | 0,942 | 7,58321326864391E-15 | 2 | CORO1A |
| **113** | 6,61512257147399E- | 0,253882124239261 | 0,526 | 0,334 | 9,01905811394764E-13 | 2 | ITM2C |
| **114** | 2,09770868396985E- | 0,793638283150939 | 0,97 | 0,814 | 2,86001601972449E-62 | 3 | FOS |
| **115** | 1,53755950365709E- | 0,695475244822316 | 0,945 | 0,726 | 2,09630862728608E-44 | 3 | FCER1A |
| **116** | 1,68139403945843E- | 0,646989969052671 | 0,872 | 0,668 | 2,29241263339762E-40 | 3 | DUSP1 |
| **117** | 1,69500970785572E- | 0,624846979500636 | 0,73 | 0,509 | 2,31097623569049E-30 | 3 | RGS2 |
| **118** | 1,88093106664701E- | 0,546720725716747 | 0,705 | 0,316 | 2,56446141626653E-48 | 3 | FCGR2B |
| **119** | 2,04561436211774E- | 0,523146174077192 | 0,134 | 0,05 | 2,78899062131132E-05 | 3 | STATH |

| | | | | | | | |
|---|---|---|---|---|---|---|---|
| 120 | 1,01101282913002E- | 0,523013002781718 | 0,686 | 0,43 | 1,37841489123587E-23 | 3 | CEBPD |
| 121 | 1,01858567666678E- | 0,511511846420106 | 0,997 | 0,977 | 1,38873971156749E-46 | 3 | CST3 |
| 122 | 8,3917658420065E-3 | 0,509584024496684 | 0,981 | 0,911 | 1,14413335489917E-32 | 3 | JUNB |
| 123 | 1,08243180607684E- | 0,495495523347708 | 0,792 | 0,605 | 1,47578752440516E-15 | 3 | HLA-DRB5 |
| 124 | 1,61627473774444E- | 0,453716754803154 | 0,904 | 0,744 | 2,20362897744076E-20 | 3 | JUN |
| 125 | 2,28299745093412E- | 0,428073498892774 | 0,339 | 0,201 | 3,11263872460357E-06 | 3 | HSPA1A |
| 126 | 1,02076078664509E- | 0,423070198321322 | 0,825 | 0,644 | 1,39170525651191E-19 | 3 | ZFP36 |
| 127 | 1,04907515803822E- | 0,392729107534102 | 0,959 | 0,846 | 1,43030907046931E-17 | 3 | IER2 |
| 128 | 3,47474878063569E- | 0,391796415675843 | 0,27 | 0,058 | 4,7374724875187E-35 | 3 | VSIG4 |
| 129 | 2,67296564232613E- | 0,364083959357785 | 1 | 0,998 | 3,64432135674745E-23 | 3 | FTL |
| 130 | 6,82994366578517E- | 0,363778041704846 | 0,38 | 0,174 | 9,3119451939315E-16 | 3 | HSPA1B |
| 131 | 6,36091234494296E- | 0,355862574798665 | 0,336 | 0,224 | 0,00867246789109524 | 3 | AC020656.1 |
| 132 | 4,22124654775109E- | 0,349622335078601 | 0,727 | 0,515 | 5,75524754320384E-18 | 3 | CLEC7A |
| 133 | 2,50314723185471E- | 0,334922885234693 | 0,265 | 0,112 | 3,41279093591071E-12 | 3 | MAFB |
| 134 | 1,676686213545E-22 | 0,331797820491544 | 0,855 | 0,642 | 2,28599398354725E-18 | 3 | MNDA |
| 135 | 9,05503246479406E- | 0,325722930919907 | 0,981 | 0,931 | 1,23456312625002E-09 | 3 | LYZ |
| 136 | 7,53589684582228E- | 0,30922057520702 | 0,557 | 0,368 | 1,02744417595941E-10 | 3 | FOSB |
| 137 | 2,17503753508805E- | 0,300623763764218 | 0,21 | 0,082 | 2,96544617533904E-10 | 3 | CCL3 |
| 138 | 1,21240361188338E- | 0,299672368472736 | 0,276 | 0,126 | 1,6529910844418E-09 | 3 | CD14 |
| 139 | 1,6686304131288E-1 | 0,298544033666936 | 0,79 | 0,566 | 2,2750107052598E-13 | 3 | IFITM3 |
| 140 | 5,55466247031237E- | 0,295872009640276 | 0,762 | 0,634 | 7,57322681202389E-09 | 3 | RNASE6 |
| 141 | 3,35702362479142E- | 0,289298450292096 | 0,653 | 0,476 | 4,57696601004062E-10 | 3 | MEF2C |
| 142 | 7,89058606098097E- | 0,270145398282957 | 0,276 | 0,099 | 1,07580250355415E-17 | 3 | NR4A2 |
| 143 | 1,85697167448441E- | 0,260713368051182 | 0,298 | 0,125 | 2,53179518099204E-13 | 3 | SGK1 |
| 144 | 7,62442179741455E- | 0,256019457527464 | 0,773 | 0,582 | 1,0395136678595E-10 | 3 | CLEC10A |
| 145 | 1,43670713990686E- | 0,255272059839612 | 0,992 | 0,927 | 1,95880651454901E-12 | 3 | AIF1 |
| 146 | 2,98043112617812E- | 0,254041673815634 | 0,156 | 0,075 | 0,00406351979743125 | 3 | HSPA6 |
| 147 | 2,10081017545823E- | 0,253219488638982 | 0,393 | 0,211 | 2,86424459321975E-11 | 3 | ATP2B1-AS1 |
| 148 | 4,69188222842251E- | 0,251092327871706 | 0,866 | 0,777 | 6,39691223023124E-07 | 3 | NEAT1 |
| 149 | 2,74367878011336E- | 0,250705428432901 | 0,926 | 0,888 | 3,74073164880655E-09 | 3 | ITM2B |
| 150 | 9,65574267045864E- | 1,72744093104607 | 0,291 | 0,004 | 1,31646395569033E-115 | 4 | PTGDS |
| 151 | 3,84052609338824E- | 1,24229705635505 | 0,939 | 0,497 | 5,23617327572553E-77 | 4 | PLAC8 |
| 152 | 8,40206102122295E- | 1,18301115882545 | 0,282 | 0,114 | 1,14553699963354E-09 | 4 | GZMB |
| 153 | 5,94163727704335E- | 1,07224183237474 | 0,93 | 0,441 | 8,1008282635209E-73 | 4 | PPP1R14A |
| 154 | 4,17672372409059E- | 1,06209492253532 | 0,592 | 0,165 | 5,69454512542511E-57 | 4 | SOX4 |
| 155 | 7,8504997155016E-5 | 1,0069759261354 | 0,446 | 0,089 | 1,07033713121149E-54 | 4 | JCHAIN |
| 156 | 9,83868347702021E- | 0,996290784164789 | 0,662 | 0,177 | 1,34140610525693E-69 | 4 | C12orf75 |
| 157 | 4,37867413128133E- | 0,966411503253062 | 0,362 | 0,035 | 5,96988431058897E-73 | 4 | HAMP |
| 158 | 6,28754432106757E- | 0,940845925198129 | 0,549 | 0,031 | 8,57243792734352E-158 | 4 | LILRA4 |
| 159 | 1,68777712058801E- | 0,926475984915176 | 0,648 | 0,173 | 2,30111532620969E-68 | 4 | SERPINF1 |
| 160 | 2,77637036320527E- | 0,855657329154429 | 0,709 | 0,351 | 3,78530335319407E-36 | 4 | IGKC |
| 161 | 2,18700480440242E- | 0,842491603012576 | 0,817 | 0,562 | 2,98176235032225E-21 | 4 | LTB |
| 162 | 4,14583167375328E- | 0,789975527366981 | 0,7 | 0,13 | 5,65242690399522E-102 | 4 | TCF4 |
| 163 | 7,05415926309296E- | 0,771486523455167 | 0,535 | 0,265 | 9,61764073930094E-20 | 4 | IRF8 |
| 164 | 3,25218773837026E- | 0,750734709136861 | 0,878 | 0,506 | 4,43403276249402E-40 | 4 | ALOX5AP |
| 165 | 2,3058991601143E-6 | 0,738537392319566 | 0,648 | 0,193 | 3,14386291489984E-58 | 4 | LGMN |
| 166 | 4,77816892290234E- | 0,733012105078623 | 0,751 | 0,352 | 6,51455550948505E-42 | 4 | PLP2 |
| 167 | 6,47294598504118E- | 0,729260626585098 | 0,643 | 0,174 | 8,82521455600514E-63 | 4 | APP |
| 168 | 1,56057871558914E- | 0,719873946964602 | 0,977 | 0,876 | 2,12769302083424E-28 | 4 | S100A10 |
| 169 | 4,83985415550692E- | 0,702423774161673 | 0,981 | 0,96 | 6,59865715561813E-25 | 4 | S100A6 |
| 170 | 4,39169090251101E- | 0,668513882682732 | 0,563 | 0,168 | 5,98763137648351E-46 | 4 | CYB561A3 |
| 171 | 1,20896915620718E- | 0,667537639051321 | 0,653 | 0,319 | 1,64830854757287E-26 | 4 | AXL |
| 172 | 3,35345340176012E- | 0,653266021426929 | 0,549 | 0,127 | 4,57209836795975E-57 | 4 | STMN1 |
| 173 | 3,82704331825519E- | 0,637275124771305 | 0,761 | 0,368 | 5,21779086010912E-41 | 4 | SEPT9 |
| 174 | 3,52457616707658E- | 0,621696285583941 | 0,812 | 0,44 | 4,80540714619221E-37 | 4 | SEPT6 |
| 175 | 1,36511980608222E- | 0,604477170752204 | 0,629 | 0,341 | 1,86120434361249E-20 | 4 | ITM2C |
| 176 | 2,84165498534082E- | 0,600757048106728 | 0,441 | 0,048 | 3,87431240701367E-84 | 4 | CLIC3 |
| 177 | 1,07200870958363E- | 0,590415876773413 | 0,526 | 0,14 | 1,46157667464632E-46 | 4 | CD2 |
| 178 | 2,8393438400682E-3 | 0,570686292593241 | 0,535 | 0,18 | 3,87116139154898E-35 | 4 | CSF2RB |
| 179 | 1,45459580288526E- | 0,559105633897085 | 0,362 | 0,026 | 1,98319591765376E-87 | 4 | TSPAN13 |
| 180 | 6,66328204084989E- | 0,558242074063048 | 0,545 | 0,098 | 9,08471873449474E-74 | 4 | CXCR3 |

| | | | | | | | |
|---|---|---|---|---|---|---|---|
| 181 | 3,34194797062714E- | 0,554254941568178 | 0,577 | 0,326 | 4,55641186315305E-13 | 4 | ATF5 |
| 182 | 3,38930259023496E- | 0,540729284909588 | 0,465 | 0,053 | 4,62097515152634E-87 | 4 | IL3RA |
| 183 | 5,60896583414479E- | 0,533861971120695 | 0,967 | 0,885 | 7,64726401827301E-11 | 4 | S100A4 |
| 184 | 2,06742046890389E- | 0,532916750134806 | 0,883 | 0,724 | 2,81872106730356E-17 | 4 | HERPUD1 |
| 185 | 3,01819120757452E- | 0,499324085121626 | 0,408 | 0,076 | 4,1150018924071E-50 | 4 | PLA2G16 |
| 186 | 7,74771048850429E- | 0,483832030481453 | 0,685 | 0,373 | 1,05632284800268E-23 | 4 | CCND3 |
| 187 | 1,74703243415299E- | 0,465633166064046 | 0,427 | 0,054 | 2,38190402072418E-72 | 4 | SUSD1 |
| 188 | 1,19831842719596E- | 0,451151083062212 | 0,526 | 0,197 | 1,63378734363898E-24 | 4 | SELL |
| 189 | 4,21639799019706E- | 0,445133939242187 | 0,474 | 0,107 | 5,74863701983467E-47 | 4 | UGCG |
| 190 | 9,40779191840466E- | 2,32252387659788 | 0,694 | 0,045 | 1,28265835015529E-142 | 5 | CCL22 |
| 191 | 1,1912409549455E-3 | 2,10874848041958 | 0,435 | 0,096 | 1,6241379179727E-26 | 5 | CCL17 |
| 192 | 4,52046983647198E- | 2,05545352112555 | 0,917 | 0,028 | 6,1632085750459E-280 | 5 | CCR7 |
| 193 | 8,37016859759177E- | 1,99121922774474 | 0,926 | 0,097 | 1,14118878659566E-151 | 5 | BIRC3 |
| 194 | 2,92191112319582E- | 1,85710977957696 | 0,917 | 0,184 | 3,98373362536518E-94 | 5 | FSCN1 |
| 195 | 4,26835533094482E- | 1,82535252681575 | 0,889 | 0,206 | 5,81947565821017E-78 | 5 | CD83 |
| 196 | 2,25700194276144E- | 1,69464503165486 | 0,972 | 0,693 | 3,07719644876094E-40 | 5 | TXN |
| 197 | 1,58276995741787E- | 1,53918292156671 | 0,472 | 0,05 | 2,15794855994352E-60 | 5 | G0S2 |
| 198 | 2,23869869606466E- | 1,51003816345539 | 0,917 | 0,347 | 3,05224180221456E-52 | 5 | MARCKSL1 |
| 199 | 2,57930783678116E- | 1,43508452528932 | 0,528 | 0,023 | 3,51662830466743E-129 | 5 | CCL19 |
| 200 | 1,9013749192667E-1 | 1,35729263701903 | 0,907 | 0,111 | 2,59233456492822E-125 | 5 | LAMP3 |
| 201 | 6,13861369758369E- | 1,31161051794536 | 0,713 | 0,101 | 8,3693859152856E-83 | 5 | TMEM176A |
| 202 | 1,43844683091959E- | 1,29711162967593 | 0,972 | 0,674 | 1,96117840927576E-46 | 5 | BTG1 |
| 203 | 7,93895073409616E- | 1,29486282239949 | 0,843 | 0,141 | 1,08239654308667E-92 | 5 | MARCKS |
| 204 | 1,88477907010091E- | 1,12411141882118 | 0,583 | 0,055 | 2,56970778417559E-88 | 5 | EBI3 |
| 205 | 7,32030082473741E- | 1,08583654644262 | 0,741 | 0,163 | 9,98049814444699E-60 | 5 | SAMSN1 |
| 206 | 5,08628324133139E- | 1,081491247888 | 0,657 | 0,13 | 6,93463857123122E-52 | 5 | TMEM176B |
| 207 | 3,37105760336379E- | 1,07481539352453 | 0,694 | 0,196 | 4,59609993642618E-41 | 5 | BCL2A1 |
| 208 | 9,26453565409629E- | 1,06087196068526 | 0,741 | 0,407 | 1,26312679107949E-22 | 5 | RPS27L |
| 209 | 1,98115928526767E- | 1,0329385288105 | 0,907 | 0,533 | 2,70111256953394E-32 | 5 | PNRC1 |
| 210 | 1,14131079696187E- | 1,01900715199183 | 0,935 | 0,828 | 1,55606314057782E-14 | 5 | CRIP1 |
| 211 | 9,37025468713564E- | 0,968487253950587 | 0,954 | 0,609 | 1,27754052404407E-32 | 5 | ID2 |
| 212 | 4,67083701378684E- | 0,963262570398729 | 0,741 | 0,191 | 6,36821918459698E-41 | 5 | IL4I1 |
| 213 | 2,40963371055609E- | 0,936519997304515 | 0,704 | 0,157 | 3,28529460097218E-50 | 5 | CD40 |
| 214 | 2,17438836980372E- | 0,931369139705837 | 0,806 | 0,181 | 2,96456110339039E-63 | 5 | RASSF4 |
| 215 | 9,59975901615106E- | 0,926435180945927 | 0,648 | 0,009 | 1,30883114426204E-244 | 5 | LAD1 |
| 216 | 1,98430166516129E- | 0,904152126408969 | 0,731 | 0,195 | 2,7053968902809E-43 | 5 | CDKN1A |
| 217 | 2,47391891847859E- | 0,90317831878166 | 1 | 0,91 | 3,37294105345371E-34 | 5 | GPX4 |
| 218 | 5,41531300850276E- | 0,899439528547946 | 0,769 | 0,102 | 7,38323775579267E-94 | 5 | RAB9A |
| 219 | 1,98100577834537E- | 0,883095866262983 | 0,648 | 0,277 | 2,70090327819608E-18 | 5 | CST7 |
| 220 | 1,89867443101723E- | 0,86440832118548 | 0,657 | 0,13 | 2,5886527192489E-50 | 5 | IL7R |
| 221 | 4,51698760093968E- | 0,851964603157232 | 0,926 | 0,598 | 6,15846089512116E-31 | 5 | CSF2RA |
| 222 | 5,46727002954473E- | 0,848249047280659 | 0,843 | 0,445 | 7,45407595828128E-23 | 5 | BASP1 |
| 223 | 1,03997269936266E- | 0,841528287280828 | 0,926 | 0,525 | 1,41789877831105E-21 | 5 | RGS1 |
| 224 | 4,97173117554328E- | 0,807139736255554 | 0,796 | 0,411 | 6,77845828473571E-20 | 5 | GADD45B |
| 225 | 1,90322191888125E- | 0,795499792914125 | 0,565 | 0,056 | 2,59485276420269E-80 | 5 | TRAF1 |
| 226 | 9,60045128574719E- | 0,789481575831667 | 0,519 | 0,084 | 1,30892552829877E-48 | 5 | DUSP5 |
| 227 | 3,80748520183596E- | 0,788399306897011 | 0,241 | 0,066 | 5,19112532418315E-08 | 5 | CD1B |
| 228 | 4,2707433638648E-4 | 0,787507117944838 | 0,676 | 0,182 | 5,82273150229326E-38 | 5 | TNFAIP2 |
| 229 | 7,09031501353569E- | 0,786489668125095 | 0,5 | 0,345 | 0,0966693548945456 | 5 | C15orf48 |
| 230 | 2,08090424502726E- | 1,86967910472753 | 0,589 | 0,101 | 2,83710484767017E-39 | 6 | S100B |
| 231 | 1,06619933722113E- | 1,55376118463183 | 0,575 | 0,363 | 0,0145365617636729 | 6 | HIST1H4C |
| 232 | 2,69938918485589E- | 1,32274973845838 | 0,603 | 0,267 | 3,68034721463253E-11 | 6 | HMGB2 |
| 233 | 1,91830671518222E- | 1,25486823243083 | 0,63 | 0,151 | 2,61541937547944E-29 | 6 | STMN1 |
| 234 | 1,78399166179977E- | 1,21588720949975 | 0,836 | 0,578 | 2,43229423169781E-12 | 6 | LTB |
| 235 | 6,06328122986117E- | 1,13953230706995 | 0,932 | 0,745 | 8,26667762879272E-17 | 6 | H2AFZ |
| 236 | 3,63846101774353E- | 1,10022709205129 | 0,548 | 0,018 | 4,96067775159153E-131 | 6 | PCLAF |
| 237 | 4,77071869070726E- | 1,08273954030816 | 0,808 | 0,619 | 6,50439786291028E-10 | 6 | ID2 |
| 238 | 9,21284665164021E- | 1,05239616174822 | 0,74 | 0,638 | 0,00125607951248463 | 6 | TUBB |
| 239 | 4,97601249168744E- | 0,993311179092869 | 0,466 | 0,003 | 6,78429543116665E-186 | 6 | TYMS |
| 240 | 2,10574821606734E- | 0,95017558326821 | 0,945 | 0,849 | 2,87097711778621E-15 | 6 | GSN |
| 241 | 1,42330836121244E- | 0,870278854660542 | 0,274 | 0,004 | 1,94053861967704E-91 | 6 | UBE2C |

| | | | | | | | |
|---|---|---|---|---|---|---|---|
| **242** | 3,04260815261741E- | 0,816967848569512 | 0,863 | 0,813 | 0,0414829195527857 | 6 | HMGN2 |
| **243** | 1,31459868731828E- | 0,788476888871008 | 0,575 | 0,082 | 1,79232385028975E-42 | 6 | CA2 |
| **244** | 2,68969777317466E- | 0,772167217181984 | 0,616 | 0,32 | 3,66713394394633E-08 | 6 | DUT |
| **245** | 2,18092491866485E- | 0,747747688857385 | 0,26 | 0,009 | 2,97347303410766E-56 | 6 | PIGR |
| **246** | 1,97611612381655E- | 0,72596178723002 | 0,986 | 0,987 | 2,69423672321148E-10 | 6 | COTL1 |
| **247** | 5,55162496166729E- | 0,678019176567431 | 0,521 | 0,126 | 7,56908547273719E-22 | 6 | CKS1B |
| **248** | 9,45838811468578E- | 0,635884847844646 | 0,466 | 0,132 | 1,28955663555626E-14 | 6 | PCNA |
| **249** | 1,06062048403624E- | 0,634696592321329 | 0,342 | 0,016 | 1,44604996793502E-61 | 6 | IGF1 |
| **250** | 1,65898094728192E- | 0,630554276122835 | 0,959 | 0,905 | 0,0226185462352417 | 6 | HMGB1 |
| **251** | 1,70562669981275E- | 0,622152791055803 | 0,795 | 0,516 | 2,3254514425247E-10 | 6 | RANBP1 |
| **252** | 4,22258246353865E- | 0,610451121692172 | 0,726 | 0,382 | 5,75706893078859E-10 | 6 | PTMS |
| **253** | 3,57481760223733E- | 0,588735120282141 | 1 | 0,999 | 4,87390631889038E-11 | 6 | GAPDH |
| **254** | 1,36098836507597E- | 0,586772073917919 | 0,932 | 0,738 | 1,85557153694458E-06 | 6 | CKLF |
| **255** | 6,438341638674E-43 | 0,577759328849489 | 0,466 | 0,058 | 8,77803499016813E-39 | 6 | MCM7 |
| **256** | 9,10791245212941E- | 0,576127642587008 | 0,877 | 0,623 | 1,24177278372332E-09 | 6 | LDHB |
| **257** | 7,0237675974661E-4 | 0,574987795796992 | 0,534 | 0,075 | 9,57620474238529E-39 | 6 | GPR82 |
| **258** | 9,00711795272732E- | 0,574426201353249 | 0,411 | 0,009 | 1,22803046167484E-112 | 6 | TK1 |
| **259** | 1,19380180092778E- | 0,571817776906438 | 0,767 | 0,374 | 1,62762937538494E-13 | 6 | MZT2A |
| **260** | 5,94383790698511E- | 0,570658692453286 | 0,836 | 0,497 | 8,1038286023835E-08 | 6 | RUNX3 |
| **261** | 1,68377202644744E- | 0,567204348955402 | 0,89 | 0,907 | 0,0229565478085844 | 6 | CTSH |
| **262** | 1,25028666155092E- | 0,558998011850773 | 0,301 | 0,037 | 1,70464083435852E-23 | 6 | TPM2 |
| **263** | 3,19439811855652E- | 0,538853796486993 | 0,589 | 0,115 | 4,35524239483997E-29 | 6 | TMEM97 |
| **264** | 2,05175660597856E- | 0,521760967149725 | 0,205 | 0,015 | 2,79736495659116E-25 | 6 | CAMP |
| **265** | 2,79880954993747E- | 0,519100965163497 | 1 | 0,994 | 3,81589694038475E-12 | 6 | ACTG1 |
| **266** | 3,41576261727287E- | 0,504016728187442 | 0,425 | 0,071 | 4,65705075238983E-24 | 6 | CDH17 |
| **267** | 1,95211077286919E- | 0,501662200932342 | 0,616 | 0,415 | 0,0266150782772985 | 6 | ANP32B |
| **268** | 7,23296710819651E- | 0,498561295051062 | 0,301 | 0,022 | 9,86142735531512E-39 | 6 | NUSAP1 |
| **269** | 1,16492350302707E- | 0,495193972589017 | 0,849 | 0,644 | 1,58825670402711E-07 | 6 | RAN |
